# A 10-Year Longitudinal Study of Brain Cortical Thickness in People with First-Episode Psychosis using Normative Models

**DOI:** 10.1101/2024.04.19.24306008

**Authors:** Pierre Berthet, Beathe C. Haatveit, Rikka Kjelkenes, Amanda Worker, Seyed Mostafa Kia, Thomas Wolfers, Saige Rutherford, Dag Alnaes, Richard Dinga, Mads L. Pedersen, Andreas Dahl, Sara Fernandez-Cabello, Paola Dazzan, Ingrid Agartz, Ragnar Nesvåg, Torill Ueland, Ole A. Andreassen, Carmen Simonsen, Lars T. Westlye, Ingrid Melle, Andre Marquand

## Abstract

**Background:** Clinical forecasting models have potential to optimize treatment and improve outcomes in psychosis, but predicting long-term outcomes is challenging and long-term follow up data are scarce. In this 10-year longitudinal study we aimed to characterize the temporal evolution of cortical correlates of psychosis and their associations with symptoms.

**Design:** Structural MRI from people with first-episode psychosis and controls (n=79 and 218) were obtained at enrollment, after 12 months (n=67 and 197), and 10 years (n=23 and 77), within the Thematically Organized Psychosis (TOP) study. Normative models for cortical thickness estimated on public MRI datasets (n=42983) were applied to TOP data to obtain deviation scores for each region and timepoint. Positive And Negative Syndrome Scale (PANSS) scores were acquired at each timepoint along with registry data. Linear mixed effects (LME) models assessed effects of diagnosis, time and their interactions on cortical deviations plus associations with symptoms.

**Results:** LMEs revealed conditional main effects of diagnosis and time x diagnosis interactions in a distributed cortical network, where negative deviations in patients attenuate over time. In patients, symptoms also attenuate over time. LMEs revealed effects of anterior cingulate on PANSS total, and insular and orbitofrontal regions on PANSS negative scores.

**Conclusions:** This long-term longitudinal study revealed a distributed pattern of cortical differences which attenuated over time together with a reduction in symptoms. These findings are not in line with a simple neurodegenerative account of schizophrenia, and deviations from normative models offer a promising avenue to develop biomarkers to track clinical trajectories over time.

## INTRODUCTION

Psychotic disorders are severe and complex conditions characterized by substantial clinical and biological heterogeneity^1–3^ and significant negative effects on quality of life and societies^4–7^. Predicting long-term outcomes and improve treatment and prognosis is a priority in schizophrenia research, and models that can predict the clinical course are highly needed, as this will help to optimize treatment planning. Longitudinal clinical studies have revealed substantial heterogeneity in the clinical and functional trajectories^8–12^, underlying neurobiology^13–16^, and its interaction with medication^17–21^. Likewise, both cross-sectional and longitudinal brain imaging studies have revealed significant yet typically diffuse brain cortical alterations in groups of individuals with psychotic disorders^22–24^. Both neurodevelopmental and neurodegenerative models have been proposed for the evolution of these changes^25–27^, and there is increasing evidence and awareness of substantial individual differences and heterogeneity in such trajectories^28–30^. However, the timeframe for most longitudinal studies is relatively short (e.g. 1-2 years) and there is a need for better characterization of the dynamics of the brain cortical correlates of the illness and their long-term temporal associations with clinical symptoms at an individual level^31–35^. Two previous long-term prospective studies of first-episode psychosis patients were conducted more than twenty years ago.^36,37^ These studies comprised patients receiving first-generation antipsychotics, a group of medications associated with findings of reduced cortical thickness.^18,38^ A more recent study reports an association between increasing expressive negative symptoms and changes in cortical thickness, reporting on the dose but not the type of medication.^39^

More recently still, the availability of large neuroimaging datasets has led to the advent of normative development charts^40,41^ which allow for individual-level statistical inference and for mapping clinical traits to extreme deviations from the normative range.^28,42–49^ Such techniques may be particularly valuable in longitudinal studies because they provide the ability to detect deviations from an expected trajectory over time, which might provide early indicators of worsening or improvement in the disease course and can accommodate heterogeneity in the pattern of atypicalities across individuals and timepoints.

Our main goal in this study was to map the associations between brain cortical abnormalities and clinical symptoms over the longer term. To achieve this, we applied a normative modeling approach to MRI-based estimates of cerebral cortical thickness of people with schizophrenia spectrum first-episode psychosis and healthy controls in a long-term longitudinal study of participants with follow-up after approximately 12 months and 10 years. We used normative models to compute individual deviation scores for the two groups at different time points, which allows meaningful comparisons even when follow up data are acquired on different scanners from the baseline scans^50^. Then, we assessed the association between cortical thickness deviations and symptom scales at clinical follow-up and between deviations and Norwegian patient registry data to address the possibility of selective retention bias influencing our findings. Given prior evidence for the heterogeneity of cortical alterations in schizophrenia^28–30^ and that only a subset of individuals with schizophrenia show progressive brain changes,^36^ we predicted that: (i) we would observe a characteristic yet diffuse pattern of case-control differences in cortical normative deviations, consistent with prior studies^22,24^, and (ii) that individual differences in cortical deviations would be coupled to clinical outcome over time. We tested these associations using linear mixed models (LMEs) with subsequent corrections for multiple comparisons.

## METHODS

### Participants

All participants were recruited to a specific first-episode sub-study of the Thematically Organized Psychosis study at the University of Oslo and Oslo University Hospital from October 27, 2004, to October 17, 2012. Here, patients with a first-episode schizophrenia spectrum diagnosis (SCZ) were consecutively recruited from the catchment-area-based inpatient and outpatient services at Oslo University Hospital and three additional hospitals in the larger Oslo area to the prospective study. Psychiatric diagnosis at baseline was established using the Structured Clinical Interview for DSM-IV Axis I Disorders (SCID-I^40^), and we included a broad range of schizophrenia spectrum diagnoses: schizophrenia (n=57, 72% of the final, quality-checked longitudinal sample), schizophreniform disorders (n=18, 23%) and schizoaffective (n=4, 5%). Information about patients’ current antipsychotic medication was gathered at each time point. Positive and negative symptoms were assessed using the Positive and Negative Syndrome Scale (PANSS^51^). Healthy controls (CTRL) from the same geographic catchment area were invited based on national records. Exclusion criteria for healthy controls included a history of drug or alcohol abuse or dependency, psychosis, bipolar disorder, or major depressive disorder, or having a first degree relative diagnosed with a psychotic or bipolar disorder. The participants were invited to a follow-up approximately 10 years after their baseline scan (patients mean [SD] 9.7 years [0.9], CTRL 8.2 years [1.1]). A subsample also participated in a follow-up scan after approximately one year.^16^ See supplementary Figure 2 for details. We also augmented our sample by including additional controls from additional TOP sub-studies acquired on the same scanners at the same time, in order to improve the fit of the normative models we employ.

We also accessed the Norwegian National Registry for health care information about all enrolled patients at baseline. This allowed us to access dates and durations of contacts with the Norwegian healthcare system for ICD-10 F-01-09 labeled events (Mental, Behavioral and Neurodevelopmental disorders) in the follow-up period (e.g. from the start of treatment to 10-year follow-up) for all participants, serving as a proxy for illness severity.

### MRI data acquisition and analysis

Three scanners at Oslo University Hospital were used in this longitudinal study without temporal overlap. The first scanner was a 1.5 Tesla Siemens MANETOM Sonata scanner with a 32-channel head coil. T1-weighted images were acquired using a MPRAGE sequence using these parameters: repetition time (TR) = 2.730 ms, echo time (TE) = 3.93 ms, flip angle (FA) = 7 degrees. The second scanner was a 3 Tesla GE Signa HDxT with a 8HRBRAIN coil. T1-weighted images were acquired using a FSPGR sequence, with the following parameters: TR = 7.8ms, TE = 3.18 ms, and FA = 12 degrees. The third scanner was a 3 Tesla GE 750 Discovery scanner with a 32-channel head coil. The T1-weighted images were here acquired using a BRAVO sequence, with the following parameters: TR = 8.16 ms, TE = 3.18 ms, FA = 12 degrees. See supplementary figure 9 for the distribution of scanners across different timepoints.

T1-weighted structural MRI scans were preprocessed through Freesurfer (version 5.3), and cortical thickness measures were parcellated using the Destrieux atlas^52^. An automatic quality check procedure based on the Freesurfer Euler characteristic was run on all data and samples with a value higher than five were removed.^53–58^

### Normative Modeling

To account for site and scanner effects we used the Hierarchical Bayesian regression (HBR) approach for normative modeling^54,55,59^, which efficiently accommodates inter-site variation and provides computational scaling, which is useful for multi-cohort and longitudinal studies with data from different scanners. We estimated a normative model for each region of interest (ROI, n=150) in the Freesurfer Destrieux atlas^52^, using HBR with age as a covariate, and sex and scanner id as batch effects, to predict cortical thickness^45,54,55,58^. This accommodated multi-site pooling using transfer learning and comparisons across scanners^54,55,59^. The deviations from these models were then used as features in the linear mixed models outlined below. Importantly, as HBR fits site-specific intercept and slopes, the resulting normative trajectory might not be linear across the lifespan, but rather piecewise linear. Using pooled data from a collection of mostly publicly available datasets from 77 sites, and n=40,435 participants^29^, the reference normative models were first trained on (95%) healthy individuals and validated on an independent set of n=2548 controls and patients (5%, stratified by sites). We then adapted the model to the three unseen Oslo scanners, by transferring the (hyper)parameters as informed priors for these new sites^55^. For this adaptation step, we used held-out cross-sectional data from CTRL from these three scanners, following methods described previously^55^ (eTable 1 in the supplement). After this transfer step, we tested the normative models on the remaining longitudinal CTRL and SCZ samples and obtained individual deviation scores for these participants at each time point, and ROI. We defined the threshold for extreme deviation values as |z| > 2.0. While this threshold is arbitrary, we consider 2 standard deviations from the mean a potentially clinically significant effect.

### Statistical analysis

To test for the potential of non-random attrition confounding our findings, we applied t-tests to check for differences between the patients followed for 10-years and the ones that dropped-out in several factors, *i.e.* number of hospitalizations, PANSS domain scores, and median cortical thickness deviation scores.

Next, we employed a linear mixed-effects (LME) model to investigate the impact of diagnosis, time since inclusion, age at inclusion, and sex on the deviation scores derived from the 150 cortical ROIs in a longitudinal setting. The interaction between group and time since inclusion (delay) was also included. The model was formulated as follows:

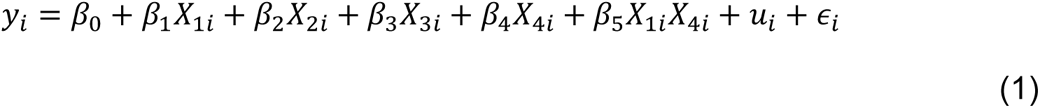

Where *i* indexes subjects, *y*_*i*_ is the deviation score at a given ROI and *β*_0_ a global intercept. The variables *X*_1*i*_, …, *X*_4*i*_ represent respectively, time since inclusion, age at baseline, sex and diagnosis with associated coefficients *β*_1_, …, *β*_4_. In addition, we model an interaction between diagnosis and time since inclusion, i.e., *X*_1*i*_*X*_4*i*_ with coefficient *β*_5_. Finally, *u*_*i*_ is a subject-specific random intercept and *∈*_*i*_ are normally distributed errors.

In all instances where multiple comparison correction was required, we applied the Benjamini-Hochberg procedure with *⍺*=.05 to control the false discovery rate^60^ corrected across ROIs. For the ROIs with a significant interaction effect, we also calculated the predicted values for a combination of time since inclusion (delay) and diagnosis levels to visualize the nature of the interaction effects in the model.

We also aimed to understand the regional distribution of extreme deviations at the level at the individual. Since these are count data (i.e. having highly skewed discrete distributions), we applied a non-parametric Wilcoxon test to the proportion of individual deviations differing between diagnostic groups at each time point, in line with prior work.^28,29,61^

To examine the development of PANSS scores during the longitudinal period we visualized the distributions and used a LME model to assess the change in PANSS subscales over time, while controlling for age at baseline and sex. The model was formulated as follows:

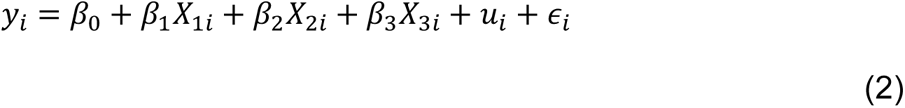

Here, *y*_*i*_ is the PANSS score (or subscale) and *X*_1*i*_, …, *X*_3*i*_ are defined as above, respectively time since inclusion, age at baseline and sex with coefficients *β*_1_, …, *β*_3_. Again, *β*_0_ is the global intercept, *u*_*i*_ is a subject-specific random intercept and *∈*_*i*_ are normally distributed errors.

Next, we employed another linear mixed-effect (LME) model to investigate the associations between the deviation score in the different ROIs, time since inclusion, age at inclusion and sex on the general and domain specific PANSS scores in the schizophrenia patients. The model was formulated as follows:

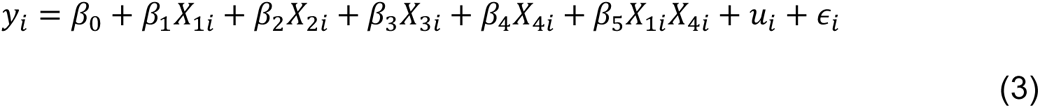

Here, *y*_*i*_, *X*_1*i*_, …, *X*_3*i*_, *u*_*i*_ and *∈*_*i*_ are defined as in equations (1) and (2), but here *X*_4*i*_ is the deviation score at a given ROI, with coefficient *β*_4_, and we also model an interaction between time since inclusion and deviation score, *X*_1*i*_*X*_4*i*_, with coefficient *β*_5_.

## RESULTS

### Participants

A total of 218 healthy controls and 79 patients were included in the longitudinal analysis (Table 1 and supplementary Figures 1 and 8). There was no significant difference either in the number of contacts with the healthcare system for ICD-10 classified “Mental, Behavioral and Neurodevelopmental disorders”, nor in duration of contacts with the healthcare system (e.g. the duration of treatment) between the longitudinal sample and the drop-outs. There was no significant association between attrition groups and PANSS scores, or median cortical deviation scores. Data on medication use for each time point showed that patients primarily used second-generation antipsychotics (62% at baseline, 48% at one year, and 17% at ten years) or did not use any antipsychotics (38%, 50%, and 83%, respectively, for details, see supplementary materials).

**Table 1:** Demographics of the participants at the three time points. *:There was a significant effect of group on age at baseline (F=24.0_1,295_, P < 0.01) and 12 months (F=18.1_1,295_, P < 0.01). There were no other significant differences for other demographic variables.

|  | Baseline |  | 12 months |  | 10 years |  |
| --- | --- | --- | --- | --- | --- | --- |
| diagnosis | CTRL | SCZ | CTRL | SCZ | CTRL | SCZ |
| n | 218 | 79 | 197 | 67 | 77 | 23 |
| Age mean [SD] | 33.9 [10.1]* | 27.8 [7.6]* | 35.1 [10.2]* | 29.3 [7.9]* | 40.7 [7.8] | 37.9 [7.3] |
| Sex ratio (female) | 42.6% | 41.7% | 40.6% | 41.8% | 37.6% | 34.7% |
| PANSS mean |  | 64 |  | 56 |  | 50 |
| [SD] |  | [14] |  | [15] |  | [16] |

### Normative modeling allows to compare across sites

Figure 1 displays the joint distributions of the median cortical thickness and their associated deviation scores for all healthy controls from the validation set of the adaptation (transfer) dataset. Of interest are the marginal densities: normative modeling of the median cortical thickness accounting for site and sex aligns the distributions and allows meaningful comparisons between samples from different sites in the deviation scores space. In contrast, estimates of cortical thickness appear highly impacted by the site effect (Figure 1). We also show an example of the deviation scores following the adaptation process in Supplementary Figure 8, which illustrates that the normative model does a good job in accounting for age-related variation. We have shown in prior work that normative modelling also allows meaningful comparisons across sex. ^40,50,55^

**Figure 1:**
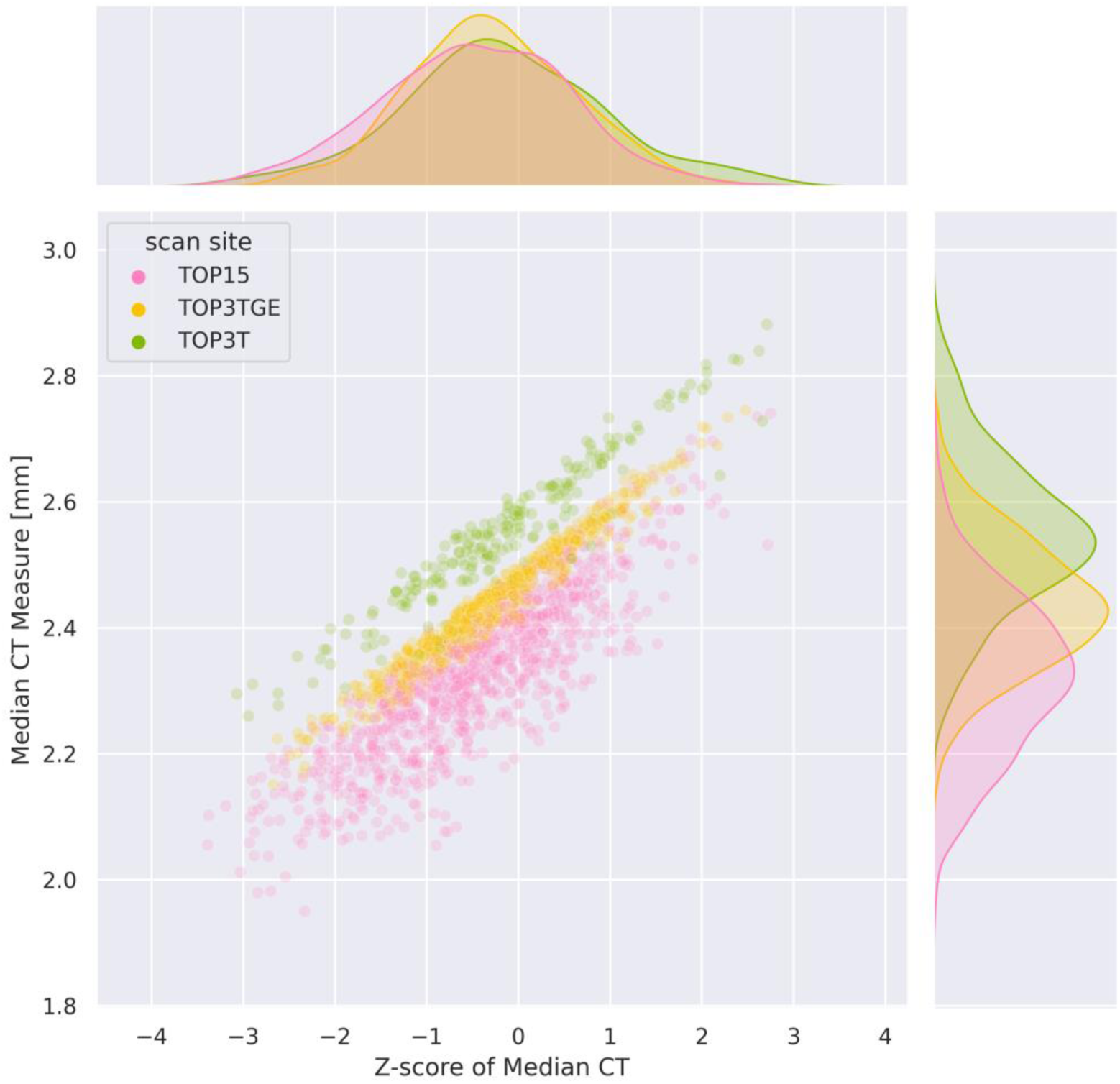
Joint-plot and marginal distributions of the median cortical thickness measures (y-axis) and associated deviation scores (x-axis), color coded by scanner, from the test samples of the adaptation set (cross-sectional TOP samples).

### Deviation scores difference by ROIs

LMEs revealed significant (p<.05, FDR corrected) conditional main effects of diagnosis and time x diagnosis interaction effects in a diffuse network of lateral temporal, parietal and frontal brain regions, and along the medial frontal and parietal lobes, bilaterally (Figure 2A-B). It should be noted that regression plots for each region (Supplementary Figure 4) showed a cross-over interaction in many (but not all) regions also having a conditional main effect of diagnosis. In such cases, the interaction effect should be considered the primary finding. Post hoc analyses (Supplementary Figures 3 and 4) showed that the interaction in most regions was principally due to more negative deviation scores in patients with SCZ at baseline, which attenuate over time such that a fewer number of significant regions were detected at the first follow up timepoint, and there were no significant differences observed at the final 10-year follow-up (Supplementary Figure 3-5).The effect sizes at the different timepoints are shown in Supplementary Figure 3D. Briefly, the Cohen’s *d* for median thickness deviation score at baseline is −0.46, −0.43 at 12-month follow-up and −0.27 at 10-year follow-up. Additionally, there were no significant differences in the deviation scores at baseline between patients who completed the 10-year follow-up scan (n=23) and those who did not but were enrolled at baseline in the 10-year study (n=40). In addition to these effects, we also detected conditional main effects for age, time since inclusion and sex (Supplementary Figure 6). However, since these are nuisance effects and all have very small effect size, we do not consider them further.

**Figure 2:**
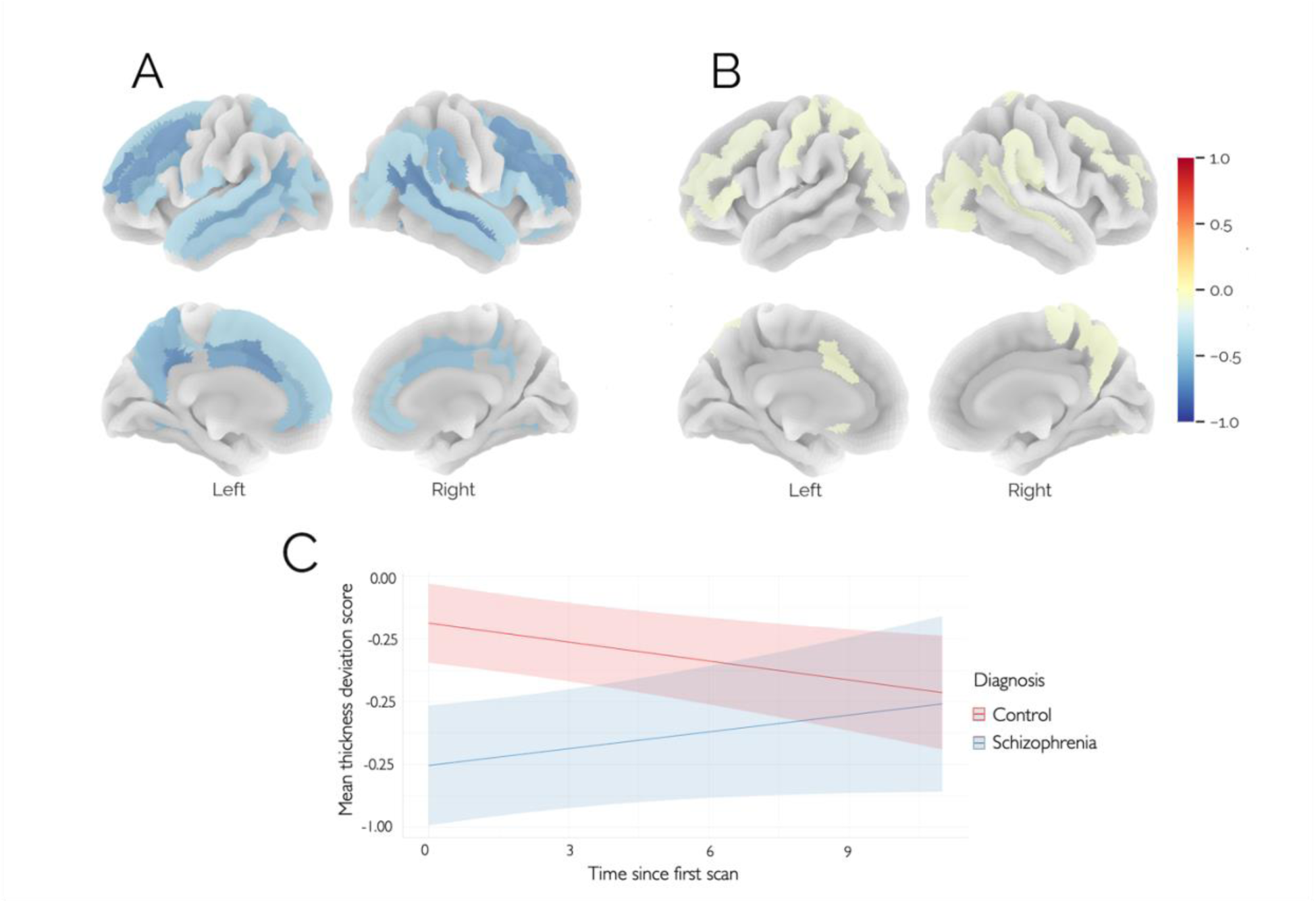
(A) Conditional main effect of diagnosis from the linear mixed model, showing that patients have reduced cortical thickness deviations overall (i.e. across all timepoints), color coded by effect size; (B) Time x diagnosis interaction effect, showing ROIs where the effect of diagnosis changes over time; (C) Regression plot for mean cortical thickness deviation across all ROIs, showing that the differences evident in the deviations at the first timepoint attenuate at later timepoints. The colour bar shows the effect size for each effect multiplied by the sign of the coefficient.

For visualization purposes, Figure 3 shows both the raw cortical thickness estimates and deviations from the normative model for mean cortical thickness in individuals with schizophrenia and healthy controls. As expected, the raw cortical thickness estimates (Figure 3A) are confounded by both ageing and scanner effects, showing the general reduction in cortical thickness that is expected over this lifespan stage.^40^ In contrast, the normative deviations are cleared from these effects (Figure 3B). In both cases the gradual attenuation of baseline reductions in cortical thickness over time is apparent.

**Figure 3:**
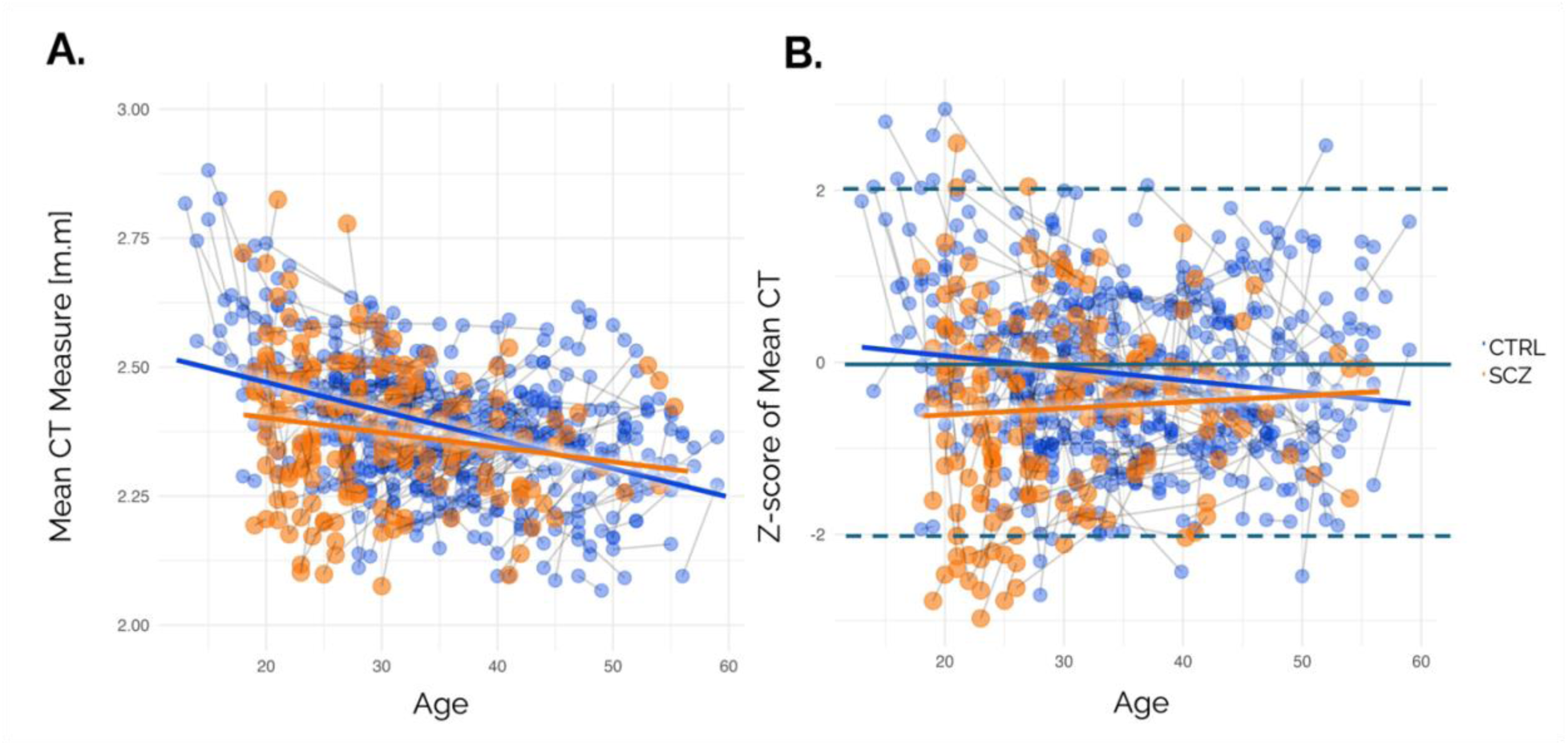
(A) Raw mean cortical thickness (CT) scores for individuals with schizophrenia (orange) and healthy controls (blue), where the line segments connect the successive time points of each individual. (B) Deviations from the normative model for mean cortical thickness.

### Number and distribution of regions with extreme deviations

Figure 4 and Suppl Table 3 summarize the distribution of ROIs showing significant higher proportion of extreme negative deviations among patients with schizophrenia compared to controls, at each timepoint. Here we find ROIs in both the left and right hemisphere with a significantly different overlap statistics of extreme negative deviation scores between patients and controls. The three ROIs with the strongest effects are the lateral aspect of the superior temporal gyrus (CV= 0.19, p < 0.01), and the opercular part of the interferer frontal gyrus (CV=0.18, p < 0.01) both in the left hemisphere, and the superior temporal sulcus (CV = 0.18, p < 0.01) in the right hemisphere. At the second and third time point none of the ROIs remained significant. A Mann-Whitney *U* test revealed no significant case-control differences in the number of positive extreme deviations, at any time points (Z>2). The analysis revealed significant case-control differences in the number of extreme negative deviations (Z<-2) at baseline (*p*=2.56*10^-5^,Common Language (CL) effect size=66%) and at the second assessment (*p*=.0006, CL effect size=63%).There was no significant case-control difference at the third time point (p = 1, CL effect size = 50%) (see eTables 4 and 5 in the supplement for further details).

**Figure 4:**
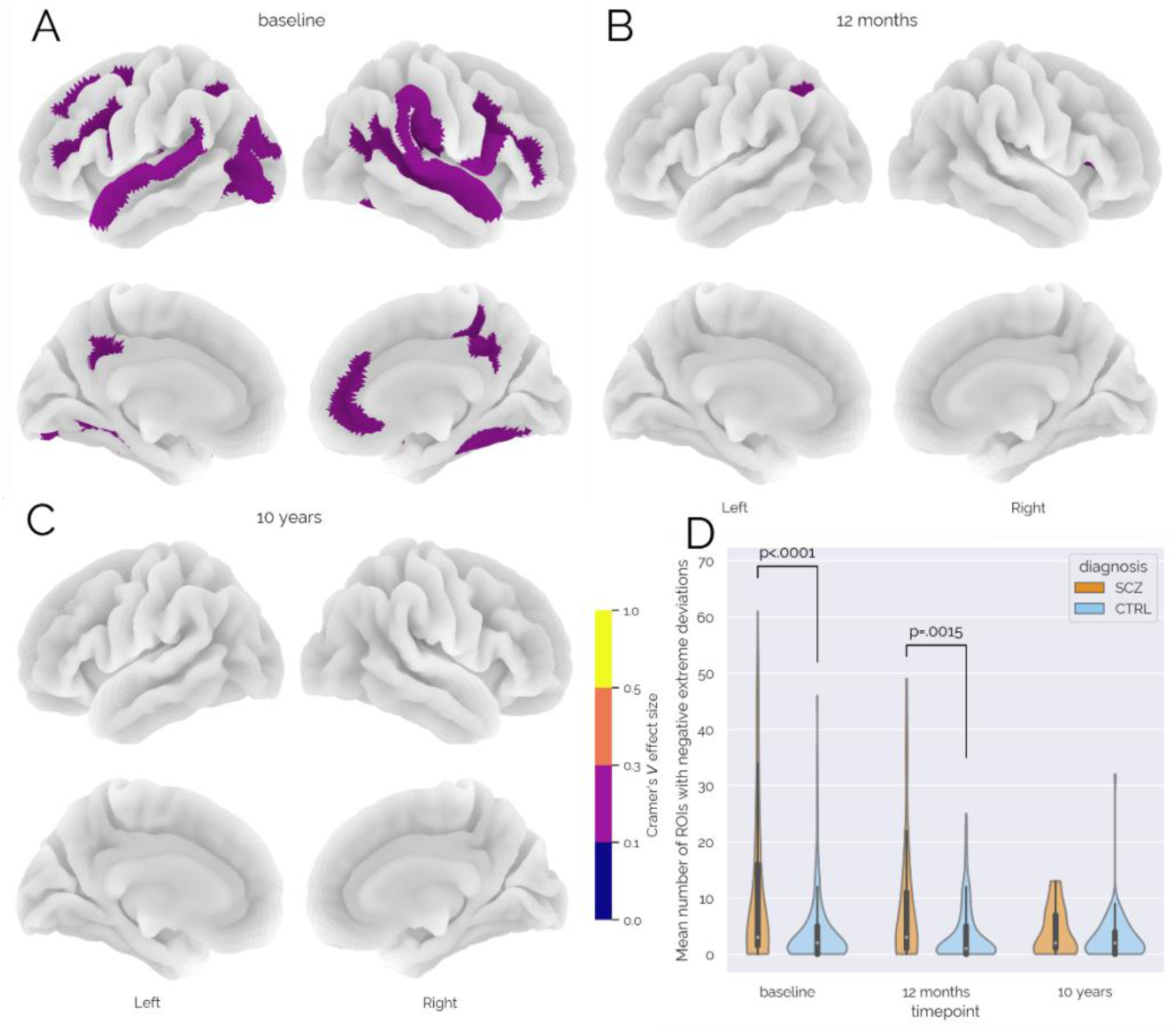
(A, B, C) ROIs showing a significantly higher proportion of negative extreme deviations among patients with schizophrenia (SCZ) compared to healthy controls (CTRL) at each timepoint. (D) *χ*^2^ test significant differences on negative extreme deviation distributions between people with SCZ and CTRL at each time point (see supplementary table S4 for a detailed summary for each ROI).

### Symptom scores from inclusion to 10-year follow-up

PANSS scores at each timepoint are shown in Figure 5.

**Figure 5:**
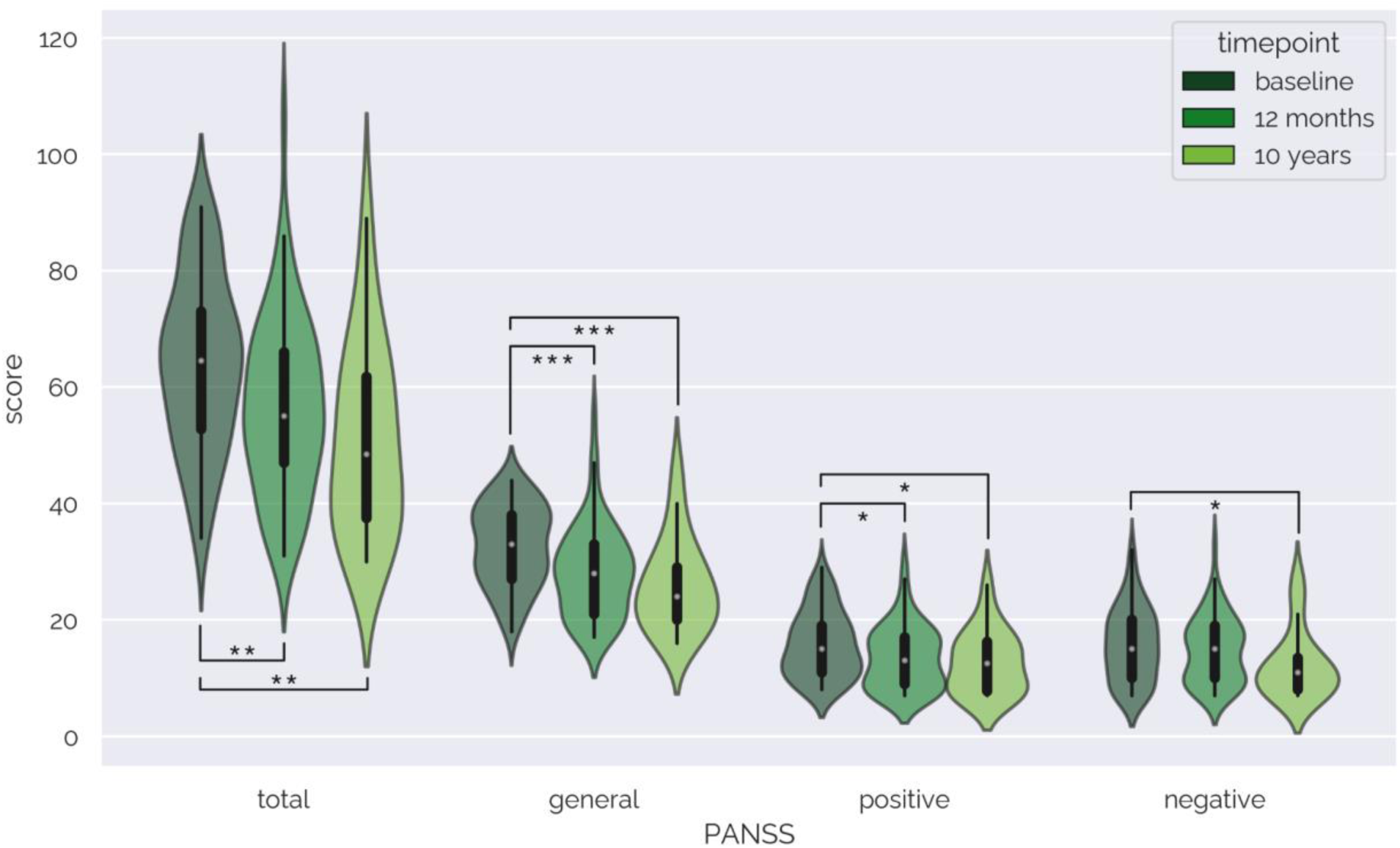
PANSS domain scores at the three timepoints. Most follow-up scores are significantly lower than baseline scores indicating a decrease in symptom severity over time (*: p<0.05, **: p<0.01, ***: p<0.001).

First, linear mixed models were fitted to examine the association between the different PANSS sub-scores and the predictor variables, including time since first scan, age at the first scan and sex. For all PANSS scores there was a significant effect of time (p< 0.001), indicating that PANSS scores decreased over time. None of the other covariates were significant (Supplementary Figure10). To further clarify the nature of these effects we also tested differences between individual timepoints, significant differences were observed only when comparing baseline values to the second follow-up scores, but not between the two follow-up time points, indicating that the main improvement took place during the first year (baseline – 12 month follow-up, *p*=.002 Cohen’s *d*=-.53; baseline – 10 year follow up *p*=.001, Cohen’s *d* = -.87). For PANSS subscales, we notice significantly lower scores for the general psychopathology scale at follow-ups compared to baseline (baseline – 12-month follow-up *p*=.0002 Cohen’s *d*=-.60; baseline – 10-year follow-up *p*=.0004 Cohen’s *d* = -.93). Symptom domains were also significantly reduced at 12-month and 10-year follow-ups compared to baseline for positive symptoms (baseline – 12-month follow-up *p*=.012 Cohen’s *d*=-.42; baseline – 10-year follow-up *p*=.026 Cohen’s *d* = -.56) and at 10 years for negative symptoms (baseline – 10-year follow-up *p*=.019 Cohen’s *d* = -.58).

### Association of cortical thickness deviation and PANSS scores

Figure 6 shows the results from the LME testing for associations between cortical deviations and PANSS. Several ROIs in the left hemisphere (LH) had a significant association with PANSS domain scores across time. Anterior cingulate gyrus was associated with PANSS total and PANSS general (coefficient=-4.0 and −1.79 respectively, p=.003 and .01). The left anterior segment of the circular sulcus of the insula (coefficient= −1.3, p=.04), the posterior ramus of the lateral sulcus (coefficient=1.52, p=.03), and the medial orbital sulcus (coefficient = −1.42, p=.02) were also significantly associated with PANSS negative. Negative associations indicate more negative deviation scores with higher symptom severity.

**Figure 6:**
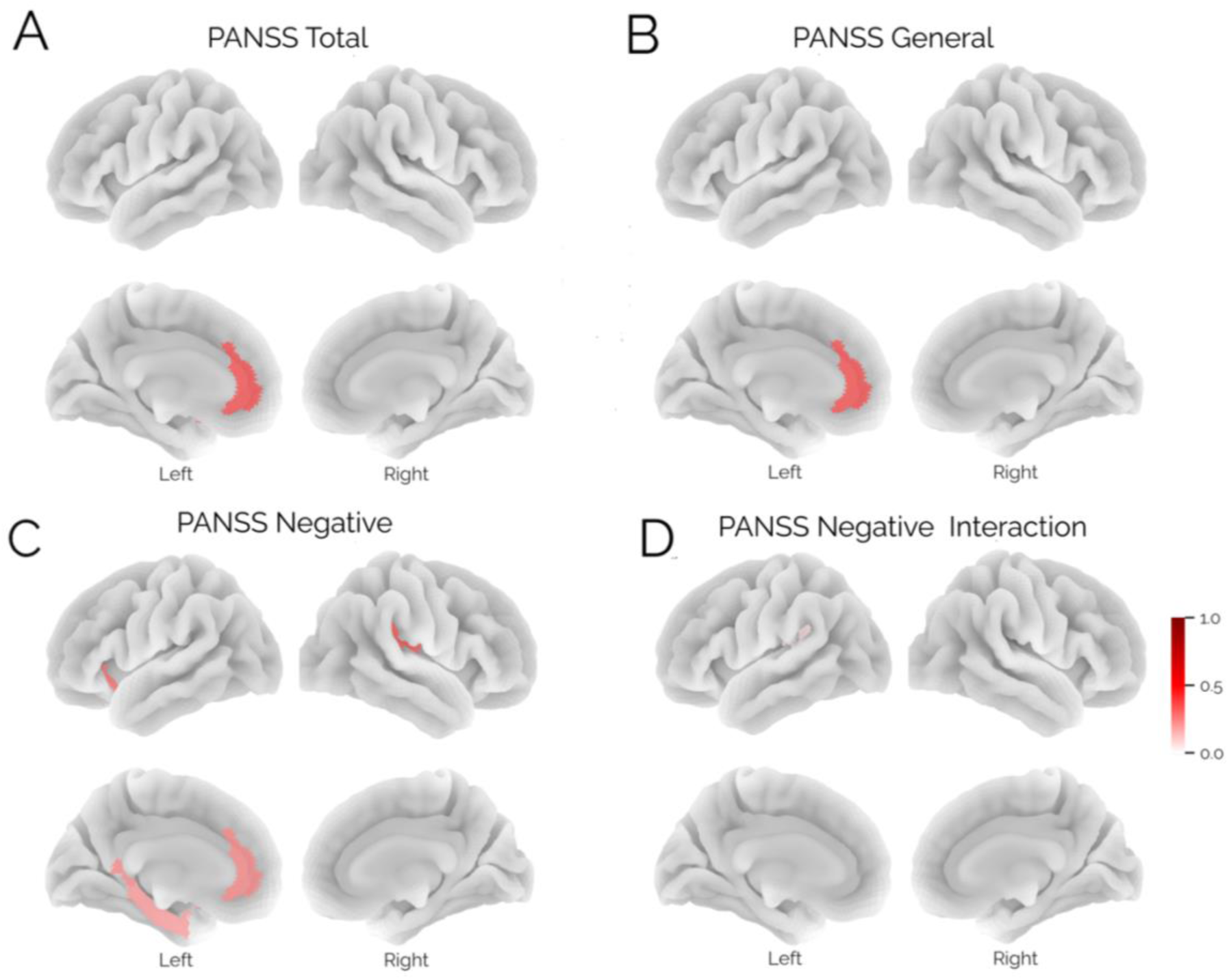
Results from the linear mixed effects model testing for associations between symptom scores and cortical deviations (equation 2). We report man effects for the PANSS total domain (A), the PANSS general domain (B), PANSS negative symptoms (C) and a significant interaction in a single region between the PANSS negative scores and follow up time (D). The colour bar shows the effects size of each effect.

There was a significant interaction between time since inclusion and the LH Posterior ramus (or segment) of the lateral sulcus on PANSS negative scores (Figure 5D, coefficient=0.28, p=.047), indicating that an initial association between larger negative deviations and more symptoms at baseline attenuated with time (see Supplementary Figure 6).

## DISCUSSION

In this study, we analyzed cortical thickness data from a 10-year longitudinal study of people with schizophrenia and healthy controls using structural brain MRI. The dataset included two or three time points for each participant, covering a period of approximately 10 years. We used normative modeling to investigate the deviations from an expected pattern of cortical thickness and how this pattern changed over time, both at the group level and at the individual level. We also examined the relationship between these deviation scores and the severity of psychotic symptoms as measured by PANSS. We report three main findings: (i) we show a diffuse pattern of cortical thickness atypicalities in schizophrenia early in the illness course, both in terms of the mean deviations across groups and in the number of extreme deviations at the individual level; and that these deviations (ii) attenuate over time; and (iii) associations with clinical symptoms across a distributed set of brain regions.

The pattern of negative deviation scores at baseline in a diffuse network of in patients compared to controls encompassed bilateral temporal, parietal and frontal regions. These significant effects are consistent with findings from large meta-analytic studies^22–24^ and more generally^14,28,62^, present both at baseline and at the 1st follow-up and are evident at the group and individual levels, both in terms of the mean and the number of extreme deviations (Figures 2 and 4, respectively). Significant interactions between diagnosis and time demonstrate that these differences attenuate over time. More specifically, between the first two time points, the number of ROIs with significant effects decreased, but also, the amplitude of the remaining effects. These findings are in line with previous reports that the gray matter differences were most severe in the early years after schizophrenia onset^36^, and are discordant with a general notion of schizophrenia as a neurodegenerative disorder with progressive brain aberrations over time^25,36^. The three previous long-term prospective brain imaging studies of first-episode psychosis also found that only a smaller subset of individuals with schizophrenia showed significant progressive brain changes and also had a high proportion of subjects using first generation antipsychotics^36,37,39^. In contrast, in our study, individuals with psychosis were almost exclusively treated with second generation antipsychotics. In view of the heterogeneity within the illness^28–30^ and since the proportion of patients with stable, poor clinical trajectories is relatively low, and the proportion with increasingly severe trajectories is even lower, we consider that the profile we detect likely reflects that very few participants with these trajectories are part of our study sample. Our findings thus primarily reflect the more common favourable trajectories. Since good-outcome first-episode patients leave the treatment services, more extensive cross-sectional studies based on clinical recruitment will include more multi-episode patients. We consider that cross-sectional studies will thus be enriched with patients with more severe trajectories, explaining the findings of a considerable heterogeneity found in these types of studies.^28,29,63^.

The pattern of brain regions showing case-control differences particularly implicated frontal and temporal regions including the paracentral lobule and sulcus, which have been associated with poor 1-year functional outcomes^64^ and the superior temporal gyrus, associated with positive symptoms.^62^ Several insula ROIs showed more negative deviations in participants with schizophrenia than in controls at both baseline and at the 1^st^ follow-up after on average 24 months. This region, especially on the left, is associated with inner speech and verbal hallucinations and reduced insula gray matter has been reported in hallucinators^65^.

We assessed the possibility of non-random attrition biasing our findings, which is often a concern in longitudinal studies and may influence the validity of regression models^66^. Logistic regressions and linear mixed effect models did not indicate any significant differences between attrition and cortical thickness deviation scores, PANSS scores, or in the frequency or duration of contacts with the health care system for mental, behavioral, and neurodevelopmental disorder-related events.

Linear mixed models revealed a few brain regions showing associations with symptoms. Most notably in the anterior cingulate gyrus where patients with more negative deviations exhibited higher symptom scores on multiple PANSS domains (total, positive and negative scales) and several other regions including the insula and parahippocampal gyrus showed an association with negative symptoms. Notably, alterations in the insula and the cingulate gyrus have been associated with negative symptoms, hallucinations and psychotic disorders^22,67,68^.

The observed case-control differences in cortical thickness deviation scores at baseline, reflecting a relatively early clinical phase, suggest that brain differences might be observable before the onset of the first episode. Indeed, large meta- of mega-analytic studies have shown that cortical alterations are present in the at-risk phase,^69^ although it has also been shown that such changes explain only a tiny proportion of the variance in regional deviations from a normative model for cortical thickness and do not predict conversion to psychosis.^70^ In individuals at clinical high risk for psychosis, multimodal (including brain MRI data) prediction of the negative symptom severity appears to yield promising results^71^. The notion of a neurodevelopmental component in the etiology of severe mental disorders is in line with previously reported correlations between deviation in cortical thickness and a general psychopathology score in a population-based sample of children and adolescents^72^. However we emphasize that our data cannot inform directly about the neurodevelopmental antecedents of schizophrenia since we lack information from important neurodevelopmental phases^41^.

### Limitations

Our study is subject to several limitations. First, even with access to the national registry data, ensuring representative recruitment in clinical and population-based cohorts is non-trivial, *e.g.* inclusion and exclusion criteria of patients^73,74^ or bias on the selection of healthy subjects^75^ or the retention of individuals with psychosis and controls. We are currently working on an extensive evaluation of these potential biases in normative models in separate work.

Second, whilst our findings are suggestive that second-generation antipsychotics may have different chronic effects on brain structure to first generation antipsychotics, we were unable to test this directly since only a very small number of particpants were taking first generation antipsychotics in our sample. Further work is therefore necessary to test this more directly.

Third, extreme cortical deviations may not only relate to schizophrenia-related pathologies but could also be markers of other effects, *e.g.* noise, artifacts, medications, co-morbidities, co-existing conditions, and various lifestyle and health-related behaviors or traits^76^. While we cannot rule out confounding effects, our quality control and validation procedures against clinical and registry data speak against this interpretation.

Finally, we acknowledge that our sample size at the third follow-up session is moderate, and this reduction in sample size could have biased our findings at later timepoints. However, we would like to emphasize that: (i) the proportion of subjects retained in our study compares favorably to the retention rates reported in the literature, particularly in view of the 10 year follow up period of this study and (ii) that the effect size estimates we present (supplementary figure 3 and 4) also speak against the possibility that the attenuation of effects we report is only attributable to a reduction in sample size. Also, despite best efforts, the inclusion of different scanners across different waves of the study may have influenced our findings. However, we have extensively validated the normative modelling framework we employ in such settings elsewhere and it shows good performance.^50^ Nevertheless, our findings should be considered preliminary at this stage and await replication in other cohorts.

## Conclusion

Using a unique dataset comprising clinical and MRI data from a 10-year longitudinal study with patients with schizophrenia we have shown an apparent gradual reduction in case-control cortical thickness deviations from the first psychotic episode to the 10-year follow-up assessment, with some evidence of regionally distributed associations with clinical symptoms over time. This study demonstrate that transfer learning from large scale reference normative models can be used to make meaningful comparisons of MRI features between participants across different scanners and provides preliminary evidence for cortical associations with longitudinal clinical outcome in people with schizophrenia.

## Data Availability

While the authors are open to collaborations using these data, data used in the present study can not be shared due to some participants not providing consent for data sharing.

## Acknowledgements

This study was supported by grant number ‘BRAINCHART’, 215698/Z/19/Z from the Wellcome Trust Innovator Award, the Research Council of Norway (#223273, #287714), the KG Jebsen Stiftelsen, the European Research Council under the European Union’s Horizon 2020 Research and Innovation program (ERC StG, grant 802998), and South-Eastern Norway Regional Health Authority (grants #2006233, #2006258, #2009037, #2011085, #2011096, #2012100, #2014102, #2015088, #2018093, #2019107, #2020086).

## REFERENCES

1. Insel, T. R. & Cuthbert, B. N. Endophenotypes: Bridging Genomic Complexity and Disorder Heterogeneity. Biological Psychiatry 66, 988–989 (2009).

2. Alnæs, D. et al. Brain Heterogeneity in Schizophrenia and Its Association With Polygenic Risk. JAMA Psychiatry 76, 739 (2019).

3. Kaufmann, T. et al. Common brain disorders are associated with heritable patterns of apparent aging of the brain. Nat Neurosci 22, 1617–1623 (2019).

4. Barr, P. B., Bigdeli, T. B. & Meyers, J. L. Prevalence, Comorbidity, and Sociodemographic Correlates of Psychiatric Diagnoses Reported in the All of Us Research Program. JAMA Psychiatry 79, 622 (2022).

5. Christensen, M. K. et al. The cost of mental disorders: a systematic review. Epidemiol Psychiatr Sci 29, e161 (2020).

6. Plana-Ripoll, O., et al. Nature and prevalence of combinations of mental disorders and their association with excess mortality in a population-based cohort study. World Psychiatry 19, 339–349 (2020).

7. Solmi, M. et al. Age at onset of mental disorders worldwide: large-scale meta-analysis of 192 epidemiological studies. Mol Psychiatry 27, 281–295 (2022).

8. Lally, J. et al. Remission and recovery from first-episode psychosis in adults: systematic review and meta-analysis of long-term outcome studies. Br J Psychiatry 211, 350–358 (2017).

9. Friis, S. et al. Early Predictors of Ten-Year Course in First-Episode Psychosis. PS 67, 438–443 (2016).

10. Austin, S. F. et al. Long-term trajectories of positive and negative symptoms in first episode psychosis: A 10year follow-up study in the OPUS cohort. Schizophrenia Research 168, 84–91 (2015).

11. O’Keeffe, D. et al. The iHOPE-20 study: Relationships between and prospective predictors of remission, clinical recovery, personal recovery and resilience 20 years on from a first episode psychosis. Aust N Z J Psychiatry 53, 1080–1092 (2019).

12. Morgan, C. et al. Rethinking the course of psychotic disorders: modelling long-term symptom trajectories. Psychol. Med. 52, 2641–2650 (2022).

13. Patel, P. K., Leathem, L. D., Currin, D. L. & Karlsgodt, K. H. Adolescent Neurodevelopment and Vulnerability to Psychosis. Biological Psychiatry 89, 184–193 (2021).

14. Van Haren, N. E. M. Changes in Cortical Thickness During the Course of Illness in Schizophrenia. Arch Gen Psychiatry 68, 871 (2011).

15. Barth, C. et al. Trajectories of brain volume change over 13 years in chronic schizophrenia. Schizophrenia Research 222, 525–527 (2020).

16. Haukvik, U. K. et al. No progressive brain changes during a 1-year follow-up of patients with first-episode psychosis. Psychol. Med. 46, 589–598 (2016).

17. Jørgensen, K. N. et al. First- and second-generation antipsychotic drug treatment and subcortical brain morphology in schizophrenia. Eur Arch Psychiatry Clin Neurosci 266, 451–460 (2016).

18. Ansell, B. R. E. et al. Divergent effects of first-generation and second-generation antipsychotics on cortical thickness in first-episode psychosis. Psychol. Med. 45, 515–527 (2015).

19. Wiegand, L. C. et al. Prefrontal cortical thickness in first-episode psychosis: a magnetic resonance imaging study. Biological Psychiatry 55, 131–140 (2004).

20. Nelson, E. A. et al. A Prospective Longitudinal Investigation of Cortical Thickness and Gyrification in Schizophrenia. Can J Psychiatry 65, 381–391 (2020).

21. Zhang, W. et al. Discrete patterns of cortical thickness in youth with bipolar disorder differentially predict treatment response to quetiapine but not lithium. Neuropsychopharmacol 43, 2256–2263 (2018).

22. Van Erp, T. G. M. et al. Cortical Brain Abnormalities in 4474 Individuals With Schizophrenia and 5098 Control Subjects via the Enhancing Neuro Imaging Genetics Through Meta Analysis (ENIGMA) Consortium. Biological Psychiatry 84, 644–654 (2018).

23. Cannon, T. D. et al. Progressive Reduction in Cortical Thickness as Psychosis Develops: A Multisite Longitudinal Neuroimaging Study of Youth at Elevated Clinical Risk. Biological Psychiatry 77, 147–157 (2015).

24. for the ENIGMA Schizophrenia Working Group et al. Subcortical brain volume abnormalities in 2028 individuals with schizophrenia and 2540 healthy controls via the ENIGMA consortium. Mol Psychiatry 21, 547–553 (2016).

25. Kochunov, P. & Hong, L. E. Neurodevelopmental and neurodegenerative models of schizophrenia: white matter at the center stage. Schizophr Bull 40, 721–728 (2014).

26. Andreasen, N. C. The lifetime trajectory of schizophrenia and the concept of neurodevelopment. Dialogues Clin Neurosci 12, 409–415 (2010).

27. Insel, T. R. Mental disorders in childhood: shifting the focus from behavioral symptoms to neurodevelopmental trajectories. JAMA 311, 1727–1728 (2014).

28. Wolfers, T. et al. Mapping the Heterogeneous Phenotype of Schizophrenia and Bipolar Disorder Using Normative Models. JAMA Psychiatry 75, 1146–1155 (2018).

29. Wolfers, T. et al. Replicating extensive brain structural heterogeneity in individuals with schizophrenia and bipolar disorder. Hum Brain Mapp 42, 2546–2555 (2021).

30. Clementz, B. A. et al. Identification of Distinct Psychosis Biotypes Using Brain-Based Biomarkers. Am J Psychiatry 173, 373–384 (2016).

31. Cuthbert, B. N. & Insel, T. R. Toward the future of psychiatric diagnosis: the seven pillars of RDoC. BMC Med 11, 126 (2013).

32. Farley, J. D. PHYLOGENETIC ADAPTATIONS AND THE GENETICS OF PSYCHOSIS. Acta Psychiatr Scand 53, 173–192 (1976).

33. Insel, T. R. Rethinking schizophrenia. Nature 468, 187–193 (2010).

34. Insel, T. R. & Cuthbert, B. N. Brain disorders? Precisely. Science 348, 499–500 (2015).

35. Rubio, J. M., Malhotra, A. K. & Kane, J. M. Towards a framework to develop neuroimaging biomarkers of relapse in schizophrenia. Behavioural Brain Research 402, 113099 (2021).

36. Andreasen, N. C. et al. Progressive Brain Change in Schizophrenia: A Prospective Longitudinal Study of First-Episode Schizophrenia. Biological Psychiatry 70, 672–679 (2011).

37. Huhtaniska, S. et al. Long-term antipsychotic and benzodiazepine use and brain volume changes in schizophrenia: The Northern Finland Birth Cohort 1966 study. Psychiatry Research: Neuroimaging 266, 73–82 (2017).

38. Vita, A., De Peri, L., Deste, G., Barlati, S. & Sacchetti, E. The Effect of Antipsychotic Treatment on Cortical Gray Matter Changes in Schizophrenia: Does the Class Matter? A Meta-analysis and Meta-regression of Longitudinal Magnetic Resonance Imaging Studies. Biological Psychiatry 78, 403–412 (2015).

39. Canal-Rivero, M. et al. Longitudinal trajectories in negative symptoms and changes in brain cortical thickness: 10-year follow-up study. Br J Psychiatry 223, 309–318 (2023).

40. Rutherford, S. et al. Charting brain growth and aging at high spatial precision. eLife 11, e72904 (2022).

41. Bethlehem, R. a. I., et al. Brain charts for the human lifespan. Nature 604, 525–533 (2022).

42. Antoniades, M. et al. Personalized Estimates of Brain Structural Variability in Individuals With Early Psychosis. Schizophrenia Bulletin 47, 1029–1038 (2021).

43. Cole, T. J. The development of growth references and growth charts. Ann Hum Biol 39, 382–394 (2012).

44. Marquand, A. F. et al. Conceptualizing mental disorders as deviations from normative functioning. Mol Psychiatry 24, 1415–1424 (2019).

45. Marquand, A. F., Rezek, I., Buitelaar, J. & Beckmann, C. F. Understanding Heterogeneity in Clinical Cohorts Using Normative Models: Beyond Case-Control Studies. Biol Psychiatry 80, 552–561 (2016).

46. Rutherford, S. et al. Evidence for embracing normative modeling. Elife 12, e85082 (2023).

47. Fraza, C. J., Dinga, R., Beckmann, C. F. & Marquand, A. F. Warped Bayesian linear regression for normative modelling of big data. Neuroimage 245, 118715 (2021).

48. Marquand, A. F., Wolfers, T., Mennes, M., Buitelaar, J. & Beckmann, C. F. Beyond Lumping and Splitting: A Review of Computational Approaches for Stratifying Psychiatric Disorders. Biological Psychiatry: Cognitive Neuroscience and Neuroimaging 1, 433–447 (2016).

49. Zabihi, M. et al. Dissecting the Heterogeneous Cortical Anatomy of Autism Spectrum Disorder Using Normative Models. Biol Psychiatry Cogn Neurosci Neuroimaging 4, 567–578 (2019).

50. Gaiser, C. et al. Estimating cortical thickness trajectories in children across different scanners using transfer learning from normative models. Human Brain Mapping 45, e26565 (2024).

51. Kay, S. R., Fiszbein, A. & Opler, L. A. The Positive and Negative Syndrome Scale (PANSS) for Schizophrenia. Schizophrenia Bulletin 13, 261–276 (1987).

52. Destrieux, C., Fischl, B., Dale, A. & Halgren, E. Automatic parcellation of human cortical gyri and sulci using standard anatomical nomenclature. Neuroimage 53, 1–15 (2010).

53. Rosen, A. F. G. et al. Quantitative assessment of structural image quality. Neuroimage 169, 407–418 (2018).

54. Kia, S. M. et al. Hierarchical Bayesian Regression for Multi-Site Normative Modeling of Neuroimaging Data. arXiv:2005.12055 [cs, stat] (2020).

55. Kia, S. M. et al. Closing the life-cycle of normative modeling using federated hierarchical Bayesian regression. PLoS One 17, e0278776 (2022).

56. Ducharme, S. et al. Trajectories of cortical thickness maturation in normal brain development — The importance of quality control procedures. NeuroImage 125, 267–279 (2016).

57. Monereo-Sánchez, J. et al. Quality control strategies for brain MRI segmentation and parcellation: Practical approaches and recommendations - insights from the Maastricht study. NeuroImage 237, 118174 (2021).

58. Rutherford, S. et al. The normative modeling framework for computational psychiatry. Nat Protoc 17, 1711–1734 (2022).

59. Bayer, J. M. M. et al. Accommodating site variation in neuroimaging data using normative and hierarchical Bayesian models. Neuroimage 264, 119699 (2022).

60. Benjamini, Y. & Hochberg, Y. Controlling the False Discovery Rate: A Practical and Powerful Approach to Multiple Testing. Journal of the Royal Statistical Society: Series B (Methodological) 57, 289–300 (1995).

61. Wolfers, T. et al. Individual differences v. the average patient: mapping the heterogeneity in ADHD using normative models. Psychol Med 50, 314–323 (2020).

62. Walton, E. et al. Positive symptoms associate with cortical thinning in the superior temporal gyrus via the ENIGMA Schizophrenia consortium. Acta Psychiatr Scand 135, 439–447 (2017).

63. Lv, J. et al. Individual deviations from normative models of brain structure in a large cross-sectional schizophrenia cohort. Mol Psychiatry 26, 3512–3523 (2021).

64. Sasabayashi, D. et al. Reduced cortical thickness of the paracentral lobule in at-risk mental state individuals with poor 1-year functional outcomes. Transl Psychiatry 11, 396 (2021).

65. Barber, L., Reniers, R. & Upthegrove, R. A review of functional and structural neuroimaging studies to investigate the inner speech model of auditory verbal hallucinations in schizophrenia. Transl Psychiatry 11, 582 (2021).

66. Wolke, D. et al. Selective drop-out in longitudinal studies and non-biased prediction of behaviour disorders. Br J Psychiatry 195, 249–256 (2009).

67. Pantelis, C. et al. Neuroanatomical abnormalities before and after onset of psychosis: a cross-sectional and longitudinal MRI comparison. Lancet 361, 281–288 (2003).

68. Wylie, K. P. & Tregellas, J. R. The role of the insula in schizophrenia. Schizophrenia Research 123, 93–104 (2010).

69. ENIGMA Clinical High Risk for Psychosis Working Group et al. Association of Structural Magnetic Resonance Imaging Measures With Psychosis Onset in Individuals at Clinical High Risk for Developing Psychosis: An ENIGMA Working Group Mega-analysis. JAMA Psychiatry 78, 753 (2021).

70. Haas, S. S., et al. Normative Modeling of Brain Morphometry in Clinical High-Risk for Psychosis. http://biorxiv.org/lookup/doi/10.1101/2023.01.17.523348 (2023) doi:10.1101/2023.01.17.523348.

71. Hauke, D. J. et al. Multimodal prognosis of negative symptom severity in individuals at increased risk of developing psychosis. Transl Psychiatry 11, 312 (2021).

72. Kjelkenes, R. et al. Deviations from normative brain white and gray matter structure are associated with psychopathology in youth. Developmental Cognitive Neuroscience 58, 101173 (2022).

73. Murray, G. K. et al. Could Polygenic Risk Scores Be Useful in Psychiatry?: A Review. JAMA Psychiatry 78, 210 (2021).

74. Taipale, H. et al. Representation and Outcomes of Individuals With Schizophrenia Seen in Everyday Practice Who Are Ineligible for Randomized Clinical Trials. JAMA Psychiatry 79, 210 (2022).

75. Fry, A. et al. Comparison of Sociodemographic and Health-Related Characteristics of UK Biobank Participants With Those of the General Population. American Journal of Epidemiology 186, 1026–1034 (2017).

76. Elad, D. et al. Improving the predictive potential of diffusion MRI in schizophrenia using normative models—Towards subject-level classification. Human Brain Mapping 42, 4658–4670 (2021).

