## Supplementary material for "A 10-Year Longitudinal Study of Brain Cortical Thickness in People with First-Episode Psychosis using Normative Models"

Supplementary Materials

**Supplementary information (Methods / Results / Future Directions)**

**Supplementary Figure 1**: Count plots for SCZ and CTRL at different timepoints, color-coded by diagnosis.

**Supplementary Figure 2**: Sankey diagram of the MRI sessions completed by patients with schizophrenia.

**Supplementary Figure 3:** Significant differences in deviation scores between healthy individuals and people with schizophrenia at baseline (A), 12-month follow-up (B) and 10-year follow-up (C), color coded by effect size (Cohen's *d*), (D) empirical cumulative distribution function plot of the effect size for the three time points.

**Supplementary Figure 4**: Unthresholded differences in deviation scores between healthy individuals and patients with schizophrenia at baseline, 12-month follow-up (B) and 10-year follow-up (C), color coded by effect size (Cohen's *d*).

**Supplementary Figure 5**: Interaction effects between time and diagnosis group for each significant interaction effect for the left hemisphere (A) and the right hemisphere (B).

**Supplementary Figure 6**: Conditional main effects for age, time since inclusion (B) and sex (C) from the linear mixed model shown in Figure 2 in the main manuscript.

**Supplementary Figure 7**: Significant interaction effects (FDR corrected) between time points and deviation score of the posterior ramus of the lateral sulcus in the left hemisphere.

**Supplementary Figure 8**: The deviation in mean cortical thickness across timepoints for the healthy controls. This shows that the normative model removes age related effects such that the trajectory of each healthy participant is flat and with the distribution centred at zero.

**Supplementary Figure 9:** age of each participant at each measurement timepoint for the healthy controls (A) and patients with schizophrenia (B).

**Supplementary Figure 10:** Results from the LME models showing the estimates for time since inclusion (delay), age at baseline and sex.

**Supplementary Table 1**: adaptation samples (CTRL only) for the transfer of normative models to the three Oslo's scanners used in the longitudinal study.

**Supplementary Table 2**: ROIs with Significantly different deviation scores between patients with schizophrenia and healthy individuals, by time point.

**Supplementary Table 3**: ROIs with Significantly different overlap statistics of extreme negative deviation scores between patients with schizophrenia and healthy individuals.

**Supplementary Table 4**: Percentage (%) of extreme positive deviation overlap by ROI, by time point, for controls and patients with schizophrenia.

**Supplementary Table 5:** Percentage (%) of extreme negative deviation overlap by ROI, by time point, for controls and patients with schizophrenia.

**Supplementary References**

### Supplementary Methods

#### MRI scanners

The participants were scanned on either a 1.5T Siemens Magnetom Sonata scanner (baseline=76% of all participants at that time point, 12-months=72%, 10-years=6%, Siemens Medical Solutions, Erlangen, Germany, TOP15), a 3T General Electric Signa HDxt scanner (baseline=5%, 12-months=4%, 10-years=0%, GE Healthcare, Milwaukee, WI, USA, TOP3T), or a 3T General Electric 3T Discovery GE750 (baseline=19%, 12-months=24%, 10-years=94%, TOP3TGE).

#### Participants

Patients (aged 18-53 years) and healthy controls (aged 13-56 years) understood and spoke a Scandinavian language, had no history of severe head trauma, and had an IQ above 70. Patients were assessed by trained physicians or clinical psychologists. In this study, the first-episode was defined as having had the first adequate treatment for a psychosis episode within the last 12 months

Patients were screened for mental illnesses before participation and were excluded from the study if they, or if any first-degree relatives, presented a history of severe mental illness or had a history of severe substance or alcohol abuse. The TOP study was approved by the Norwegian Regional Committee for Medical Research Ethics and the Norwegian Data Protection Authority, and conducted in accordance with the Helsinki declaration. All participants were informed about the study before participating and provided written informed consent.

Patients were not considered to be first-episode patients if they previously, on any occasion before the starting point of the index treatment, had been treated with antipsychotic medication for more than 12 weeks (or shorter if symptomatic remission was achieved before 12 weeks).

Among the participants who were eligible for the 10-year follow-up, some did not reply, could not be located, or decided not to complete part or all of the 10-year interviews or MRI scan, and some missed the 12-month follow-up but completed the 10-year follow-up.

#### Normative Models

The models were trained using the Destrieux et al. (2010) atlas [1]. A linear regression model also pointed to a gradual cortical thinning in the test participant age range [18-59][2–4] and thus validated the linear regression method used in the hierarchical Bayesian regression models [5]. In publicly available datasets used for estimating the reference normative models, we included only data from the first visit when multiple visits were available.

#### PANSS scores

Whenever possible the PANSS scores were computed using data from all patients available for each time points.

#### Norwegian National Registry

All participants recruited to the TOP study consented to linkage of relevant healthcare information from registry data to track individual disease trajectories across different treatment facilities in Norway[6]. In the current study we used data from the National Patient Registry (NPR) to track contact with the healthcare system after first inclusion. From NPR, we extracted data including level of care (inpatient/outpatient data), type of care (somatic hospital, mental health care facilities, substance use treatment facility or psychiatrists/psychologists with governmental reimbursement) and assigned ICD-10 diagnoses (for mental, behavioral, and neurodevelopmental disorders, F01–F99), and all contact (number and duration of visits) related to a schizophrenia-like psychotic disorder (F20–F29) were obtained from the period (15.08.05 – 31.12.20).

#### Statistical tests

We compared the deviations from patients to controls at the different time points with two-sided t-tests. We used $\chi^2$ tests on the distributions of extreme deviation between patients and controls at the different time points, and on the sex distribution at each time point. We compared PANSS scores from all available patients at each time point with the non-parametric Mann-Whitney U test. We also applied this test to assess possible differences between patients who completed the 10-year longitudinal study and those who dropped out, based on their history of contacts with the healthcare system (count and duration, available for all patients, over the 10-year span).

### Results

**Attrition**

The number of longitudinal patients retained between the 12 month and 10-year follow-ups is n=23, (38.3%) respectively, for controls the retention rate is 90% (n=197) after 12 months, and 35% (n=77) after 10 years. $\chi^2$ tests on sex distribution at each time point, for each diagnosis, did not return any significant differences. Neither a Mann-Whitney U test returned any significant differences in PANSS scores at baseline or 12-month follow-up, nor did a t-test on the deviation scores at these time points, between patients who eventually dropped out and those who were retained. An additional Mann-Whitney U test did not find any significant differences between the two subpopulations in neither the number, nor the duration, of contacts with the healthcare system for ICD-10 classified "Mental, Behavioral and Neurodevelopmental disorders" reason. Furthermore, we used a GLMM with binomial family to learn the relation between PANSS scores and deviation scores at the first two time points with the attrition/dropout status of the patients with schizophrenia at the 10-year follow-up. There were no significant effects reported by the model.

**Medication**

At the baseline assessment, 62% (49 out of 79) of the patients were taking second-generation antipsychotic medications, while the remaining 38% (30) were not taking any antipsychotic medications. At 12-months follow-up, 48% (26 out of 54) of the patients were taking second-generation antipsychotic medications, while only 2% (1) were taking first-generation antipsychotic medications. The remaining 50% (27) did not report taking any antipsychotic medications. At the 10-year follow-up, 17% (2 out of 12) of the patients were taking second-generation antipsychotic medications, while the remaining 83% (10) were not taking any medications.

### Future directions / Developments:

Several continuations naturally follow this study. The application of normative modeling and brain imaging in childhood and adolescent samples provides an opportunity to link relevant genetic, clinical and environmental risk factors to neurodevelopmental processes[11,12]. The use of multimodal deviation scores (surface and volume, DTI, fMRI, rsfMRI) and of other dimensions (genetics, phenotypes, clinical scores, cortical asymmetry) represents an additional extension.

Another consideration would be to look at differences in patients fulfilling some remission criteria and those who do not[13]. It could be interesting to see if the delay of transitioning to a psychosis in at clinical high-risk individuals can be related to any deviations from the normative models[14]. A further development from this study would be to determine the impact of medication depending on the type and defined daily dose[15,16]. Finally, how would the deviation scores of these patients look in a 20-year follow-up? How did they look a few months before the acute phase?

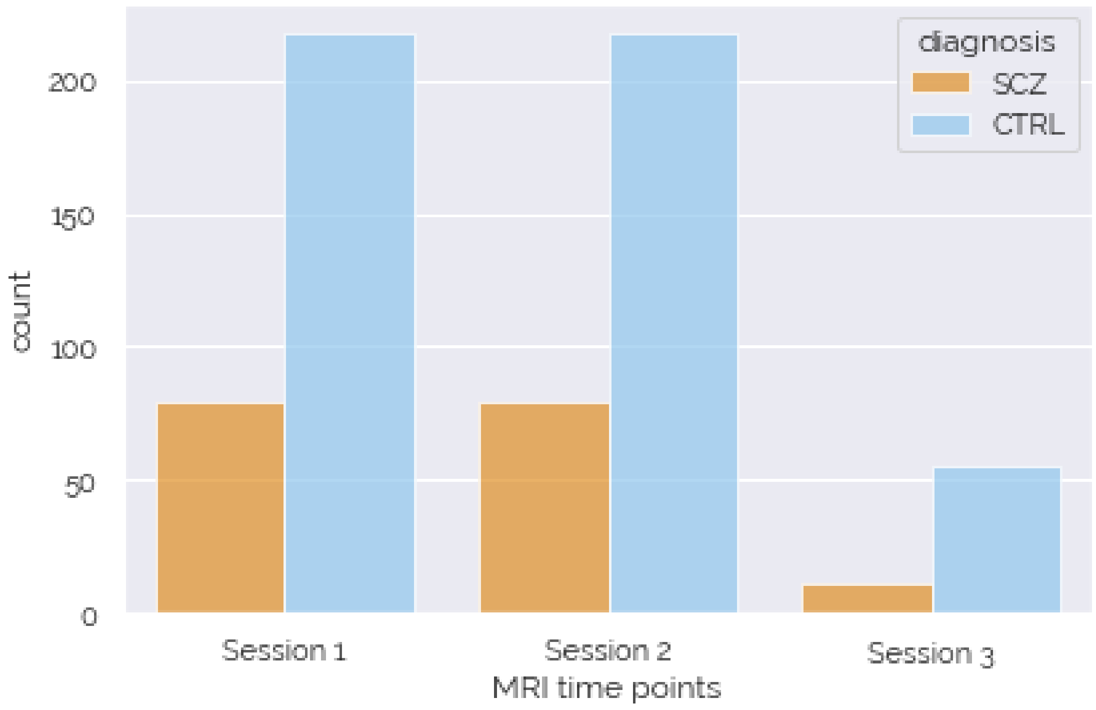


**Supplementary Figure 1**: Count plots for SCZ and CTRL at different timepoints, color-coded by diagnosis.

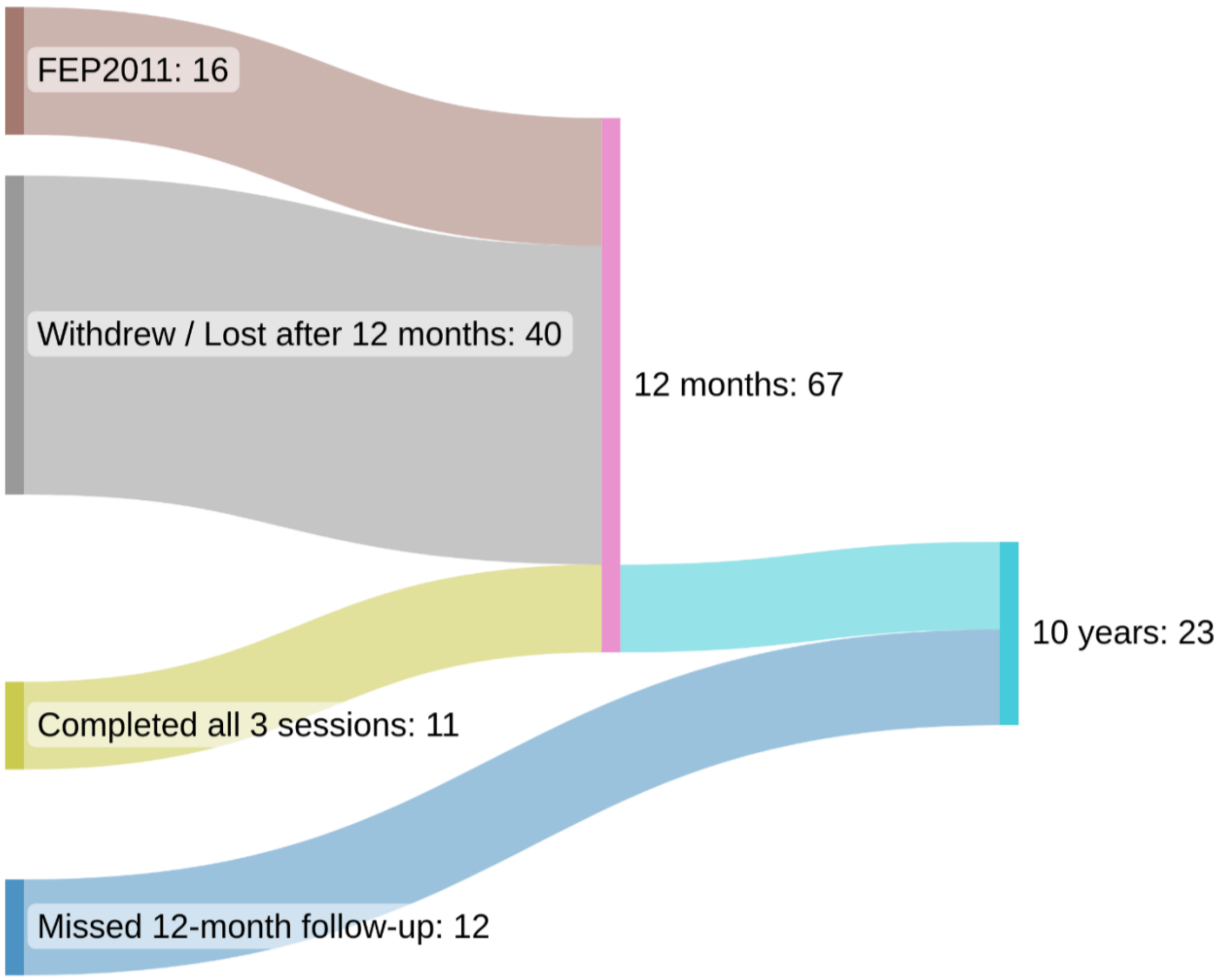


**Suppelementary Figure 2**: Sankey diagram of the MRI sessions completed by patients with schizophrenia.

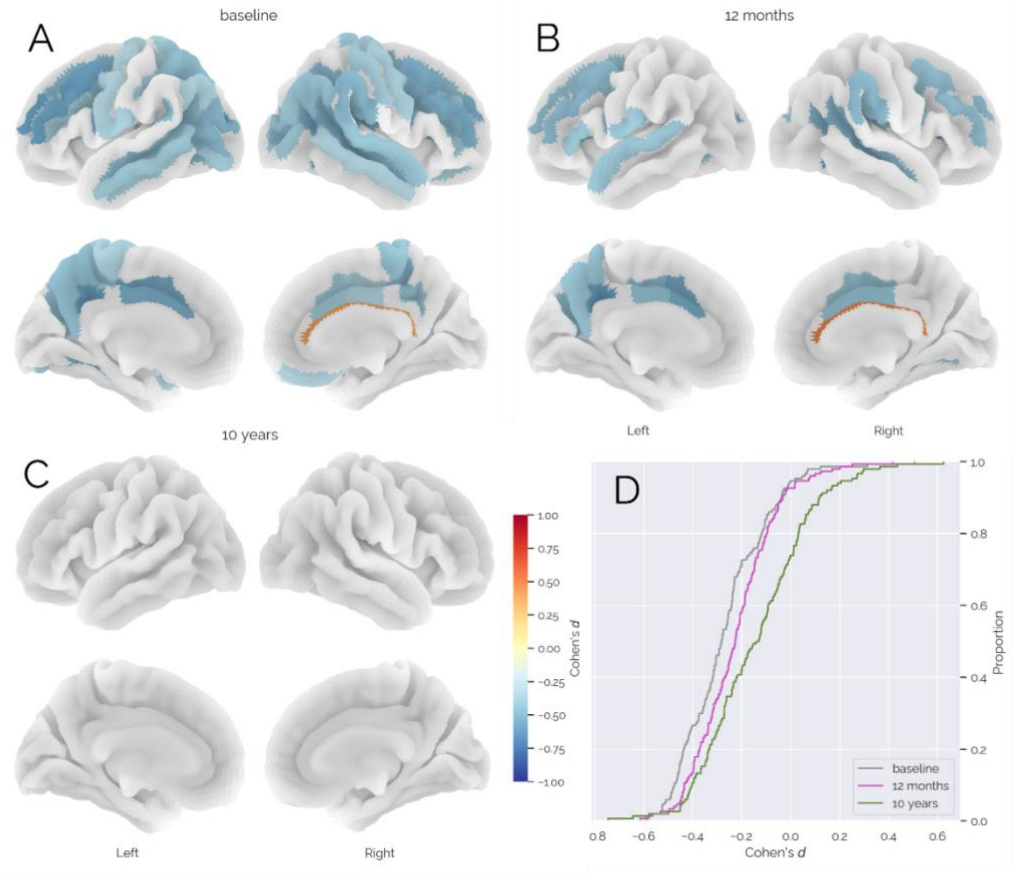


**Supplementary Figure 3:** Significant differences in deviation scores between healthy individuals and people with schizophrenia at baseline (A), 2-year follow-up (B) and 10-year follow-up (C), color coded by effect size (Cohen's *d*), (D) empirical cumulative distribution function plot of the effect size for the three time points.

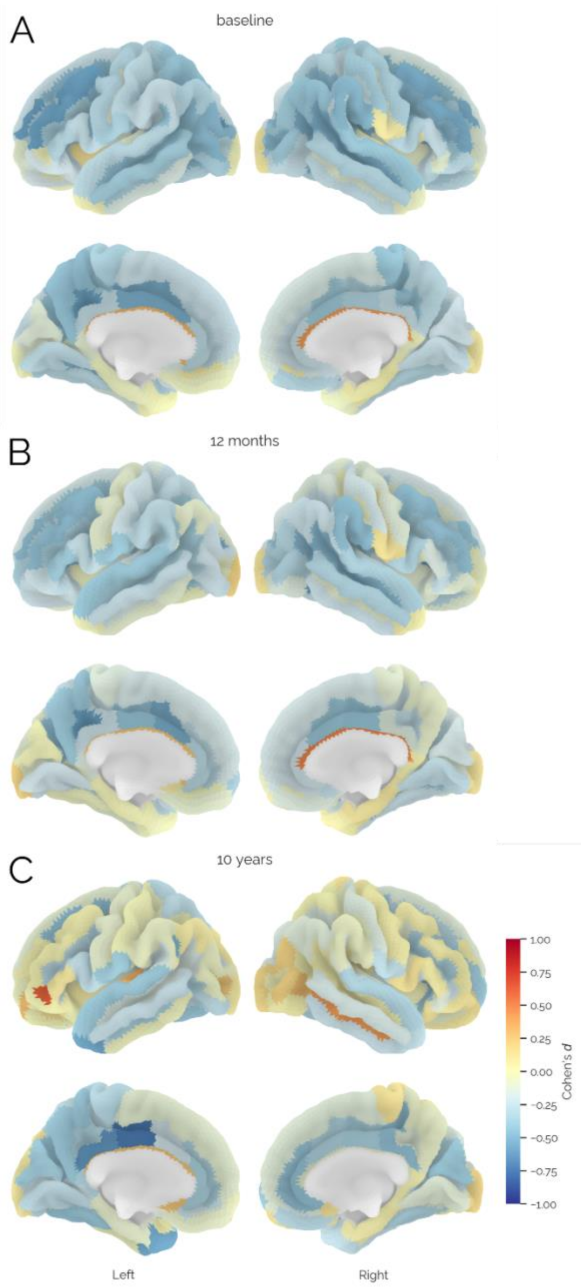


**Supplementary Figure 4**: Unthresholded differences in deviation scores between healthy individuals and patients with schizophrenia at baseline (A), Session two (B) and session three(C), color coded by effect size (Cohen's *d*).

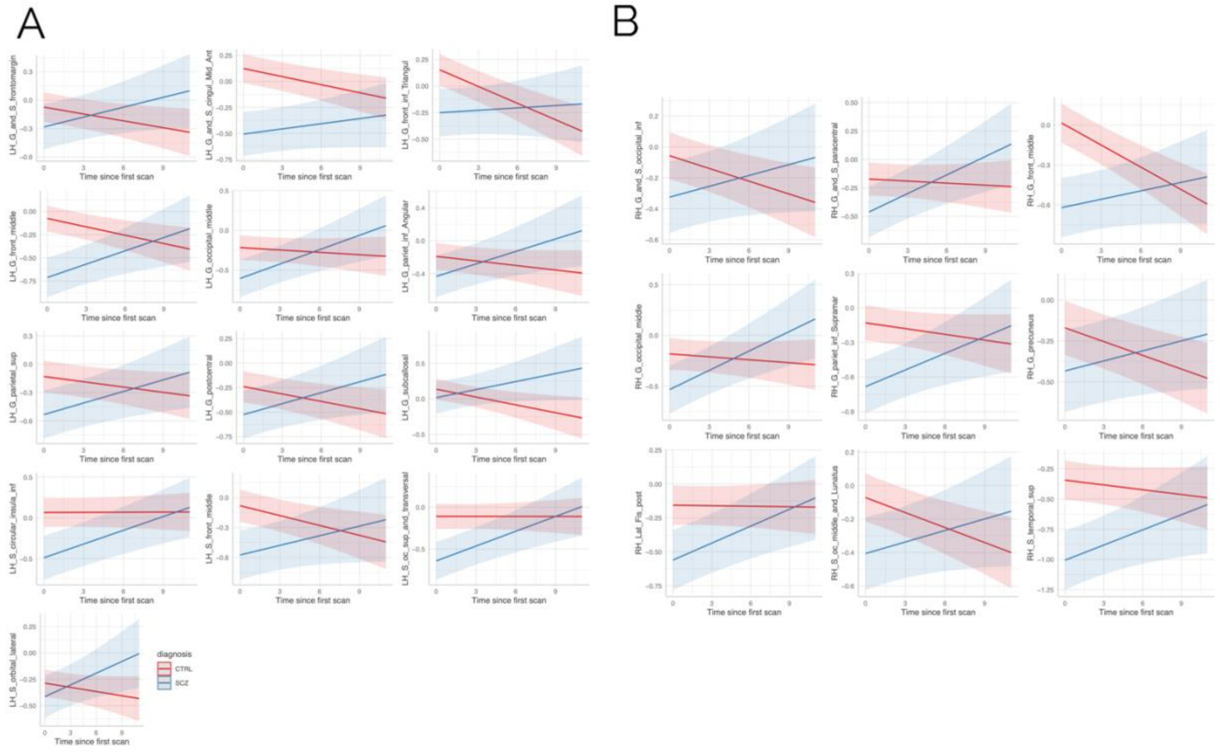


**Supplementary Figure 5**: Interaction effects between time and diagnosis group for each significant interaction effect for the left hemisphere (A) and the right hemisphere (B).

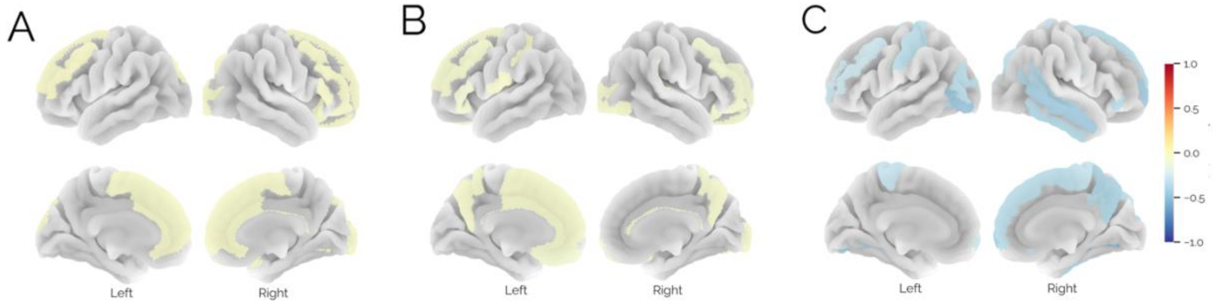


**Supplementary Figure 6:** Conditional main effects for age (A), time since inclusion (B) and sex (C) from the linear mixed model shown in Figure 2 in the main manuscript.

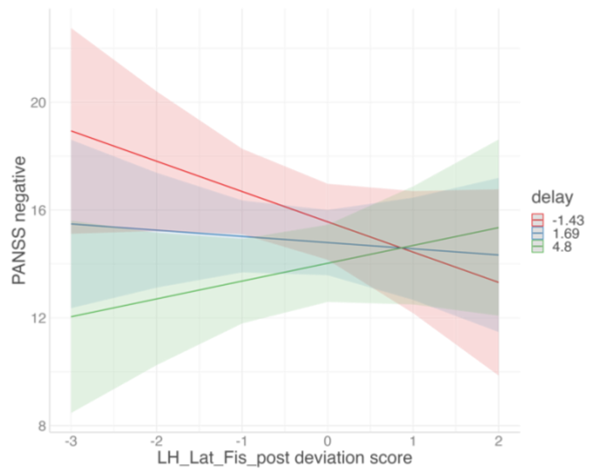


**Supplementary Figure 7**: Significant interaction effects (FDR corrected) between time points and deviation score of the posterior ramus of the lateral sulcus in the left hemisphere.

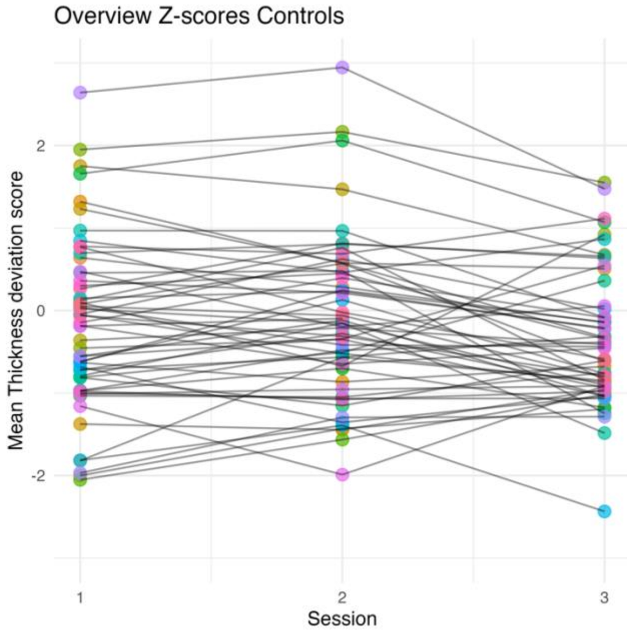


**Supplementary Figure 8:** The deviation in mean cortical thickness across timepoints for the healthy controls. This shows that the normative model removes age related effects such that the trajectory of each healthy participant is flat and with the distribution centred at zero.

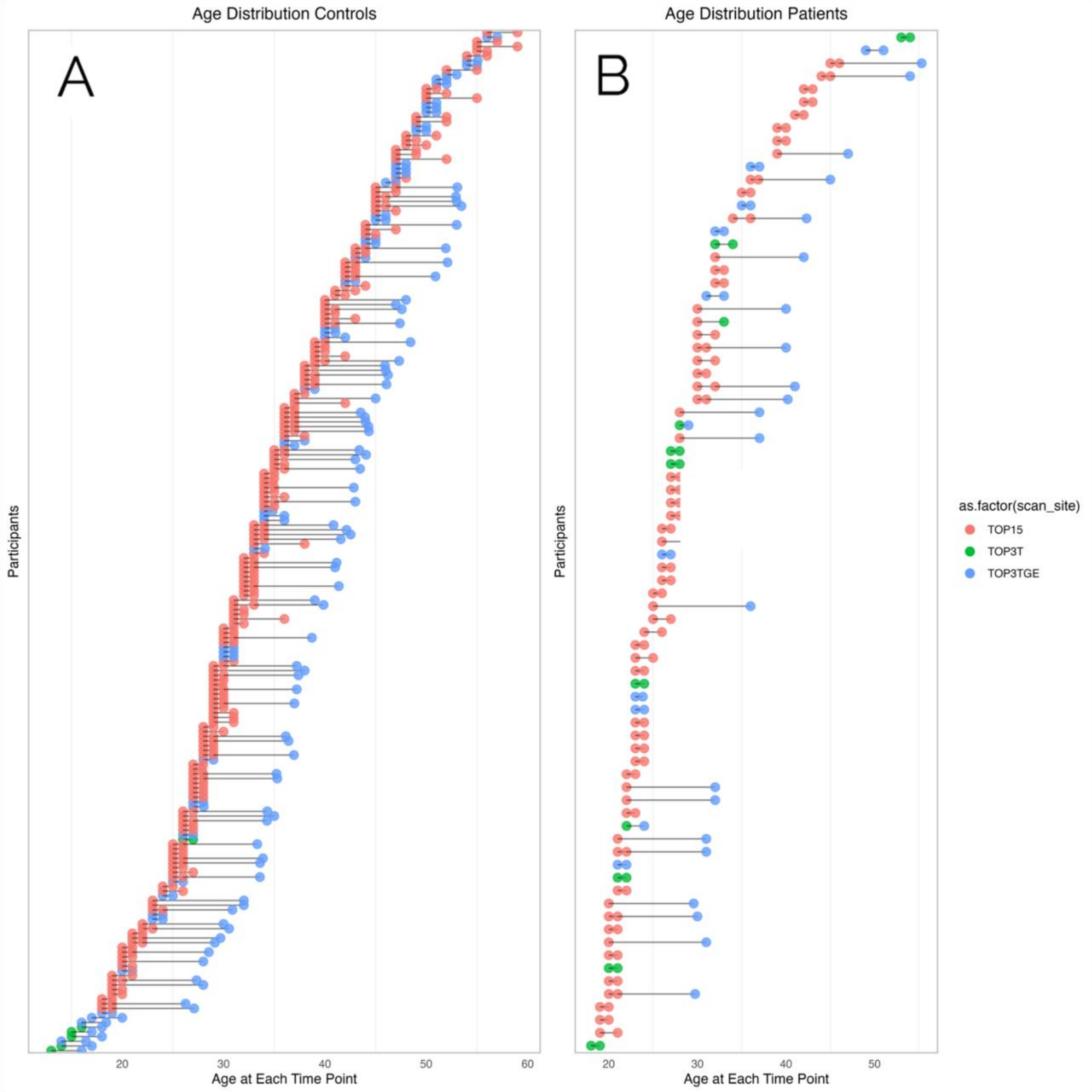


**Supplementary Figure 9:** age of each participant at each measurement timepoint for the healthy controls (A) and patients with schizophrenia (B). The colours indicate which scanner each subject was scanned on at each time point.

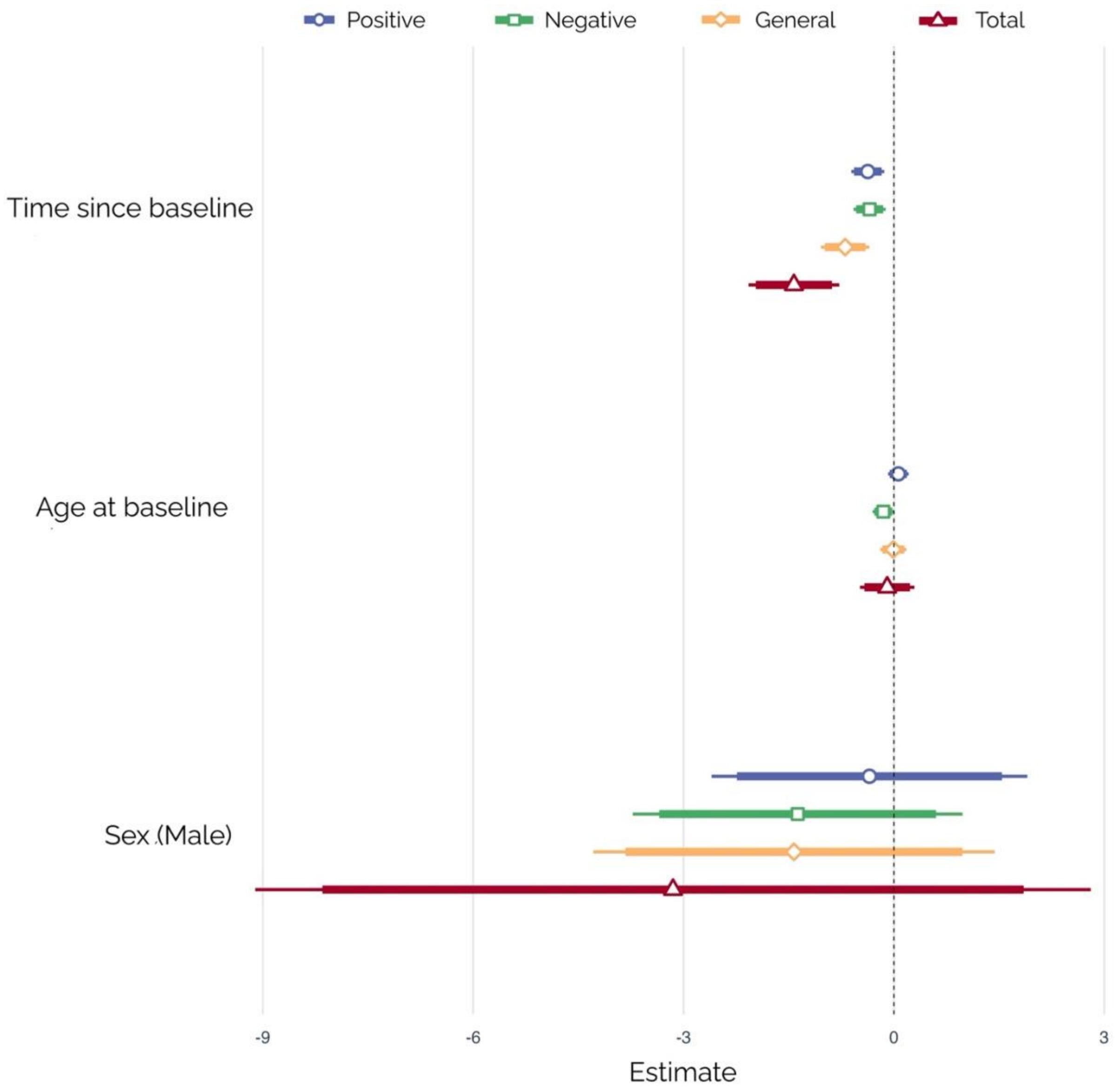


**Supplementary Figure 10:** Results from the LME models showing the estimates for time since inclusion (delay), age at baseline and sex.

**Supplementary Table 1**: adaptation samples (CTRL only) for the transfer of normative models to the three Oslo's scanners used in the longitudinal study.

| | TOP15 | TOP3T | TOP3T-GE |
|---|---|---|---|
| **Age mean** | 36.6 | 31.2 | 33.3 |
| **[SD]** | [10.3] | [7.7] | [11.1] |
| **Sex ratio (male)** | 42.7% | 48.8% | 49.8% |
| **n** | 103 | 294 | 393 |

**Supplementary Table 2**: ROIs with Significantly different deviation scores between patients with schizophrenia and healthy individuals, by time point (differences in deviation scores and p-values included):

| Session | ROI | Cohen's d | p-value |
|---|---|---|---|
| Session 1 | LH_G_and_S_occipital_inf | -0,351 | 0,006 |
| Session 1 | LH_G_and_S_paracentral | -0,337 | 0,011 |

| | | | |
|---|---|---|---|
| Session 1 | LH_G_and_S_subcentral | -0,348 | 0,009 |
| Session 1 | LH_G_and_S_cingul_Ant | -0,279 | 0,031 |
| Session 1 | LH_G_and_S_cingul_Mid_Ant | -0,525 | 0,000 |
| Session 1 | LH_G_and_S_cingul_Mid_Post | -0,532 | 0,000 |
| Session 1 | LH_G_front_inf_Triangul | -0,304 | 0,017 |
| Session 1 | LH_G_front_middle | -0,587 | 0,000 |
| Session 1 | LH_G_occipital_middle | -0,464 | 0,000 |
| Session 1 | LH_G_oc_temp_lat_fusifor | -0,292 | 0,019 |
| Session 1 | LH_G_oc_temp_med_Lingual | -0,275 | 0,041 |
| Session 1 | LH_G_pariet_inf_Angular | -0,315 | 0,018 |
| Session 1 | LH_G_parietal_sup | -0,420 | 0,002 |
| Session 1 | LH_G_postcentral | -0,279 | 0,034 |
| Session 1 | LH_G_precentral | -0,316 | 0,018 |
| Session 1 | LH_G_precuneus | -0,435 | 0,001 |
| Session 1 | LH_G_temp_sup_Lateral | -0,283 | 0,022 |
| Session 1 | LH_G_temp_sup_Plan_polar | -0,311 | 0,019 |
| Session 1 | LH_G_temp_sup_Plan_tempo | -0,416 | 0,001 |
| Session 1 | LH_G_temporal_middle | -0,376 | 0,004 |
| Session 1 | LH_Lat_Fis_ant_Horizont | -0,261 | 0,048 |
| Session 1 | LH_S_cingul_Marginalis | -0,403 | 0,003 |
| Session 1 | LH_S_circular_insula_inf | -0,489 | 0,000 |
| Session 1 | LH_S_circular_insula_sup | -0,325 | 0,012 |
| Session 1 | LH_S_collat_transv_ant | -0,374 | 0,005 |
| Session 1 | LH_S_collat_transv_post | -0,314 | 0,016 |
| Session 1 | LH_S_front_inf | -0,357 | 0,005 |
| Session 1 | LH_S_front_middle | -0,327 | 0,012 |
| Session 1 | LH_S_front_sup | -0,456 | 0,000 |
| Session 1 | LH_S_intrapariet_and_P_trans | -0,444 | 0,001 |
| Session 1 | LH_S_oc_middle_and_Lunatus | -0,285 | 0,031 |
| Session 1 | LH_S_oc_sup_and_transversal | -0,575 | 0,000 |
| Session 1 | LH_S_occipital_ant | -0,455 | 0,000 |
| Session 1 | LH_S_oc_temp_lat | -0,483 | 0,000 |
| Session 1 | LH_S_oc_temp_med_and_Lingual | -0,492 | 0,000 |
| Session 1 | LH_S_parieto_occipital | -0,426 | 0,001 |
| Session 1 | LH_S_pericallosal | 0,313 | 0,013 |
| Session 1 | LH_S_postcentral | -0,340 | 0,010 |
| Session 1 | LH_S_precentral_inf_part | -0,480 | 0,000 |
| Session 1 | LH_S_subparietal | -0,578 | 0,000 |
| Session 1 | LH_S_temporal_inf | -0,349 | 0,008 |
| Session 1 | LH_S_temporal_sup | -0,440 | 0,001 |
| Session 1 | RH_G_and_S_paracentral | -0,409 | 0,002 |
| Session 1 | RH_G_and_S_cingul_Mid_Ant | -0,387 | 0,004 |
| Session 1 | RH_G_and_S_cingul_Mid_Post | -0,327 | 0,012 |
| Session 1 | RH_G_front_inf_Opercular | -0,294 | 0,020 |
| Session 1 | RH_G_front_middle | -0,524 | 0,000 |

| | | | |
|---|---|---|---|
| Session 1 | RH_G_occipital_middle | -0,380 | 0,004 |
| Session 1 | RH_G_oc_temp_lat_fusifor | -0,305 | 0,020 |
| Session 1 | RH_G_orbital | -0,281 | 0,037 |
| Session 1 | RH_G_pariet_inf_Angular | -0,418 | 0,001 |
| Session 1 | RH_G_pariet_inf_Supramar | -0,462 | 0,000 |
| Session 1 | RH_G_parietal_sup | -0,292 | 0,031 |
| Session 1 | RH_G_postcentral | -0,360 | 0,006 |
| Session 1 | RH_G_precentral | -0,331 | 0,012 |
| Session 1 | RH_G_precuneus | -0,287 | 0,032 |
| Session 1 | RH_G_rectus | -0,305 | 0,022 |
| Session 1 | RH_G_temp_sup_Lateral | -0,406 | 0,001 |
| Session 1 | RH_G_temp_sup_Plan_tempo | -0,262 | 0,041 |
| Session 1 | RH_G_temporal_middle | -0,319 | 0,013 |
| Session 1 | RH_Lat_Fis_ant_Horizont | -0,320 | 0,012 |
| Session 1 | RH_Lat_Fis_post | -0,446 | 0,001 |
| Session 1 | RH_S_cingul_Marginalis | -0,439 | 0,001 |
| Session 1 | RH_S_circular_insula_inf | -0,525 | 0,000 |
| Session 1 | RH_S_circular_insula_sup | -0,442 | 0,001 |
| Session 1 | RH_S_collat_transv_ant | -0,305 | 0,020 |
| Session 1 | RH_S_front_inf | -0,460 | 0,000 |
| Session 1 | RH_S_front_middle | -0,356 | 0,008 |
| Session 1 | RH_S_front_sup | -0,433 | 0,001 |
| Session 1 | RH_S_intrapariet_and_P_trans | -0,453 | 0,000 |
| Session 1 | RH_S_oc_middle_and_Lunatus | -0,474 | 0,000 |
| Session 1 | RH_S_oc_sup_and_transversal | -0,454 | 0,001 |
| Session 1 | RH_S_occipital_ant | -0,257 | 0,048 |
| Session 1 | RH_S_oc_temp_lat | -0,509 | 0,000 |
| Session 1 | RH_S_oc_temp_med_and_Lingual | -0,311 | 0,016 |
| Session 1 | RH_S_orbital_lateral | -0,376 | 0,003 |
| Session 1 | RH_S_orbital_H_Shaped | -0,341 | 0,008 |
| Session 1 | RH_S_parieto_occipital | -0,275 | 0,033 |
| Session 1 | RH_S_pericallosal | 0,416 | 0,001 |
| Session 1 | RH_S_precentral_inf_part | -0,476 | 0,000 |
| Session 1 | RH_S_precentral_sup_part | -0,488 | 0,000 |
| Session 1 | RH_S_subparietal | -0,413 | 0,002 |
| Session 1 | RH_S_temporal_sup | -0,515 | 0,000 |
| Session 1 | Mean_thickness | -0,450 | 0,001 |
| Session 1 | Median_thickness | -0,466 | 0,000 |
| Session 2 | LH_G_and_S_subcentral | -0,431 | 0,001 |
| Session 2 | LH_G_and_S_transv_frontopol | -0,346 | 0,008 |
| Session 2 | LH_G_and_S_cingul_Ant | -0,307 | 0,021 |
| Session 2 | LH_G_and_S_cingul_Mid_Ant | -0,552 | 0,000 |
| Session 2 | LH_G_and_S_cingul_Mid_Post | -0,517 | 0,000 |
| Session 2 | LH_G_cingul_Post_dorsal | -0,280 | 0,033 |
| Session 2 | LH_G_front_inf_Orbital | -0,279 | 0,030 |

| | | | |
|---|---|---|---|
| Session 2 | LH_G_front_inf_Triangul | -0,381 | 0,003 |
| Session 2 | LH_G_front_middle | -0,399 | 0,003 |
| Session 2 | LH_G_parietal_sup | -0,298 | 0,027 |
| Session 2 | LH_G_precuneus | -0,451 | 0,001 |
| Session 2 | LH_G_temp_sup_Lateral | -0,394 | 0,002 |
| Session 2 | LH_G_temp_sup_Plan_tempo | -0,332 | 0,011 |
| Session 2 | LH_G_temporal_middle | -0,290 | 0,027 |
| Session 2 | LH_Lat_Fis_ant_Horizont | -0,367 | 0,005 |
| Session 2 | LH_Lat_Fis_ant_Vertical | -0,325 | 0,013 |
| Session 2 | LH_Lat_Fis_post | -0,364 | 0,007 |
| Session 2 | LH_S_cingul_Marginalis | -0,317 | 0,016 |
| Session 2 | LH_S_circular_insula_inf | -0,348 | 0,008 |
| Session 2 | LH_S_circular_insula_sup | -0,396 | 0,002 |
| Session 2 | LH_S_front_inf | -0,413 | 0,002 |
| Session 2 | LH_S_front_middle | -0,261 | 0,040 |
| Session 2 | LH_S_front_sup | -0,412 | 0,001 |
| Session 2 | LH_S_interm_prim_Jensen | -0,320 | 0,019 |
| Session 2 | LH_S_intrapariet_and_P_trans | -0,360 | 0,006 |
| Session 2 | LH_S_oc_sup_and_transversal | -0,461 | 0,001 |
| Session 2 | LH_S_occipital_ant | -0,340 | 0,009 |
| Session 2 | LH_S_oc_temp_lat | -0,391 | 0,004 |
| Session 2 | LH_S_oc_temp_med_and_Lingual | -0,321 | 0,015 |
| Session 2 | LH_S_parieto_occipital | -0,459 | 0,001 |
| Session 2 | LH_S_postcentral | -0,261 | 0,050 |
| Session 2 | LH_S_precentral_inf_part | -0,431 | 0,001 |
| Session 2 | LH_S_subparietal | -0,541 | 0,000 |
| Session 2 | LH_S_temporal_sup | -0,345 | 0,010 |
| Session 2 | RH_G_and_S_cingul_Mid_Ant | -0,425 | 0,002 |
| Session 2 | RH_G_and_S_cingul_Mid_Post | -0,470 | 0,000 |
| Session 2 | RH_G_front_inf_Opercular | -0,312 | 0,014 |
| Session 2 | RH_G_front_inf_Orbital | -0,288 | 0,026 |
| Session 2 | RH_G_front_middle | -0,426 | 0,001 |
| Session 2 | RH_G_oc_temp_lat_fusifor | -0,296 | 0,025 |
| Session 2 | RH_G_pariet_inf_Supramar | -0,367 | 0,004 |
| Session 2 | RH_G_subcallosal | -0,288 | 0,037 |
| Session 2 | RH_G_temp_sup_Lateral | -0,325 | 0,009 |
| Session 2 | RH_G_temporal_inf | -0,262 | 0,042 |
| Session 2 | RH_G_temporal_middle | -0,314 | 0,016 |
| Session 2 | RH_Lat_Fis_ant_Horizont | -0,359 | 0,005 |
| Session 2 | RH_Lat_Fis_post | -0,330 | 0,015 |
| Session 2 | RH_S_cingul_Marginalis | -0,310 | 0,017 |
| Session 2 | RH_S_circular_insula_inf | -0,427 | 0,001 |
| Session 2 | RH_S_circular_insula_sup | -0,581 | 0,000 |
| Session 2 | RH_S_collat_transv_ant | -0,434 | 0,002 |
| Session 2 | RH_S_front_inf | -0,360 | 0,005 |

| | | | |
|---|---|---|---|
| Session 2 | RH_S_front_middle | -0,373 | 0,004 |
| Session 2 | RH_S_intrapariet_and_P_trans | -0,287 | 0,029 |
| Session 2 | RH_S_oc_middle_and_Lunatus | -0,310 | 0,023 |
| Session 2 | RH_S_oc_sup_and_transversal | -0,359 | 0,006 |
| Session 2 | RH_S_occipital_ant | -0,265 | 0,042 |
| Session 2 | RH_S_oc_temp_lat | -0,569 | 0,000 |
| Session 2 | RH_S_oc_temp_med_and_Lingual | -0,351 | 0,008 |
| Session 2 | RH_S_orbital_lateral | -0,290 | 0,023 |
| Session 2 | RH_S_orbital_H_Shaped | -0,276 | 0,031 |
| Session 2 | RH_S_parieto_occipital | -0,318 | 0,013 |
| Session 2 | RH_S_pericallosal | 0,386 | 0,003 |
| Session 2 | RH_S_precentral_inf_part | -0,350 | 0,007 |
| Session 2 | RH_S_precentral_sup_part | -0,316 | 0,020 |
| Session 2 | RH_S_subparietal | -0,300 | 0,024 |
| Session 2 | RH_S_temporal_sup | -0,426 | 0,002 |
| Session 2 | Mean_thickness | -0,419 | 0,001 |
| Session 2 | Median_thickness | -0,458 | 0,001 |
| Session 3 | LH_G_subcallosal | 0,532 | 0,034 |
| Session 3 | LH_S_orbital_lateral | 1,088 | 0,004 |
| Session 3 | RH_G_subcallosal | 0,604 | 0,037 |

**Supplementary Table 3**: ROIs with Significantly different overlap statistics of extreme negative deviation scores between patients with schizophrenia and healthy individuals.

| Session | ROI | Cramer´s V | P-value |
|---|---|---|---|
| session 1 | RH_Lateral aspect of the superior temporal gyrus | 0.191176 | 5e-06 |
| session 1 | RH_Opercular part of the inferior frontal gyrus | 0.183738 | 1.2e-05 |
| session 1 | LH_Superior frontal sulcus | 0.179788 | 1.8e-05 |
| session 1 | RH_Superior temporal sulcus (parallel sulcus) | 0.169442 | 5.2e-05 |
| session 1 | RH_Inferior frontal sulcus | 0.169102 | 5.4e-05 |
| session 1 | LH_Inferior segment of the circular sulcus of the insula | 0.165028 | 8.1e-05 |
| session 1 | LH_Superior occipital sulcus and transverse occipital sulcus | 0.150891 | 0.000315 |
| session 1 | LH_Intraparietal sulcus (interparietal sulcus) and transverse parietal sulci | 0.14866 | 0.000386 |
| session 1 | LH_Medial occipito_temporal sulcus (collateral sulcus) and lingual sulcus | 0.147875 | 0.000415 |
| session 1 | RH_Posterior ramus (or segment) of the lateral sulcus (or fissure) | 0.146971 | 0.00045 |
| session 1 | RH_Marginal branch (or part) of the cingulate sulcus | 0.142267 | 0.000682 |

| | | | |
|---|---|---|---|
| session 1 | RH_Inferior segment of the circular sulcus of the insula | 0.139753 | 0.000848 |
| session 1 | LH_Lateral occipito_temporal sulcus | 0.139753 | 0.000848 |
| session 1 | RH_Anterior part of the cingulate gyrus and sulcus (ACC) | 0.136582 | 0.001111 |
| session 1 | LH_Inferior occipital gyrus (O3) and sulcus | 0.135561 | 0.00121 |
| session 1 | RH_Supramarginal gyrus | 0.130944 | 0.001771 |
| session 1 | RH_Subparietal sulcus | 0.12997 | 0.001916 |
| session 1 | LH_Lateral aspect of the superior temporal gyrus | 0.127184 | 0.002394 |
| session 1 | LH_Planum temporale or temporal plane of the superior temporal gyrus | 0.12652 | 0.002523 |
| session 1 | LH_Subparietal sulcus | 0.122266 | 0.003511 |
| session 1 | RH_Lateral orbital sulcus | 0.121487 | 0.003726 |
| session 1 | RH_Lateral occipito_temporal gyrus (fusiform gyrus, O4_T4) | 0.11996 | 0.004183 |
| session 1 | LH_Inferior part of the precentral sulcus | 0.118665 | 0.00461 |
| session 1 | LH_Anterior occipital sulcus and preoccipital notch (temporo_occipital incisure) | 0.116843 | 0.005277 |
| session 1 | RH_Inferior part of the precentral sulcus | 0.114195 | 0.006404 |
| session 1 | LH_Middle occipital gyrus (O2, lateral occipital gyrus) | 0.114195 | 0.006404 |
| session 1 | LH_Inferior frontal sulcus | 0.110349 | 0.008425 |
| session 2 | RH_Superior segment of the circular sulcus of the insula | 0.159154 | 0.000145 |

**Supplementary Table 4**: Percentage (%) of extreme positive deviation overlap by ROI, by time point, for controls and patients with schizophrenia.

| **Session** | **ROI** | **Controls** | **Patients** |
|---|---|---|---|
| **session 1** | LH_Fronto_marginal gyrus (of Wernicke) and sulcus | 4.6 | 1.3 |
| **session 1** | LH_Inferior occipital gyrus (O3) and sulcus | 3.7 | 1.3 |
| **session 1** | LH_Paracentral lobule and sulcus | 0.9 | 0.0 |
| **session 1** | LH_Subcentral gyrus (central operculum) and sulci | 2.3 | 1.3 |
| **session 1** | LH_Transverse frontopolar gyri and sulci | 2.3 | 5.1 |
| **session 1** | LH_Anterior part of the cingulate gyrus and sulcus (ACC) | 1.4 | 0.0 |
| **session 1** | LH_Middle_anterior part of the cingulate gyrus and sulcus (aMCC) | 4.1 | 1.3 |
| **session 1** | LH_Middle_posterior part of the cingulate gyrus and sulcus (pMCC) | 2.8 | 0.0 |
| **session 1** | LH_Posterior_dorsal part of the cingulate gyrus (dPCC) | 5.5 | 5.1 |
| **session 1** | LH_Posterior_ventral part of the cingulate gyrus (vPCC, isthmus of the cingulate gyrus) | 3.2 | 2.5 |
| **session 1** | LH_Cuneus (O6) | 3.2 | 1.3 |
| **session 1** | LH_Opercular part of the inferior frontal gyrus | 0.5 | 0.0 |
| **session 1** | LH_Orbital part of the inferior frontal gyrus | 1.4 | 2.5 |
| **session 1** | LH_Triangular part of the inferior frontal gyrus | 2.3 | 2.5 |
| **session 1** | LH_Middle frontal gyrus (F2) | 0.9 | 0.0 |
| **session 1** | LH_Superior frontal gyrus (F1) | 2.8 | 0.0 |
| **session 1** | LH_Long insular gyrus and central sulcus of the insula | 0.9 | 1.3 |
| **session 1** | LH_Short insular gyri | 3.2 | 1.3 |
| **session 1** | LH_Middle occipital gyrus (O2, lateral occipital gyrus) | 2.3 | 1.3 |
| **session 1** | LH_Superior occipital gyrus (O1) | 0.9 | 0.0 |
| **session 1** | LH_Lateral occipito_temporal gyrus (fusiform gyrus, O4_T4) | 1.8 | 2.5 |

| | | | |
|---|---|---|---|
| **session 1** | LH_Lingual gyrus, ligual part of the medial occipito_temporal gyrus, (O5) | 2.3 | 0.0 |
| **session 1** | LH_Parahippocampal gyrus, parahippocampal part of the medial occipito_temporal gyrus, (T5) | 0.9 | 3.8 |
| **session 1** | LH_Orbital gyri | 2.3 | 2.5 |
| **session 1** | LH_Angular gyrus | 5.0 | 2.5 |
| **session 1** | LH_Supramarginal gyrus | 2.8 | 1.3 |
| **session 1** | LH_Superior parietal lobule (lateral part of P1) | 5.5 | 0.0 |
| **session 1** | LH_Postcentral gyrus | 3.7 | 1.3 |
| **session 1** | LH_Precentral gyrus | 0.9 | 0.0 |
| **session 1** | LH_Precuneus (medial part of P1) | 3.2 | 2.5 |
| **session 1** | LH_Straight gyrus, Gyrus rectus | 4.6 | 3.8 |
| **session 1** | LH_Subcallosal area, subcallosal gyrus | 5.0 | 6.3 |
| **session 1** | LH_Anterior transverse temporal gyrus (of Heschl) | 1.8 | 0.0 |
| **session 1** | LH_Lateral aspect of the superior temporal gyrus | 2.3 | 2.5 |
| **session 1** | LH_Planum polare of the superior temporal gyrus | 2.8 | 1.3 |
| **session 1** | LH_Planum temporale or temporal plane of the superior temporal gyrus | 0.9 | 0.0 |
| **session 1** | LH_Inferior temporal gyrus (T3) | 3.7 | 6.3 |
| **session 1** | LH_Middle temporal gyrus (T2) | 0.9 | 1.3 |
| **session 1** | LH_Horizontal ramus of the anterior segment of the lateral sulcus (or fissure) | 1.8 | 5.1 |
| **session 1** | LH_Vertical ramus of the anterior segment of the lateral sulcus (or fissure) | 0.9 | 2.5 |
| **session 1** | LH_Posterior ramus (or segment) of the lateral sulcus (or fissure) | 2.3 | 1.3 |
| **session 1** | LH_Occipital pole | 1.8 | 1.3 |
| **session 1** | LH_Temporal pole | 2.3 | 0.0 |
| **session 1** | LH_Calcarine sulcus | 3.7 | 2.5 |
| **session 1** | LH_Central sulcus (Rolandos fissure) | 4.1 | 5.1 |
| **session 1** | LH_Marginal branch (or part) of the cingulate sulcus | 2.8 | 1.3 |
| **session 1** | LH_Anterior segment of the circular sulcus of the insula | 2.3 | 0.0 |
| **session 1** | LH_Inferior segment of the circular sulcus of the insula | 2.8 | 2.5 |
| **session 1** | LH_Superior segment of the circular sulcus of the insula | 1.8 | 1.3 |
| **session 1** | LH_Anterior transverse collateral sulcus | 1.8 | 0.0 |
| **session 1** | LH_Posterior transverse collateral sulcus | 2.3 | 0.0 |
| **session 1** | LH_Inferior frontal sulcus | 2.3 | 3.8 |
| **session 1** | LH_Middle frontal sulcus | 2.8 | 1.3 |
| **session 1** | LH_Superior frontal sulcus | 0.9 | 1.3 |
| **session 1** | LH_Sulcus intermedius primus (of Jensen) | 5.5 | 2.5 |
| **session 1** | LH_Intraparietal sulcus (interparietal sulcus) and transverse parietal sulci | 3.2 | 1.3 |
| **session 1** | LH_Middle occipital sulcus and lunatus sulcus | 2.8 | 1.3 |
| **session 1** | LH_Superior occipital sulcus and transverse occipital sulcus | 3.7 | 0.0 |
| **session 1** | LH_Anterior occipital sulcus and preoccipital notch (temporo_occipital incisure) | 2.3 | 0.0 |
| **session 1** | LH_Lateral occipito_temporal sulcus | 3.2 | 1.3 |
| **session 1** | LH_Medial occipito_temporal sulcus (collateral sulcus) and lingual sulcus | 2.3 | 1.3 |
| **session 1** | LH_Lateral orbital sulcus | 0.9 | 3.8 |
| **session 1** | LH_Medial orbital sulcus (olfactory sulcus) | 2.3 | 1.3 |

| | | | |
|---|---|---|---|
| **session 1** | LH_Orbital sulci (H_shaped sulci) | 5.5 | 2.5 |
| **session 1** | LH_Parieto_occipital sulcus (or fissure) | 3.7 | 1.3 |
| **session 1** | LH_Pericallosal sulcus (S of corpus callosum) | 4.1 | 12.7 |
| **session 1** | LH_Postcentral sulcus | 2.8 | 0.0 |
| **session 1** | LH_Inferior part of the precentral sulcus | 1.8 | 1.3 |
| **session 1** | LH_Superior part of the precentral sulcus | 2.3 | 2.5 |
| **session 1** | LH_Suborbital sulcus (sulcus rostrales, supraorbital sulcus) | 2.3 | 3.8 |
| **session 1** | LH_Subparietal sulcus | 6.9 | 3.8 |
| **session 1** | LH_Inferior temporal sulcus | 7.3 | 2.5 |
| **session 1** | LH_Superior temporal sulcus (parallel sulcus) | 1.8 | 0.0 |
| **session 1** | LH_Transverse temporal sulcus | 1.8 | 1.3 |
| **session 1** | RH_Fronto_marginal gyrus (of Wernicke) and sulcus | 2.8 | 1.3 |
| **session 1** | RH_Inferior occipital gyrus (O3) and sulcus | 2.3 | 5.1 |
| **session 1** | RH_Paracentral lobule and sulcus | 2.8 | 0.0 |
| **session 1** | RH_Subcentral gyrus (central operculum) and sulci | 0.9 | 1.3 |
| **session 1** | RH_Transverse frontopolar gyri and sulci | 5.5 | 0.0 |
| **session 1** | RH_Anterior part of the cingulate gyrus and sulcus (ACC) | 4.6 | 1.3 |
| **session 1** | RH_Middle_anterior part of the cingulate gyrus and sulcus (aMCC) | 1.8 | 1.3 |
| **session 1** | RH_Middle_posterior part of the cingulate gyrus and sulcus (pMCC) | 2.3 | 2.5 |
| **session 1** | RH_Posterior_dorsal part of the cingulate gyrus (dPCC) | 2.3 | 1.3 |
| **session 1** | RH_Posterior_ventral part of the cingulate gyrus (vPCC, isthmus of the cingulate gyrus) | 1.8 | 0.0 |
| **session 1** | RH_Cuneus (O6) | 2.8 | 0.0 |
| **session 1** | RH_Opercular part of the inferior frontal gyrus | 1.8 | 1.3 |
| **session 1** | RH_Orbital part of the inferior frontal gyrus | 1.4 | 1.3 |
| **session 1** | RH_Triangular part of the inferior frontal gyrus | 0.9 | 2.5 |
| **session 1** | RH_Middle frontal gyrus (F2) | 2.3 | 0.0 |
| **session 1** | RH_Superior frontal gyrus (F1) | 2.8 | 0.0 |
| **session 1** | RH_Long insular gyrus and central sulcus of the insula | 1.8 | 2.5 |
| **session 1** | RH_Short insular gyri | 1.8 | 2.5 |
| **session 1** | RH_Middle occipital gyrus (O2, lateral occipital gyrus) | 0.9 | 1.3 |
| **session 1** | RH_Superior occipital gyrus (O1) | 2.3 | 1.3 |
| **session 1** | RH_Lateral occipito_temporal gyrus (fusiform gyrus, O4_T4) | 2.3 | 2.5 |
| **session 1** | RH_Lingual gyrus, ligual part of the medial occipito_temporal gyrus, (O5) | 1.8 | 0.0 |
| **session 1** | RH_Parahippocampal gyrus, parahippocampal part of the medial occipito_temporal gyrus, (T5) | 2.3 | 2.5 |
| **session 1** | RH_Orbital gyri | 4.1 | 2.5 |
| **session 1** | RH_Angular gyrus | 0.9 | 1.3 |
| **session 1** | RH_Supramarginal gyrus | 1.8 | 1.3 |
| **session 1** | RH_Superior parietal lobule (lateral part of P1) | 1.4 | 0.0 |
| **session 1** | RH_Postcentral gyrus | 4.1 | 3.8 |
| **session 1** | RH_Precentral gyrus | 0.5 | 0.0 |
| **session 1** | RH_Precuneus (medial part of P1) | 3.7 | 1.3 |
| **session 1** | RH_Straight gyrus, Gyrus rectus | 1.8 | 2.5 |
| **session 1** | RH_Subcallosal area, subcallosal gyrus | 4.1 | 0.0 |
| **session 1** | RH_Anterior transverse temporal gyrus (of Heschl) | 3.7 | 0.0 |

| | | | |
|---|---|---|---|
| **session 1** | RH_Lateral aspect of the superior temporal gyrus | 1.4 | 1.3 |
| **session 1** | RH_Planum polare of the superior temporal gyrus | 1.4 | 1.3 |
| **session 1** | RH_Planum temporale or temporal plane of the superior temporal gyrus | 0.5 | 2.5 |
| **session 1** | RH_Inferior temporal gyrus (T3) | 1.8 | 2.5 |
| **session 1** | RH_Middle temporal gyrus (T2) | 3.2 | 3.8 |
| **session 1** | RH_Horizontal ramus of the anterior segment of the lateral sulcus (or fissure) | 1.8 | 1.3 |
| **session 1** | RH_Vertical ramus of the anterior segment of the lateral sulcus (or fissure) | 5.0 | 6.3 |
| **session 1** | RH_Posterior ramus (or segment) of the lateral sulcus (or fissure) | 2.8 | 0.0 |
| **session 1** | RH_Occipital pole | 3.7 | 3.8 |
| **session 1** | RH_Temporal pole | 4.1 | 5.1 |
| **session 1** | RH_Calcarine sulcus | 5.5 | 3.8 |
| **session 1** | RH_Central sulcus (Rolandos fissure) | 4.1 | 2.5 |
| **session 1** | RH_Marginal branch (or part) of the cingulate sulcus | 2.3 | 1.3 |
| **session 1** | RH_Anterior segment of the circular sulcus of the insula | 1.4 | 1.3 |
| **session 1** | RH_Inferior segment of the circular sulcus of the insula | 2.3 | 2.5 |
| **session 1** | RH_Superior segment of the circular sulcus of the insula | 0.5 | 1.3 |
| **session 1** | RH_Anterior transverse collateral sulcus | 2.3 | 0.0 |
| **session 1** | RH_Posterior transverse collateral sulcus | 0.5 | 0.0 |
| **session 1** | RH_Inferior frontal sulcus | 3.7 | 2.5 |
| **session 1** | RH_Middle frontal sulcus | 4.1 | 2.5 |
| **session 1** | RH_Superior frontal sulcus | 0.5 | 2.5 |
| **session 1** | RH_Sulcus intermedius primus (of Jensen) | 0.9 | 0.0 |
| **session 1** | RH_Intraparietal sulcus (interparietal sulcus) and transverse parietal sulci | 0.9 | 0.0 |
| **session 1** | RH_Middle occipital sulcus and lunatus sulcus | 3.7 | 0.0 |
| **session 1** | RH_Superior occipital sulcus and transverse occipital sulcus | 3.2 | 0.0 |
| **session 1** | RH_Anterior occipital sulcus and preoccipital notch (temporo_occipital incisure) | 3.2 | 2.5 |
| **session 1** | RH_Lateral occipito_temporal sulcus | 2.8 | 2.5 |
| **session 1** | RH_Medial occipito_temporal sulcus (collateral sulcus) and lingual sulcus | 1.8 | 2.5 |
| **session 1** | RH_Lateral orbital sulcus | 5.0 | 2.5 |
| **session 1** | RH_Medial orbital sulcus (olfactory sulcus) | 1.8 | 0.0 |
| **session 1** | RH_Orbital sulci (H_shaped sulci) | 3.2 | 1.3 |
| **session 1** | RH_Parieto_occipital sulcus (or fissure) | 2.3 | 1.3 |
| **session 1** | RH_Pericallosal sulcus (S of corpus callosum) | 6.0 | 13.9 |
| **session 1** | RH_Postcentral sulcus | 2.8 | 0.0 |
| **session 1** | RH_Inferior part of the precentral sulcus | 0.5 | 0.0 |
| **session 1** | RH_Superior part of the precentral sulcus | 0.9 | 0.0 |
| **session 1** | RH_Suborbital sulcus (sulcus rostrales, supraorbital sulcus) | 2.3 | 2.5 |
| **session 1** | RH_Subparietal sulcus | 4.1 | 2.5 |
| **session 1** | RH_Inferior temporal sulcus | 2.3 | 2.5 |
| **session 1** | RH_Superior temporal sulcus (parallel sulcus) | 2.8 | 0.0 |
| **session 1** | RH_Transverse temporal sulcus | 2.3 | 0.0 |
| **session 2** | LH_Fronto_marginal gyrus (of Wernicke) and sulcus | 3.2 | 0.0 |

| | | | |
|---|---|---|---|
| **session 2** | LH_Inferior occipital gyrus (O3) and sulcus | 3.2 | 1.3 |
| **session 2** | LH_Paracentral lobule and sulcus | 2.3 | 1.3 |
| **session 2** | LH_Subcentral gyrus (central operculum) and sulci | 1.8 | 0.0 |
| **session 2** | LH_Transverse frontopolar gyri and sulci | 2.3 | 0.0 |
| **session 2** | LH_Anterior part of the cingulate gyrus and sulcus (ACC) | 1.4 | 1.3 |
| **session 2** | LH_Middle_anterior part of the cingulate gyrus and sulcus (aMCC) | 1.4 | 0.0 |
| **session 2** | LH_Middle_posterior part of the cingulate gyrus and sulcus (pMCC) | 1.4 | 0.0 |
| **session 2** | LH_Posterior_dorsal part of the cingulate gyrus (dPCC) | 4.1 | 1.3 |
| **session 2** | LH_Posterior_ventral part of the cingulate gyrus (vPCC, isthmus of the cingulate gyrus) | 2.3 | 2.5 |
| **session 2** | LH_Cuneus (O6) | 2.3 | 0.0 |
| **session 2** | LH_Opercular part of the inferior frontal gyrus | 0.9 | 0.0 |
| **session 2** | LH_Orbital part of the inferior frontal gyrus | 1.8 | 1.3 |
| **session 2** | LH_Triangular part of the inferior frontal gyrus | 2.8 | 3.8 |
| **session 2** | LH_Middle frontal gyrus (F2) | 2.8 | 0.0 |
| **session 2** | LH_Superior frontal gyrus (F1) | 1.4 | 0.0 |
| **session 2** | LH_Long insular gyrus and central sulcus of the insula | 0.0 | 2.5 |
| **session 2** | LH_Short insular gyri | 3.2 | 2.5 |
| **session 2** | LH_Middle occipital gyrus (O2, lateral occipital gyrus) | 1.4 | 1.3 |
| **session 2** | LH_Superior occipital gyrus (O1) | 0.5 | 0.0 |
| **session 2** | LH_Lateral occipito_temporal gyrus (fusiform gyrus, O4_T4) | 1.8 | 5.1 |
| **session 2** | LH_Lingual gyrus, ligual part of the medial occipito_temporal gyrus, (O5) | 0.5 | 0.0 |
| **session 2** | LH_Parahippocampal gyrus, parahippocampal part of the medial occipito_temporal gyrus, (T5) | 1.8 | 3.8 |
| **session 2** | LH_Orbital gyri | 1.4 | 5.1 |
| **session 2** | LH_Angular gyrus | 2.8 | 1.3 |
| **session 2** | LH_Supramarginal gyrus | 1.8 | 2.5 |
| **session 2** | LH_Superior parietal lobule (lateral part of P1) | 3.2 | 0.0 |
| **session 2** | LH_Postcentral gyrus | 3.7 | 2.5 |
| **session 2** | LH_Precentral gyrus | 0.9 | 0.0 |
| **session 2** | LH_Precuneus (medial part of P1) | 2.3 | 1.3 |
| **session 2** | LH_Straight gyrus, Gyrus rectus | 2.8 | 5.1 |
| **session 2** | LH_Subcallosal area, subcallosal gyrus | 4.1 | 3.8 |
| **session 2** | LH_Anterior transverse temporal gyrus (of Heschl) | 0.9 | 1.3 |
| **session 2** | LH_Lateral aspect of the superior temporal gyrus | 1.8 | 3.8 |
| **session 2** | LH_Planum polare of the superior temporal gyrus | 1.8 | 2.5 |
| **session 2** | LH_Planum temporale or temporal plane of the superior temporal gyrus | 2.3 | 1.3 |
| **session 2** | LH_Inferior temporal gyrus (T3) | 2.8 | 3.8 |
| **session 2** | LH_Middle temporal gyrus (T2) | 2.3 | 0.0 |
| **session 2** | LH_Horizontal ramus of the anterior segment of the lateral sulcus (or fissure) | 3.2 | 3.8 |
| **session 2** | LH_Vertical ramus of the anterior segment of the lateral sulcus (or fissure) | 0.9 | 2.5 |
| **session 2** | LH_Posterior ramus (or segment) of the lateral sulcus (or fissure) | 1.8 | 1.3 |
| **session 2** | LH_Occipital pole | 2.8 | 0.0 |
| **session 2** | LH_Temporal pole | 1.4 | 0.0 |

| | | | |
|---|---|---|---|
| **session 2** | LH_Calcarine sulcus | 5.0 | 1.3 |
| **session 2** | LH_Central sulcus (Rolandos fissure) | 1.8 | 2.5 |
| **session 2** | LH_Marginal branch (or part) of the cingulate sulcus | 0.9 | 0.0 |
| **session 2** | LH_Anterior segment of the circular sulcus of the insula | 2.3 | 1.3 |
| **session 2** | LH_Inferior segment of the circular sulcus of the insula | 3.7 | 2.5 |
| **session 2** | LH_Superior segment of the circular sulcus of the insula | 0.9 | 0.0 |
| **session 2** | LH_Anterior transverse collateral sulcus | 1.8 | 2.5 |
| **session 2** | LH_Posterior transverse collateral sulcus | 1.8 | 2.5 |
| **session 2** | LH_Inferior frontal sulcus | 1.8 | 2.5 |
| **session 2** | LH_Middle frontal sulcus | 1.4 | 2.5 |
| **session 2** | LH_Superior frontal sulcus | 0.9 | 3.8 |
| **session 2** | LH_Sulcus intermedius primus (of Jensen) | 6.0 | 2.5 |
| **session 2** | LH_Intraparietal sulcus (interparietal sulcus) and transverse parietal sulci | 2.3 | 0.0 |
| **session 2** | LH_Middle occipital sulcus and lunatus sulcus | 3.7 | 5.1 |
| **session 2** | LH_Superior occipital sulcus and transverse occipital sulcus | 1.4 | 1.3 |
| **session 2** | LH_Anterior occipital sulcus and preoccipital notch (temporo_occipital incisure) | 2.3 | 2.5 |
| **session 2** | LH_Lateral occipito_temporal sulcus | 1.8 | 1.3 |
| **session 2** | LH_Medial occipito_temporal sulcus (collateral sulcus) and lingual sulcus | 2.3 | 1.3 |
| **session 2** | LH_Lateral orbital sulcus | 1.8 | 0.0 |
| **session 2** | LH_Medial orbital sulcus (olfactory sulcus) | 3.2 | 3.8 |
| **session 2** | LH_Orbital sulci (H_shaped sulci) | 7.3 | 3.8 |
| **session 2** | LH_Parieto_occipital sulcus (or fissure) | 4.6 | 0.0 |
| **session 2** | LH_Pericallosal sulcus (S of corpus callosum) | 3.7 | 8.9 |
| **session 2** | LH_Postcentral sulcus | 1.8 | 0.0 |
| **session 2** | LH_Inferior part of the precentral sulcus | 0.5 | 2.5 |
| **session 2** | LH_Superior part of the precentral sulcus | 1.8 | 2.5 |
| **session 2** | LH_Suborbital sulcus (sulcus rostrales, supraorbital sulcus) | 1.8 | 1.3 |
| **session 2** | LH_Subparietal sulcus | 3.2 | 0.0 |
| **session 2** | LH_Inferior temporal sulcus | 6.0 | 5.1 |
| **session 2** | LH_Superior temporal sulcus (parallel sulcus) | 0.5 | 1.3 |
| **session 2** | LH_Transverse temporal sulcus | 0.5 | 3.8 |
| **session 2** | RH_Fronto_marginal gyrus (of Wernicke) and sulcus | 2.3 | 5.1 |
| **session 2** | RH_Inferior occipital gyrus (O3) and sulcus | 1.4 | 2.5 |
| **session 2** | RH_Paracentral lobule and sulcus | 1.4 | 3.8 |
| **session 2** | RH_Subcentral gyrus (central operculum) and sulci | 0.9 | 3.8 |
| **session 2** | RH_Transverse frontopolar gyri and sulci | 3.2 | 1.3 |
| **session 2** | RH_Anterior part of the cingulate gyrus and sulcus (ACC) | 2.3 | 1.3 |
| **session 2** | RH_Middle_anterior part of the cingulate gyrus and sulcus (aMCC) | 0.5 | 0.0 |
| **session 2** | RH_Middle_posterior part of the cingulate gyrus and sulcus (pMCC) | 1.4 | 1.3 |
| **session 2** | RH_Posterior_dorsal part of the cingulate gyrus (dPCC) | 1.4 | 1.3 |
| **session 2** | RH_Posterior_ventral part of the cingulate gyrus (vPCC, isthmus of the cingulate gyrus) | 1.4 | 1.3 |
| **session 2** | RH_Cuneus (O6) | 1.4 | 1.3 |
| **session 2** | RH_Opercular part of the inferior frontal gyrus | 1.8 | 0.0 |

| | | | |
|---|---|---|---|
| **session 2** | RH_Orbital part of the inferior frontal gyrus | 3.7 | 1.3 |
| **session 2** | RH_Triangular part of the inferior frontal gyrus | 2.3 | 1.3 |
| **session 2** | RH_Middle frontal gyrus (F2) | 1.4 | 1.3 |
| **session 2** | RH_Superior frontal gyrus (F1) | 2.8 | 0.0 |
| **session 2** | RH_Long insular gyrus and central sulcus of the insula | 1.4 | 1.3 |
| **session 2** | RH_Short insular gyri | 1.8 | 1.3 |
| **session 2** | RH_Middle occipital gyrus (O2, lateral occipital gyrus) | 1.8 | 1.3 |
| **session 2** | RH_Superior occipital gyrus (O1) | 0.9 | 2.5 |
| **session 2** | RH_Lateral occipito_temporal gyrus (fusiform gyrus, O4_T4) | 1.4 | 3.8 |
| **session 2** | RH_Lingual gyrus, ligual part of the medial occipito_temporal gyrus, (O5) | 1.4 | 0.0 |
| **session 2** | RH_Parahippocampal gyrus, parahippocampal part of the medial occipito_temporal gyrus, (T5) | 1.4 | 2.5 |
| **session 2** | RH_Orbital gyri | 2.3 | 6.3 |
| **session 2** | RH_Angular gyrus | 1.4 | 0.0 |
| **session 2** | RH_Supramarginal gyrus | 2.3 | 1.3 |
| **session 2** | RH_Superior parietal lobule (lateral part of P1) | 4.1 | 0.0 |
| **session 2** | RH_Postcentral gyrus | 3.7 | 2.5 |
| **session 2** | RH_Precentral gyrus | 0.5 | 0.0 |
| **session 2** | RH_Precuneus (medial part of P1) | 3.7 | 0.0 |
| **session 2** | RH_Straight gyrus, Gyrus rectus | 2.8 | 1.3 |
| **session 2** | RH_Subcallosal area, subcallosal gyrus | 2.8 | 0.0 |
| **session 2** | RH_Anterior transverse temporal gyrus (of Heschl) | 5.5 | 1.3 |
| **session 2** | RH_Lateral aspect of the superior temporal gyrus | 0.9 | 0.0 |
| **session 2** | RH_Planum polare of the superior temporal gyrus | 1.4 | 0.0 |
| **session 2** | RH_Planum temporale or temporal plane of the superior temporal gyrus | 1.4 | 2.5 |
| **session 2** | RH_Inferior temporal gyrus (T3) | 2.8 | 2.5 |
| **session 2** | RH_Middle temporal gyrus (T2) | 1.4 | 2.5 |
| **session 2** | RH_Horizontal ramus of the anterior segment of the lateral sulcus (or fissure) | 2.3 | 1.3 |
| **session 2** | RH_Vertical ramus of the anterior segment of the lateral sulcus (or fissure) | 5.0 | 6.3 |
| **session 2** | RH_Posterior ramus (or segment) of the lateral sulcus (or fissure) | 2.3 | 0.0 |
| **session 2** | RH_Occipital pole | 5.5 | 2.5 |
| **session 2** | RH_Temporal pole | 3.2 | 3.8 |
| **session 2** | RH_Calcarine sulcus | 3.2 | 2.5 |
| **session 2** | RH_Central sulcus (Rolandos fissure) | 1.8 | 2.5 |
| **session 2** | RH_Marginal branch (or part) of the cingulate sulcus | 3.2 | 0.0 |
| **session 2** | RH_Anterior segment of the circular sulcus of the insula | 3.7 | 2.5 |
| **session 2** | RH_Inferior segment of the circular sulcus of the insula | 1.4 | 1.3 |
| **session 2** | RH_Superior segment of the circular sulcus of the insula | 1.4 | 1.3 |
| **session 2** | RH_Anterior transverse collateral sulcus | 2.3 | 0.0 |
| **session 2** | RH_Posterior transverse collateral sulcus | 0.9 | 0.0 |
| **session 2** | RH_Inferior frontal sulcus | 2.8 | 5.1 |
| **session 2** | RH_Middle frontal sulcus | 5.0 | 2.5 |
| **session 2** | RH_Superior frontal sulcus | 0.9 | 3.8 |
| **session 2** | RH_Sulcus intermedius primus (of Jensen) | 0.5 | 3.8 |

| | | | |
|---|---|---|---|
| **session 2** | RH_Intraparietal sulcus (interparietal sulcus) and transverse parietal sulci | 0.5 | 0.0 |
| **session 2** | RH_Middle occipital sulcus and lunatus sulcus | 0.9 | 0.0 |
| **session 2** | RH_Superior occipital sulcus and transverse occipital sulcus | 2.3 | 1.3 |
| **session 2** | RH_Anterior occipital sulcus and preoccipital notch (temporo_occipital incisure) | 2.3 | 2.5 |
| **session 2** | RH_Lateral occipito_temporal sulcus | 2.3 | 1.3 |
| **session 2** | RH_Medial occipito_temporal sulcus (collateral sulcus) and lingual sulcus | 2.8 | 2.5 |
| **session 2** | RH_Lateral orbital sulcus | 3.7 | 3.8 |
| **session 2** | RH_Medial orbital sulcus (olfactory sulcus) | 2.3 | 1.3 |
| **session 2** | RH_Orbital sulci (H_shaped sulci) | 3.7 | 2.5 |
| **session 2** | RH_Parieto_occipital sulcus (or fissure) | 2.3 | 2.5 |
| **session 2** | RH_Pericallosal sulcus (S of corpus callosum) | 6.4 | 8.9 |
| **session 2** | RH_Postcentral sulcus | 1.8 | 0.0 |
| **session 2** | RH_Inferior part of the precentral sulcus | 0.9 | 0.0 |
| **session 2** | RH_Superior part of the precentral sulcus | 1.4 | 0.0 |
| **session 2** | RH_Suborbital sulcus (sulcus rostrales, supraorbital sulcus) | 1.8 | 2.5 |
| **session 2** | RH_Subparietal sulcus | 2.8 | 1.3 |
| **session 2** | RH_Inferior temporal sulcus | 1.4 | 3.8 |
| **session 2** | RH_Superior temporal sulcus (parallel sulcus) | 2.8 | 0.0 |
| **session 2** | RH_Transverse temporal sulcus | 1.8 | 0.0 |
| **session 3** | LH_Fronto_marginal gyrus (of Wernicke) and sulcus | 3.6 | 0.0 |
| **session 3** | LH_Inferior occipital gyrus (O3) and sulcus | 0.0 | 0.0 |
| **session 3** | LH_Paracentral lobule and sulcus | 0.0 | 0.0 |
| **session 3** | LH_Subcentral gyrus (central operculum) and sulci | 0.0 | 0.0 |
| **session 3** | LH_Transverse frontopolar gyri and sulci | 1.8 | 0.0 |
| **session 3** | LH_Anterior part of the cingulate gyrus and sulcus (ACC) | 1.8 | 0.0 |
| **session 3** | LH_Middle_anterior part of the cingulate gyrus and sulcus (aMCC) | 0.0 | 0.0 |
| **session 3** | LH_Middle_posterior part of the cingulate gyrus and sulcus (pMCC) | 0.0 | 0.0 |
| **session 3** | LH_Posterior_dorsal part of the cingulate gyrus (dPCC) | 0.0 | 0.0 |
| **session 3** | LH_Posterior_ventral part of the cingulate gyrus (vPCC, isthmus of the cingulate gyrus) | 0.0 | 0.0 |
| **session 3** | LH_Cuneus (O6) | 0.0 | 0.0 |
| **session 3** | LH_Opercular part of the inferior frontal gyrus | 0.0 | 0.0 |
| **session 3** | LH_Orbital part of the inferior frontal gyrus | 0.0 | 0.0 |
| **session 3** | LH_Triangular part of the inferior frontal gyrus | 1.8 | 0.0 |
| **session 3** | LH_Middle frontal gyrus (F2) | 0.0 | 0.0 |
| **session 3** | LH_Superior frontal gyrus (F1) | 0.0 | 0.0 |
| **session 3** | LH_Long insular gyrus and central sulcus of the insula | 0.0 | 0.0 |
| **session 3** | LH_Short insular gyri | 0.0 | 0.0 |
| **session 3** | LH_Middle occipital gyrus (O2, lateral occipital gyrus) | 0.0 | 0.0 |
| **session 3** | LH_Superior occipital gyrus (O1) | 1.8 | 0.0 |
| **session 3** | LH_Lateral occipito_temporal gyrus (fusiform gyrus, O4_T4) | 0.0 | 0.0 |
| **session 3** | LH_Lingual gyrus, ligual part of the medial occipito_temporal gyrus, (O5) | 1.8 | 0.0 |
| **session 3** | LH_Parahippocampal gyrus, parahippocampal part of the medial occipito_temporal gyrus, (T5) | 0.0 | 0.0 |

| | | | |
|---|---|---|---|
| **session 3** | LH_Orbital gyri | 0.0 | 0.0 |
| **session 3** | LH_Angular gyrus | 0.0 | 0.0 |
| **session 3** | LH_Supramarginal gyrus | 1.8 | 0.0 |
| **session 3** | LH_Superior parietal lobule (lateral part of P1) | 0.0 | 0.0 |
| **session 3** | LH_Postcentral gyrus | 0.0 | 0.0 |
| **session 3** | LH_Precentral gyrus | 0.0 | 0.0 |
| **session 3** | LH_Precuneus (medial part of P1) | 0.0 | 0.0 |
| **session 3** | LH_Straight gyrus, Gyrus rectus | 0.0 | 0.0 |
| **session 3** | LH_Subcallosal area, subcallosal gyrus | 0.0 | 9.1 |
| **session 3** | LH_Anterior transverse temporal gyrus (of Heschl) | 1.8 | 0.0 |
| **session 3** | LH_Lateral aspect of the superior temporal gyrus | 1.8 | 0.0 |
| **session 3** | LH_Planum polare of the superior temporal gyrus | 3.6 | 0.0 |
| **session 3** | LH_Planum temporale or temporal plane of the superior temporal gyrus | 0.0 | 0.0 |
| **session 3** | LH_Inferior temporal gyrus (T3) | 0.0 | 9.1 |
| **session 3** | LH_Middle temporal gyrus (T2) | 0.0 | 0.0 |
| **session 3** | LH_Horizontal ramus of the anterior segment of the lateral sulcus (or fissure) | 3.6 | 0.0 |
| **session 3** | LH_Vertical ramus of the anterior segment of the lateral sulcus (or fissure) | 1.8 | 0.0 |
| **session 3** | LH_Posterior ramus (or segment) of the lateral sulcus (or fissure) | 1.8 | 0.0 |
| **session 3** | LH_Occipital pole | 3.6 | 0.0 |
| **session 3** | LH_Temporal pole | 0.0 | 0.0 |
| **session 3** | LH_Calcarine sulcus | 1.8 | 0.0 |
| **session 3** | LH_Central sulcus (Rolandos fissure) | 1.8 | 0.0 |
| **session 3** | LH_Marginal branch (or part) of the cingulate sulcus | 3.6 | 0.0 |
| **session 3** | LH_Anterior segment of the circular sulcus of the insula | 0.0 | 0.0 |
| **session 3** | LH_Inferior segment of the circular sulcus of the insula | 3.6 | 0.0 |
| **session 3** | LH_Superior segment of the circular sulcus of the insula | 0.0 | 0.0 |
| **session 3** | LH_Anterior transverse collateral sulcus | 3.6 | 18.2 |
| **session 3** | LH_Posterior transverse collateral sulcus | 3.6 | 9.1 |
| **session 3** | LH_Inferior frontal sulcus | 0.0 | 0.0 |
| **session 3** | LH_Middle frontal sulcus | 0.0 | 0.0 |
| **session 3** | LH_Superior frontal sulcus | 0.0 | 0.0 |
| **session 3** | LH_Sulcus intermedius primus (of Jensen) | 5.5 | 0.0 |
| **session 3** | LH_Intraparietal sulcus (interparietal sulcus) and transverse parietal sulci | 0.0 | 0.0 |
| **session 3** | LH_Middle occipital sulcus and lunatus sulcus | 0.0 | 0.0 |
| **session 3** | LH_Superior occipital sulcus and transverse occipital sulcus | 0.0 | 0.0 |
| **session 3** | LH_Anterior occipital sulcus and preoccipital notch (temporo_occipital incisure) | 0.0 | 0.0 |
| **session 3** | LH_Lateral occipito_temporal sulcus | 0.0 | 0.0 |
| **session 3** | LH_Medial occipito_temporal sulcus (collateral sulcus) and lingual sulcus | 1.8 | 0.0 |
| **session 3** | LH_Lateral orbital sulcus | 0.0 | 0.0 |
| **session 3** | LH_Medial orbital sulcus (olfactory sulcus) | 1.8 | 0.0 |
| **session 3** | LH_Orbital sulci (H_shaped sulci) | 1.8 | 0.0 |
| **session 3** | LH_Parieto_occipital sulcus (or fissure) | 1.8 | 0.0 |

| | | | |
|---|---|---|---|
| **session 3** | LH_Pericallosal sulcus (S of corpus callosum) | 0.0 | 0.0 |
| **session 3** | LH_Postcentral sulcus | 0.0 | 0.0 |
| **session 3** | LH_Inferior part of the precentral sulcus | 3.6 | 0.0 |
| **session 3** | LH_Superior part of the precentral sulcus | 0.0 | 0.0 |
| **session 3** | LH_Suborbital sulcus (sulcus rostrales, supraorbital sulcus) | 0.0 | 0.0 |
| **session 3** | LH_Subparietal sulcus | 3.6 | 0.0 |
| **session 3** | LH_Inferior temporal sulcus | 0.0 | 0.0 |
| **session 3** | LH_Superior temporal sulcus (parallel sulcus) | 0.0 | 0.0 |
| **session 3** | LH_Transverse temporal sulcus | 0.0 | 0.0 |
| **session 3** | RH_Fronto_marginal gyrus (of Wernicke) and sulcus | 0.0 | 0.0 |
| **session 3** | RH_Inferior occipital gyrus (O3) and sulcus | 1.8 | 0.0 |
| **session 3** | RH_Paracentral lobule and sulcus | 1.8 | 0.0 |
| **session 3** | RH_Subcentral gyrus (central operculum) and sulci | 0.0 | 0.0 |
| **session 3** | RH_Transverse frontopolar gyri and sulci | 0.0 | 0.0 |
| **session 3** | RH_Anterior part of the cingulate gyrus and sulcus (ACC) | 0.0 | 0.0 |
| **session 3** | RH_Middle_anterior part of the cingulate gyrus and sulcus (aMCC) | 1.8 | 0.0 |
| **session 3** | RH_Middle_posterior part of the cingulate gyrus and sulcus (pMCC) | 1.8 | 0.0 |
| **session 3** | RH_Posterior_dorsal part of the cingulate gyrus (dPCC) | 0.0 | 0.0 |
| **session 3** | RH_Posterior_ventral part of the cingulate gyrus (vPCC, isthmus of the cingulate gyrus) | 1.8 | 0.0 |
| **session 3** | RH_Cuneus (O6) | 0.0 | 0.0 |
| **session 3** | RH_Opercular part of the inferior frontal gyrus | 1.8 | 0.0 |
| **session 3** | RH_Orbital part of the inferior frontal gyrus | 1.8 | 0.0 |
| **session 3** | RH_Triangular part of the inferior frontal gyrus | 0.0 | 0.0 |
| **session 3** | RH_Middle frontal gyrus (F2) | 0.0 | 0.0 |
| **session 3** | RH_Superior frontal gyrus (F1) | 1.8 | 0.0 |
| **session 3** | RH_Long insular gyrus and central sulcus of the insula | 0.0 | 0.0 |
| **session 3** | RH_Short insular gyri | 0.0 | 0.0 |
| **session 3** | RH_Middle occipital gyrus (O2, lateral occipital gyrus) | 0.0 | 0.0 |
| **session 3** | RH_Superior occipital gyrus (O1) | 0.0 | 0.0 |
| **session 3** | RH_Lateral occipito_temporal gyrus (fusiform gyrus, O4_T4) | 0.0 | 0.0 |
| **session 3** | RH_Lingual gyrus, ligual part of the medial occipito_temporal gyrus, (O5) | 1.8 | 0.0 |
| **session 3** | RH_Parahippocampal gyrus, parahippocampal part of the medial occipito_temporal gyrus, (T5) | 1.8 | 9.1 |
| **session 3** | RH_Orbital gyri | 0.0 | 0.0 |
| **session 3** | RH_Angular gyrus | 0.0 | 0.0 |
| **session 3** | RH_Supramarginal gyrus | 0.0 | 0.0 |
| **session 3** | RH_Superior parietal lobule (lateral part of P1) | 0.0 | 0.0 |
| **session 3** | RH_Postcentral gyrus | 1.8 | 0.0 |
| **session 3** | RH_Precentral gyrus | 0.0 | 0.0 |
| **session 3** | RH_Precuneus (medial part of P1) | 1.8 | 0.0 |
| **session 3** | RH_Straight gyrus, Gyrus rectus | 1.8 | 0.0 |
| **session 3** | RH_Subcallosal area, subcallosal gyrus | 0.0 | 27.3 |
| **session 3** | RH_Anterior transverse temporal gyrus (of Heschl) | 7.3 | 0.0 |
| **session 3** | RH_Lateral aspect of the superior temporal gyrus | 0.0 | 0.0 |
| **session 3** | RH_Planum polare of the superior temporal gyrus | 0.0 | 0.0 |

| | | | |
|---|---|---|---|
| **session 3** | RH_Planum temporale or temporal plane of the superior temporal gyrus | 1.8 | 0.0 |
| **session 3** | RH_Inferior temporal gyrus (T3) | 0.0 | 0.0 |
| **session 3** | RH_Middle temporal gyrus (T2) | 1.8 | 0.0 |
| **session 3** | RH_Horizontal ramus of the anterior segment of the lateral sulcus (or fissure) | 0.0 | 9.1 |
| **session 3** | RH_Vertical ramus of the anterior segment of the lateral sulcus (or fissure) | 5.5 | 0.0 |
| **session 3** | RH_Posterior ramus (or segment) of the lateral sulcus (or fissure) | 3.6 | 0.0 |
| **session 3** | RH_Occipital pole | 1.8 | 9.1 |
| **session 3** | RH_Temporal pole | 0.0 | 0.0 |
| **session 3** | RH_Calcarine sulcus | 5.5 | 0.0 |
| **session 3** | RH_Central sulcus (Rolandos fissure) | 3.6 | 0.0 |
| **session 3** | RH_Marginal branch (or part) of the cingulate sulcus | 1.8 | 0.0 |
| **session 3** | RH_Anterior segment of the circular sulcus of the insula | 1.8 | 0.0 |
| **session 3** | RH_Inferior segment of the circular sulcus of the insula | 1.8 | 0.0 |
| **session 3** | RH_Superior segment of the circular sulcus of the insula | 1.8 | 0.0 |
| **session 3** | RH_Anterior transverse collateral sulcus | 1.8 | 0.0 |
| **session 3** | RH_Posterior transverse collateral sulcus | 0.0 | 0.0 |
| **session 3** | RH_Inferior frontal sulcus | 3.6 | 0.0 |
| **session 3** | RH_Middle frontal sulcus | 0.0 | 0.0 |
| **session 3** | RH_Superior frontal sulcus | 0.0 | 0.0 |
| **session 3** | RH_Sulcus intermedius primus (of Jensen) | 0.0 | 9.1 |
| **session 3** | RH_Intraparietal sulcus (interparietal sulcus) and transverse parietal sulci | 1.8 | 0.0 |
| **session 3** | RH_Middle occipital sulcus and lunatus sulcus | 0.0 | 0.0 |
| **session 3** | RH_Superior occipital sulcus and transverse occipital sulcus | 1.8 | 0.0 |
| **session 3** | RH_Anterior occipital sulcus and preoccipital notch (temporo_occipital incisure) | 0.0 | 0.0 |
| **session 3** | RH_Lateral occipito_temporal sulcus | 5.5 | 0.0 |
| **session 3** | RH_Medial occipito_temporal sulcus (collateral sulcus) and lingual sulcus | 1.8 | 0.0 |
| **session 3** | RH_Lateral orbital sulcus | 3.6 | 0.0 |
| **session 3** | RH_Medial orbital sulcus (olfactory sulcus) | 0.0 | 0.0 |
| **session 3** | RH_Orbital sulci (H_shaped sulci) | 3.6 | 0.0 |
| **session 3** | RH_Parieto_occipital sulcus (or fissure) | 0.0 | 0.0 |
| **session 3** | RH_Pericallosal sulcus (S of corpus callosum) | 5.5 | 0.0 |
| **session 3** | RH_Postcentral sulcus | 0.0 | 0.0 |
| **session 3** | RH_Inferior part of the precentral sulcus | 0.0 | 0.0 |
| **session 3** | RH_Superior part of the precentral sulcus | 3.6 | 0.0 |
| **session 3** | RH_Suborbital sulcus (sulcus rostrales, supraorbital sulcus) | 1.8 | 0.0 |
| **session 3** | RH_Subparietal sulcus | 1.8 | 0.0 |
| **session 3** | RH_Inferior temporal sulcus | 0.0 | 0.0 |
| **session 3** | RH_Superior temporal sulcus (parallel sulcus) | 1.8 | 0.0 |
| **session 3** | RH_Transverse temporal sulcus | 1.8 | 0.0 |

**Supplementary Table 5:** Percentage (%) of extreme negative deviation overlap by ROI, by time point, for controls and patients with schizophrenia.

| Session | ROI | Controls | Patients |
|---|---|---|---|
| **session 1** | LH_Fronto_marginal gyrus (of Wernicke) and sulcus | 4.1 | 3.8 |
| **session 1** | LH_Inferior occipital gyrus (O3) and sulcus | 1.8 | 12.7 |
| **session 1** | LH_Paracentral lobule and sulcus | 1.4 | 1.3 |
| **session 1** | LH_Subcentral gyrus (central operculum) and sulci | 0.5 | 2.5 |
| **session 1** | LH_Transverse frontopolar gyri and sulci | 1.4 | 5.1 |
| **session 1** | LH_Anterior part of the cingulate gyrus and sulcus (ACC) | 3.7 | 10.1 |
| **session 1** | LH_Middle_anterior part of the cingulate gyrus and sulcus (aMCC) | 2.3 | 5.1 |
| **session 1** | LH_Middle_posterior part of the cingulate gyrus and sulcus (pMCC) | 1.8 | 7.6 |
| **session 1** | LH_Posterior_dorsal part of the cingulate gyrus (dPCC) | 2.3 | 3.8 |
| **session 1** | LH_Posterior_ventral part of the cingulate gyrus (vPCC, isthmus of the cingulate gyrus) | 0.9 | 2.5 |
| **session 1** | LH_Cuneus (O6) | 0.5 | 1.3 |
| **session 1** | LH_Opercular part of the inferior frontal gyrus | 5.0 | 7.6 |
| **session 1** | LH_Orbital part of the inferior frontal gyrus | 3.2 | 2.5 |
| **session 1** | LH_Triangular part of the inferior frontal gyrus | 1.4 | 3.8 |
| **session 1** | LH_Middle frontal gyrus (F2) | 1.4 | 3.8 |
| **session 1** | LH_Superior frontal gyrus (F1) | 2.3 | 5.1 |
| **session 1** | LH_Long insular gyrus and central sulcus of the insula | 3.7 | 2.5 |
| **session 1** | LH_Short insular gyri | 3.2 | 1.3 |
| **session 1** | LH_Middle occipital gyrus (O2, lateral occipital gyrus) | 2.8 | 12.7 |
| **session 1** | LH_Superior occipital gyrus (O1) | 2.3 | 3.8 |
| **session 1** | LH_Lateral occipito_temporal gyrus (fusiform gyrus, O4_T4) | 5.0 | 13.9 |
| **session 1** | LH_Lingual gyrus, ligual part of the medial occipito_temporal gyrus, (O5) | 1.4 | 2.5 |
| **session 1** | LH_Parahippocampal gyrus, parahippocampal part of the medial occipito_temporal gyrus, (T5) | 2.3 | 6.3 |
| **session 1** | LH_Orbital gyri | 3.2 | 6.3 |
| **session 1** | LH_Angular gyrus | 5.0 | 6.3 |
| **session 1** | LH_Supramarginal gyrus | 4.1 | 7.6 |
| **session 1** | LH_Superior parietal lobule (lateral part of P1) | 1.4 | 10.1 |
| **session 1** | LH_Postcentral gyrus | 3.2 | 7.6 |
| **session 1** | LH_Precentral gyrus | 3.2 | 6.3 |
| **session 1** | LH_Precuneus (medial part of P1) | 4.1 | 8.9 |
| **session 1** | LH_Straight gyrus, Gyrus rectus | 4.1 | 5.1 |
| **session 1** | LH_Subcallosal area, subcallosal gyrus | 0.9 | 5.1 |
| **session 1** | LH_Anterior transverse temporal gyrus (of Heschl) | 3.7 | 7.6 |
| **session 1** | LH_Lateral aspect of the superior temporal gyrus | 1.8 | 10.1 |
| **session 1** | LH_Planum polare of the superior temporal gyrus | 1.8 | 6.3 |
| **session 1** | LH_Planum temporale or temporal plane of the superior temporal gyrus | 4.1 | 12.7 |
| **session 1** | LH_Inferior temporal gyrus (T3) | 3.2 | 10.1 |
| **session 1** | LH_Middle temporal gyrus (T2) | 2.3 | 7.6 |

| | | | |
|---|---|---|---|
| **session 1** | LH_Horizontal ramus of the anterior segment of the lateral sulcus (or fissure) | 1.4 | 1.3 |
| **session 1** | LH_Vertical ramus of the anterior segment of the lateral sulcus (or fissure) | 1.4 | 3.8 |
| **session 1** | LH_Posterior ramus (or segment) of the lateral sulcus (or fissure) | 0.9 | 2.5 |
| **session 1** | LH_Occipital pole | 2.3 | 2.5 |
| **session 1** | LH_Temporal pole | 3.7 | 1.3 |
| **session 1** | LH_Calcarine sulcus | 1.4 | 3.8 |
| **session 1** | LH_Central sulcus (Rolandos fissure) | 1.8 | 3.8 |
| **session 1** | LH_Marginal branch (or part) of the cingulate sulcus | 1.8 | 6.3 |
| **session 1** | LH_Anterior segment of the circular sulcus of the insula | 3.7 | 5.1 |
| **session 1** | LH_Inferior segment of the circular sulcus of the insula | 3.7 | 17.7 |
| **session 1** | LH_Superior segment of the circular sulcus of the insula | 3.2 | 7.6 |
| **session 1** | LH_Anterior transverse collateral sulcus | 2.8 | 5.1 |
| **session 1** | LH_Posterior transverse collateral sulcus | 1.4 | 6.3 |
| **session 1** | LH_Inferior frontal sulcus | 5.0 | 12.7 |
| **session 1** | LH_Middle frontal sulcus | 3.7 | 8.9 |
| **session 1** | LH_Superior frontal sulcus | 2.8 | 16.5 |
| **session 1** | LH_Sulcus intermedius primus (of Jensen) | 1.4 | 2.5 |
| **session 1** | LH_Intraparietal sulcus (interparietal sulcus) and transverse parietal sulci | 3.7 | 15.2 |
| **session 1** | LH_Middle occipital sulcus and lunatus sulcus | 0.9 | 3.8 |
| **session 1** | LH_Superior occipital sulcus and transverse occipital sulcus | 1.4 | 12.7 |
| **session 1** | LH_Anterior occipital sulcus and preoccipital notch (temporo_occipital incisure) | 2.3 | 11.4 |
| **session 1** | LH_Lateral occipito_temporal sulcus | 1.8 | 10.1 |
| **session 1** | LH_Medial occipito_temporal sulcus (collateral sulcus) and lingual sulcus | 0.0 | 8.9 |
| **session 1** | LH_Lateral orbital sulcus | 1.8 | 1.3 |
| **session 1** | LH_Medial orbital sulcus (olfactory sulcus) | 3.2 | 3.8 |
| **session 1** | LH_Orbital sulci (H_shaped sulci) | 4.1 | 6.3 |
| **session 1** | LH_Parieto_occipital sulcus (or fissure) | 3.7 | 6.3 |
| **session 1** | LH_Pericallosal sulcus (S of corpus callosum) | 0.0 | 0.0 |
| **session 1** | LH_Postcentral sulcus | 2.8 | 5.1 |
| **session 1** | LH_Inferior part of the precentral sulcus | 6.4 | 15.2 |
| **session 1** | LH_Superior part of the precentral sulcus | 4.6 | 8.9 |
| **session 1** | LH_Suborbital sulcus (sulcus rostrales, supraorbital sulcus) | 3.2 | 5.1 |
| **session 1** | LH_Subparietal sulcus | 2.8 | 12.7 |
| **session 1** | LH_Inferior temporal sulcus | 7.3 | 13.9 |
| **session 1** | LH_Superior temporal sulcus (parallel sulcus) | 4.1 | 11.4 |
| **session 1** | LH_Transverse temporal sulcus | 0.5 | 3.8 |
| **session 1** | RH_Fronto_marginal gyrus (of Wernicke) and sulcus | 4.1 | 3.8 |
| **session 1** | RH_Inferior occipital gyrus (O3) and sulcus | 0.9 | 3.8 |
| **session 1** | RH_Paracentral lobule and sulcus | 1.8 | 2.5 |
| **session 1** | RH_Subcentral gyrus (central operculum) and sulci | 0.5 | 3.8 |
| **session 1** | RH_Transverse frontopolar gyri and sulci | 1.4 | 1.3 |
| **session 1** | RH_Anterior part of the cingulate gyrus and sulcus (ACC) | 2.3 | 11.4 |
| **session 1** | RH_Middle_anterior part of the cingulate gyrus and sulcus (aMCC) | 2.8 | 5.1 |

| | | | |
|---|---|---|---|
| **session 1** | RH_Middle_posterior part of the cingulate gyrus and sulcus (pMCC) | 2.8 | 6.3 |
| **session 1** | RH_Posterior_dorsal part of the cingulate gyrus (dPCC) | 1.4 | 6.3 |
| **session 1** | RH_Posterior_ventral part of the cingulate gyrus (vPCC, isthmus of the cingulate gyrus) | 2.8 | 0.0 |
| **session 1** | RH_Cuneus (O6) | 1.4 | 2.5 |
| **session 1** | RH_Opercular part of the inferior frontal gyrus | 2.3 | 15.2 |
| **session 1** | RH_Orbital part of the inferior frontal gyrus | 2.8 | 6.3 |
| **session 1** | RH_Triangular part of the inferior frontal gyrus | 1.8 | 6.3 |
| **session 1** | RH_Middle frontal gyrus (F2) | 2.8 | 5.1 |
| **session 1** | RH_Superior frontal gyrus (F1) | 1.8 | 6.3 |
| **session 1** | RH_Long insular gyrus and central sulcus of the insula | 3.2 | 2.5 |
| **session 1** | RH_Short insular gyri | 2.3 | 5.1 |
| **session 1** | RH_Middle occipital gyrus (O2, lateral occipital gyrus) | 3.2 | 10.1 |
| **session 1** | RH_Superior occipital gyrus (O1) | 5.0 | 10.1 |
| **session 1** | RH_Lateral occipito_temporal gyrus (fusiform gyrus, O4_T4) | 4.6 | 13.9 |
| **session 1** | RH_Lingual gyrus, ligual part of the medial occipito_temporal gyrus, (O5) | 3.2 | 2.5 |
| **session 1** | RH_Parahippocampal gyrus, parahippocampal part of the medial occipito_temporal gyrus, (T5) | 1.8 | 5.1 |
| **session 1** | RH_Orbital gyri | 1.4 | 6.3 |
| **session 1** | RH_Angular gyrus | 2.8 | 6.3 |
| **session 1** | RH_Supramarginal gyrus | 2.3 | 12.7 |
| **session 1** | RH_Superior parietal lobule (lateral part of P1) | 2.8 | 2.5 |
| **session 1** | RH_Postcentral gyrus | 2.3 | 5.1 |
| **session 1** | RH_Precentral gyrus | 4.6 | 8.9 |
| **session 1** | RH_Precuneus (medial part of P1) | 3.2 | 6.3 |
| **session 1** | RH_Straight gyrus, Gyrus rectus | 4.1 | 3.8 |
| **session 1** | RH_Subcallosal area, subcallosal gyrus | 0.5 | 1.3 |
| **session 1** | RH_Anterior transverse temporal gyrus (of Heschl) | 2.8 | 2.5 |
| **session 1** | RH_Lateral aspect of the superior temporal gyrus | 0.9 | 13.9 |
| **session 1** | RH_Planum polare of the superior temporal gyrus | 1.8 | 7.6 |
| **session 1** | RH_Planum temporale or temporal plane of the superior temporal gyrus | 1.4 | 5.1 |
| **session 1** | RH_Inferior temporal gyrus (T3) | 1.4 | 5.1 |
| **session 1** | RH_Middle temporal gyrus (T2) | 1.8 | 10.1 |
| **session 1** | RH_Horizontal ramus of the anterior segment of the lateral sulcus (or fissure) | 0.5 | 6.3 |
| **session 1** | RH_Vertical ramus of the anterior segment of the lateral sulcus (or fissure) | 1.4 | 5.1 |
| **session 1** | RH_Posterior ramus (or segment) of the lateral sulcus (or fissure) | 1.4 | 10.1 |
| **session 1** | RH_Occipital pole | 1.8 | 2.5 |
| **session 1** | RH_Temporal pole | 5.5 | 10.1 |
| **session 1** | RH_Calcarine sulcus | 1.8 | 3.8 |
| **session 1** | RH_Central sulcus (Rolandos fissure) | 1.8 | 2.5 |
| **session 1** | RH_Marginal branch (or part) of the cingulate sulcus | 1.4 | 11.4 |
| **session 1** | RH_Anterior segment of the circular sulcus of the insula | 2.8 | 1.3 |
| **session 1** | RH_Inferior segment of the circular sulcus of the insula | 0.9 | 10.1 |
| **session 1** | RH_Superior segment of the circular sulcus of the insula | 2.8 | 7.6 |

| | | | |
|---|---|---|---|
| **session 1** | RH_Anterior transverse collateral sulcus | 2.8 | 6.3 |
| **session 1** | RH_Posterior transverse collateral sulcus | 1.4 | 3.8 |
| **session 1** | RH_Inferior frontal sulcus | 3.2 | 17.7 |
| **session 1** | RH_Middle frontal sulcus | 0.9 | 1.3 |
| **session 1** | RH_Superior frontal sulcus | 2.8 | 10.1 |
| **session 1** | RH_Sulcus intermedius primus (of Jensen) | 3.2 | 7.6 |
| **session 1** | RH_Intraparietal sulcus (interparietal sulcus) and transverse parietal sulci | 1.8 | 7.6 |
| **session 1** | RH_Middle occipital sulcus and lunatus sulcus | 0.9 | 3.8 |
| **session 1** | RH_Superior occipital sulcus and transverse occipital sulcus | 3.7 | 11.4 |
| **session 1** | RH_Anterior occipital sulcus and preoccipital notch (temporo_occipital incisure) | 5.0 | 12.7 |
| **session 1** | RH_Lateral occipito_temporal sulcus | 2.8 | 11.4 |
| **session 1** | RH_Medial occipito_temporal sulcus (collateral sulcus) and lingual sulcus | 2.8 | 3.8 |
| **session 1** | RH_Lateral orbital sulcus | 0.5 | 6.3 |
| **session 1** | RH_Medial orbital sulcus (olfactory sulcus) | 4.1 | 2.5 |
| **session 1** | RH_Orbital sulci (H_shaped sulci) | 3.2 | 6.3 |
| **session 1** | RH_Parieto_occipital sulcus (or fissure) | 3.7 | 10.1 |
| **session 1** | RH_Pericallosal sulcus (S of corpus callosum) | 0.0 | 0.0 |
| **session 1** | RH_Postcentral sulcus | 1.8 | 5.1 |
| **session 1** | RH_Inferior part of the precentral sulcus | 4.6 | 12.7 |
| **session 1** | RH_Superior part of the precentral sulcus | 2.3 | 7.6 |
| **session 1** | RH_Suborbital sulcus (sulcus rostrales, supraorbital sulcus) | 2.8 | 1.3 |
| **session 1** | RH_Subparietal sulcus | 6.0 | 17.7 |
| **session 1** | RH_Inferior temporal sulcus | 5.0 | 11.4 |
| **session 1** | RH_Superior temporal sulcus (parallel sulcus) | 5.0 | 20.3 |
| **session 1** | RH_Transverse temporal sulcus | 3.2 | 7.6 |
| **session 2** | LH_Fronto_marginal gyrus (of Wernicke) and sulcus | 1.8 | 2.5 |
| **session 2** | LH_Inferior occipital gyrus (O3) and sulcus | 3.2 | 6.3 |
| **session 2** | LH_Paracentral lobule and sulcus | 1.8 | 2.5 |
| **session 2** | LH_Subcentral gyrus (central operculum) and sulci | 0.9 | 6.3 |
| **session 2** | LH_Transverse frontopolar gyri and sulci | 0.5 | 8.9 |
| **session 2** | LH_Anterior part of the cingulate gyrus and sulcus (ACC) | 3.2 | 6.3 |
| **session 2** | LH_Middle_anterior part of the cingulate gyrus and sulcus (aMCC) | 1.8 | 2.5 |
| **session 2** | LH_Middle_posterior part of the cingulate gyrus and sulcus (pMCC) | 2.8 | 6.3 |
| **session 2** | LH_Posterior_dorsal part of the cingulate gyrus (dPCC) | 1.4 | 7.6 |
| **session 2** | LH_Posterior_ventral part of the cingulate gyrus (vPCC, isthmus of the cingulate gyrus) | 1.4 | 2.5 |
| **session 2** | LH_Cuneus (O6) | 0.9 | 3.8 |
| **session 2** | LH_Opercular part of the inferior frontal gyrus | 3.2 | 6.3 |
| **session 2** | LH_Orbital part of the inferior frontal gyrus | 0.9 | 5.1 |
| **session 2** | LH_Triangular part of the inferior frontal gyrus | 1.4 | 1.3 |
| **session 2** | LH_Middle frontal gyrus (F2) | 1.8 | 5.1 |
| **session 2** | LH_Superior frontal gyrus (F1) | 0.9 | 3.8 |
| **session 2** | LH_Long insular gyrus and central sulcus of the insula | 3.2 | 2.5 |
| **session 2** | LH_Short insular gyri | 2.8 | 3.8 |

| | | | |
|---|---|---|---|
| **session 2** | LH_Middle occipital gyrus (O2, lateral occipital gyrus) | 3.7 | 7.6 |
| **session 2** | LH_Superior occipital gyrus (O1) | 1.4 | 5.1 |
| **session 2** | LH_Lateral occipito_temporal gyrus (fusiform gyrus, O4_T4) | 5.0 | 10.1 |
| **session 2** | LH_Lingual gyrus, ligual part of the medial occipito_temporal gyrus, (O5) | 1.8 | 2.5 |
| **session 2** | LH_Parahippocampal gyrus, parahippocampal part of the medial occipito_temporal gyrus, (T5) | 2.3 | 7.6 |
| **session 2** | LH_Orbital gyri | 1.8 | 6.3 |
| **session 2** | LH_Angular gyrus | 4.6 | 3.8 |
| **session 2** | LH_Supramarginal gyrus | 4.6 | 10.1 |
| **session 2** | LH_Superior parietal lobule (lateral part of P1) | 4.1 | 6.3 |
| **session 2** | LH_Postcentral gyrus | 1.8 | 6.3 |
| **session 2** | LH_Precentral gyrus | 2.8 | 5.1 |
| **session 2** | LH_Precuneus (medial part of P1) | 3.7 | 7.6 |
| **session 2** | LH_Straight gyrus, Gyrus rectus | 2.3 | 3.8 |
| **session 2** | LH_Subcallosal area, subcallosal gyrus | 1.4 | 1.3 |
| **session 2** | LH_Anterior transverse temporal gyrus (of Heschl) | 3.7 | 7.6 |
| **session 2** | LH_Lateral aspect of the superior temporal gyrus | 1.8 | 6.3 |
| **session 2** | LH_Planum polare of the superior temporal gyrus | 1.8 | 5.1 |
| **session 2** | LH_Planum temporale or temporal plane of the superior temporal gyrus | 1.8 | 7.6 |
| **session 2** | LH_Inferior temporal gyrus (T3) | 3.2 | 6.3 |
| **session 2** | LH_Middle temporal gyrus (T2) | 3.7 | 7.6 |
| **session 2** | LH_Horizontal ramus of the anterior segment of the lateral sulcus (or fissure) | 0.0 | 2.5 |
| **session 2** | LH_Vertical ramus of the anterior segment of the lateral sulcus (or fissure) | 0.0 | 0.0 |
| **session 2** | LH_Posterior ramus (or segment) of the lateral sulcus (or fissure) | 0.5 | 5.1 |
| **session 2** | LH_Occipital pole | 2.8 | 1.3 |
| **session 2** | LH_Temporal pole | 1.8 | 5.1 |
| **session 2** | LH_Calcarine sulcus | 1.4 | 6.3 |
| **session 2** | LH_Central sulcus (Rolandos fissure) | 2.8 | 2.5 |
| **session 2** | LH_Marginal branch (or part) of the cingulate sulcus | 3.2 | 6.3 |
| **session 2** | LH_Anterior segment of the circular sulcus of the insula | 2.3 | 3.8 |
| **session 2** | LH_Inferior segment of the circular sulcus of the insula | 3.7 | 7.6 |
| **session 2** | LH_Superior segment of the circular sulcus of the insula | 1.4 | 8.9 |
| **session 2** | LH_Anterior transverse collateral sulcus | 1.8 | 2.5 |
| **session 2** | LH_Posterior transverse collateral sulcus | 1.4 | 3.8 |
| **session 2** | LH_Inferior frontal sulcus | 2.3 | 3.8 |
| **session 2** | LH_Middle frontal sulcus | 1.8 | 8.9 |
| **session 2** | LH_Superior frontal sulcus | 1.4 | 7.6 |
| **session 2** | LH_Sulcus intermedius primus (of Jensen) | 1.8 | 2.5 |
| **session 2** | LH_Intraparietal sulcus (interparietal sulcus) and transverse parietal sulci | 3.2 | 13.9 |
| **session 2** | LH_Middle occipital sulcus and lunatus sulcus | 2.8 | 3.8 |
| **session 2** | LH_Superior occipital sulcus and transverse occipital sulcus | 2.8 | 5.1 |
| **session 2** | LH_Anterior occipital sulcus and preoccipital notch (temporo_occipital incisure) | 3.7 | 8.9 |
| **session 2** | LH_Lateral occipito_temporal sulcus | 0.5 | 3.8 |

| | | | |
|---|---|---|---|
| **session 2** | LH_Medial occipito_temporal sulcus (collateral sulcus) and lingual sulcus | 1.4 | 5.1 |
| **session 2** | LH_Lateral orbital sulcus | 0.5 | 2.5 |
| **session 2** | LH_Medial orbital sulcus (olfactory sulcus) | 0.5 | 3.8 |
| **session 2** | LH_Orbital sulci (H_shaped sulci) | 0.9 | 3.8 |
| **session 2** | LH_Parieto_occipital sulcus (or fissure) | 2.8 | 6.3 |
| **session 2** | LH_Pericallosal sulcus (S of corpus callosum) | 0.5 | 0.0 |
| **session 2** | LH_Postcentral sulcus | 2.3 | 3.8 |
| **session 2** | LH_Inferior part of the precentral sulcus | 3.7 | 8.9 |
| **session 2** | LH_Superior part of the precentral sulcus | 4.1 | 7.6 |
| **session 2** | LH_Suborbital sulcus (sulcus rostrales, supraorbital sulcus) | 2.3 | 2.5 |
| **session 2** | LH_Subparietal sulcus | 3.2 | 8.9 |
| **session 2** | LH_Inferior temporal sulcus | 6.9 | 7.6 |
| **session 2** | LH_Superior temporal sulcus (parallel sulcus) | 4.1 | 7.6 |
| **session 2** | LH_Transverse temporal sulcus | 2.3 | 5.1 |
| **session 2** | RH_Fronto_marginal gyrus (of Wernicke) and sulcus | 3.7 | 2.5 |
| **session 2** | RH_Inferior occipital gyrus (O3) and sulcus | 1.4 | 1.3 |
| **session 2** | RH_Paracentral lobule and sulcus | 0.9 | 6.3 |
| **session 2** | RH_Subcentral gyrus (central operculum) and sulci | 0.9 | 2.5 |
| **session 2** | RH_Transverse frontopolar gyri and sulci | 0.5 | 2.5 |
| **session 2** | RH_Anterior part of the cingulate gyrus and sulcus (ACC) | 1.8 | 3.8 |
| **session 2** | RH_Middle_anterior part of the cingulate gyrus and sulcus (aMCC) | 0.9 | 3.8 |
| **session 2** | RH_Middle_posterior part of the cingulate gyrus and sulcus (pMCC) | 1.4 | 3.8 |
| **session 2** | RH_Posterior_dorsal part of the cingulate gyrus (dPCC) | 3.2 | 3.8 |
| **session 2** | RH_Posterior_ventral part of the cingulate gyrus (vPCC, isthmus of the cingulate gyrus) | 1.8 | 2.5 |
| **session 2** | RH_Cuneus (O6) | 2.3 | 3.8 |
| **session 2** | RH_Opercular part of the inferior frontal gyrus | 1.4 | 6.3 |
| **session 2** | RH_Orbital part of the inferior frontal gyrus | 1.8 | 5.1 |
| **session 2** | RH_Triangular part of the inferior frontal gyrus | 1.8 | 2.5 |
| **session 2** | RH_Middle frontal gyrus (F2) | 1.4 | 6.3 |
| **session 2** | RH_Superior frontal gyrus (F1) | 1.8 | 3.8 |
| **session 2** | RH_Long insular gyrus and central sulcus of the insula | 2.8 | 5.1 |
| **session 2** | RH_Short insular gyri | 3.2 | 6.3 |
| **session 2** | RH_Middle occipital gyrus (O2, lateral occipital gyrus) | 2.8 | 6.3 |
| **session 2** | RH_Superior occipital gyrus (O1) | 3.2 | 5.1 |
| **session 2** | RH_Lateral occipito_temporal gyrus (fusiform gyrus, O4_T4) | 3.2 | 7.6 |
| **session 2** | RH_Lingual gyrus, ligual part of the medial occipito_temporal gyrus, (O5) | 1.4 | 1.3 |
| **session 2** | RH_Parahippocampal gyrus, parahippocampal part of the medial occipito_temporal gyrus, (T5) | 1.4 | 6.3 |
| **session 2** | RH_Orbital gyri | 2.8 | 6.3 |
| **session 2** | RH_Angular gyrus | 2.8 | 5.1 |
| **session 2** | RH_Supramarginal gyrus | 2.8 | 10.1 |
| **session 2** | RH_Superior parietal lobule (lateral part of P1) | 1.8 | 1.3 |
| **session 2** | RH_Postcentral gyrus | 0.9 | 3.8 |
| **session 2** | RH_Precentral gyrus | 4.1 | 6.3 |

| | | | |
|---|---|---|---|
| **session 2** | RH_Precuneus (medial part of P1) | 3.7 | 2.5 |
| **session 2** | RH_Straight gyrus, Gyrus rectus | 1.4 | 1.3 |
| **session 2** | RH_Subcallosal area, subcallosal gyrus | 0.5 | 0.0 |
| **session 2** | RH_Anterior transverse temporal gyrus (of Heschl) | 3.7 | 5.1 |
| **session 2** | RH_Lateral aspect of the superior temporal gyrus | 1.4 | 5.1 |
| **session 2** | RH_Planum polare of the superior temporal gyrus | 0.5 | 5.1 |
| **session 2** | RH_Planum temporale or temporal plane of the superior temporal gyrus | 1.8 | 3.8 |
| **session 2** | RH_Inferior temporal gyrus (T3) | 2.3 | 3.8 |
| **session 2** | RH_Middle temporal gyrus (T2) | 3.7 | 7.6 |
| **session 2** | RH_Horizontal ramus of the anterior segment of the lateral sulcus (or fissure) | 0.5 | 1.3 |
| **session 2** | RH_Vertical ramus of the anterior segment of the lateral sulcus (or fissure) | 2.3 | 3.8 |
| **session 2** | RH_Posterior ramus (or segment) of the lateral sulcus (or fissure) | 0.9 | 2.5 |
| **session 2** | RH_Occipital pole | 1.4 | 1.3 |
| **session 2** | RH_Temporal pole | 6.0 | 7.6 |
| **session 2** | RH_Calcarine sulcus | 1.4 | 3.8 |
| **session 2** | RH_Central sulcus (Rolandos fissure) | 2.3 | 0.0 |
| **session 2** | RH_Marginal branch (or part) of the cingulate sulcus | 1.8 | 10.1 |
| **session 2** | RH_Anterior segment of the circular sulcus of the insula | 2.3 | 1.3 |
| **session 2** | RH_Inferior segment of the circular sulcus of the insula | 0.9 | 2.5 |
| **session 2** | RH_Superior segment of the circular sulcus of the insula | 0.9 | 13.9 |
| **session 2** | RH_Anterior transverse collateral sulcus | 2.8 | 3.8 |
| **session 2** | RH_Posterior transverse collateral sulcus | 2.3 | 2.5 |
| **session 2** | RH_Inferior frontal sulcus | 3.7 | 8.9 |
| **session 2** | RH_Middle frontal sulcus | 0.9 | 7.6 |
| **session 2** | RH_Superior frontal sulcus | 3.2 | 5.1 |
| **session 2** | RH_Sulcus intermedius primus (of Jensen) | 2.8 | 3.8 |
| **session 2** | RH_Intraparietal sulcus (interparietal sulcus) and transverse parietal sulci | 4.6 | 3.8 |
| **session 2** | RH_Middle occipital sulcus and lunatus sulcus | 2.8 | 2.5 |
| **session 2** | RH_Superior occipital sulcus and transverse occipital sulcus | 3.2 | 5.1 |
| **session 2** | RH_Anterior occipital sulcus and preoccipital notch (temporo_occipital incisure) | 5.0 | 7.6 |
| **session 2** | RH_Lateral occipito_temporal sulcus | 2.3 | 10.1 |
| **session 2** | RH_Medial occipito_temporal sulcus (collateral sulcus) and lingual sulcus | 1.8 | 3.8 |
| **session 2** | RH_Lateral orbital sulcus | 0.5 | 6.3 |
| **session 2** | RH_Medial orbital sulcus (olfactory sulcus) | 0.9 | 1.3 |
| **session 2** | RH_Orbital sulci (H_shaped sulci) | 1.4 | 6.3 |
| **session 2** | RH_Parieto_occipital sulcus (or fissure) | 3.2 | 8.9 |
| **session 2** | RH_Pericallosal sulcus (S of corpus callosum) | 0.0 | 0.0 |
| **session 2** | RH_Postcentral sulcus | 1.8 | 1.3 |
| **session 2** | RH_Inferior part of the precentral sulcus | 3.2 | 11.4 |
| **session 2** | RH_Superior part of the precentral sulcus | 1.8 | 6.3 |
| **session 2** | RH_Suborbital sulcus (sulcus rostrales, supraorbital sulcus) | 0.5 | 0.0 |
| **session 2** | RH_Subparietal sulcus | 6.4 | 7.6 |

| | | | |
|---|---|---|---|
| **session 2** | RH_Inferior temporal sulcus | 3.2 | 6.3 |
| **session 2** | RH_Superior temporal sulcus (parallel sulcus) | 4.6 | 12.7 |
| **session 2** | RH_Transverse temporal sulcus | 3.2 | 3.8 |
| **session 3** | LH_Fronto_marginal gyrus (of Wernicke) and sulcus | 1.8 | 0.0 |
| **session 3** | LH_Inferior occipital gyrus (O3) and sulcus | 1.8 | 0.0 |
| **session 3** | LH_Paracentral lobule and sulcus | 0.0 | 0.0 |
| **session 3** | LH_Subcentral gyrus (central operculum) and sulci | 3.6 | 0.0 |
| **session 3** | LH_Transverse frontopolar gyri and sulci | 1.8 | 0.0 |
| **session 3** | LH_Anterior part of the cingulate gyrus and sulcus (ACC) | 1.8 | 9.1 |
| **session 3** | LH_Middle_anterior part of the cingulate gyrus and sulcus (aMCC) | 0.0 | 0.0 |
| **session 3** | LH_Middle_posterior part of the cingulate gyrus and sulcus (pMCC) | 1.8 | 9.1 |
| **session 3** | LH_Posterior_dorsal part of the cingulate gyrus (dPCC) | 0.0 | 18.2 |
| **session 3** | LH_Posterior_ventral part of the cingulate gyrus (vPCC, isthmus of the cingulate gyrus) | 1.8 | 0.0 |
| **session 3** | LH_Cuneus (O6) | 1.8 | 9.1 |
| **session 3** | LH_Opercular part of the inferior frontal gyrus | 1.8 | 9.1 |
| **session 3** | LH_Orbital part of the inferior frontal gyrus | 1.8 | 0.0 |
| **session 3** | LH_Triangular part of the inferior frontal gyrus | 3.6 | 9.1 |
| **session 3** | LH_Middle frontal gyrus (F2) | 3.6 | 9.1 |
| **session 3** | LH_Superior frontal gyrus (F1) | 5.5 | 0.0 |
| **session 3** | LH_Long insular gyrus and central sulcus of the insula | 1.8 | 0.0 |
| **session 3** | LH_Short insular gyri | 1.8 | 18.2 |
| **session 3** | LH_Middle occipital gyrus (O2, lateral occipital gyrus) | 5.5 | 0.0 |
| **session 3** | LH_Superior occipital gyrus (O1) | 0.0 | 0.0 |
| **session 3** | LH_Lateral occipito_temporal gyrus (fusiform gyrus, O4_T4) | 1.8 | 9.1 |
| **session 3** | LH_Lingual gyrus, ligual part of the medial occipito_temporal gyrus, (O5) | 1.8 | 0.0 |
| **session 3** | LH_Parahippocampal gyrus, parahippocampal part of the medial occipito_temporal gyrus, (T5) | 1.8 | 0.0 |
| **session 3** | LH_Orbital gyri | 7.3 | 0.0 |
| **session 3** | LH_Angular gyrus | 0.0 | 9.1 |
| **session 3** | LH_Supramarginal gyrus | 1.8 | 0.0 |
| **session 3** | LH_Superior parietal lobule (lateral part of P1) | 0.0 | 0.0 |
| **session 3** | LH_Postcentral gyrus | 1.8 | 0.0 |
| **session 3** | LH_Precentral gyrus | 7.3 | 9.1 |
| **session 3** | LH_Precuneus (medial part of P1) | 3.6 | 0.0 |
| **session 3** | LH_Straight gyrus, Gyrus rectus | 1.8 | 9.1 |
| **session 3** | LH_Subcallosal area, subcallosal gyrus | 5.5 | 0.0 |
| **session 3** | LH_Anterior transverse temporal gyrus (of Heschl) | 7.3 | 0.0 |
| **session 3** | LH_Lateral aspect of the superior temporal gyrus | 0.0 | 9.1 |
| **session 3** | LH_Planum polare of the superior temporal gyrus | 3.6 | 0.0 |
| **session 3** | LH_Planum temporale or temporal plane of the superior temporal gyrus | 1.8 | 0.0 |
| **session 3** | LH_Inferior temporal gyrus (T3) | 3.6 | 9.1 |
| **session 3** | LH_Middle temporal gyrus (T2) | 3.6 | 0.0 |
| **session 3** | LH_Horizontal ramus of the anterior segment of the lateral sulcus (or fissure) | 0.0 | 0.0 |

| | | | |
|---|---|---|---|
| **session 3** | LH_Vertical ramus of the anterior segment of the lateral sulcus (or fissure) | 3.6 | 0.0 |
| **session 3** | LH_Posterior ramus (or segment) of the lateral sulcus (or fissure) | 0.0 | 0.0 |
| **session 3** | LH_Occipital pole | 1.8 | 0.0 |
| **session 3** | LH_Temporal pole | 1.8 | 0.0 |
| **session 3** | LH_Calcarine sulcus | 3.6 | 0.0 |
| **session 3** | LH_Central sulcus (Rolandos fissure) | 3.6 | 0.0 |
| **session 3** | LH_Marginal branch (or part) of the cingulate sulcus | 1.8 | 0.0 |
| **session 3** | LH_Anterior segment of the circular sulcus of the insula | 1.8 | 0.0 |
| **session 3** | LH_Inferior segment of the circular sulcus of the insula | 1.8 | 9.1 |
| **session 3** | LH_Superior segment of the circular sulcus of the insula | 1.8 | 0.0 |
| **session 3** | LH_Anterior transverse collateral sulcus | 1.8 | 0.0 |
| **session 3** | LH_Posterior transverse collateral sulcus | 0.0 | 0.0 |
| **session 3** | LH_Inferior frontal sulcus | 3.6 | 0.0 |
| **session 3** | LH_Middle frontal sulcus | 5.5 | 0.0 |
| **session 3** | LH_Superior frontal sulcus | 3.6 | 0.0 |
| **session 3** | LH_Sulcus intermedius primus (of Jensen) | 0.0 | 9.1 |
| **session 3** | LH_Intraparietal sulcus (interparietal sulcus) and transverse parietal sulci | 0.0 | 0.0 |
| **session 3** | LH_Middle occipital sulcus and lunatus sulcus | 1.8 | 0.0 |
| **session 3** | LH_Superior occipital sulcus and transverse occipital sulcus | 0.0 | 0.0 |
| **session 3** | LH_Anterior occipital sulcus and preoccipital notch (temporo_occipital incisure) | 0.0 | 0.0 |
| **session 3** | LH_Lateral occipito_temporal sulcus | 1.8 | 0.0 |
| **session 3** | LH_Medial occipito_temporal sulcus (collateral sulcus) and lingual sulcus | 0.0 | 0.0 |
| **session 3** | LH_Lateral orbital sulcus | 0.0 | 0.0 |
| **session 3** | LH_Medial orbital sulcus (olfactory sulcus) | 0.0 | 0.0 |
| **session 3** | LH_Orbital sulci (H_shaped sulci) | 1.8 | 0.0 |
| **session 3** | LH_Parieto_occipital sulcus (or fissure) | 0.0 | 0.0 |
| **session 3** | LH_Pericallosal sulcus (S of corpus callosum) | 0.0 | 0.0 |
| **session 3** | LH_Postcentral sulcus | 5.5 | 0.0 |
| **session 3** | LH_Inferior part of the precentral sulcus | 1.8 | 0.0 |
| **session 3** | LH_Superior part of the precentral sulcus | 5.5 | 9.1 |
| **session 3** | LH_Suborbital sulcus (sulcus rostrales, supraorbital sulcus) | 3.6 | 0.0 |
| **session 3** | LH_Subparietal sulcus | 3.6 | 9.1 |
| **session 3** | LH_Inferior temporal sulcus | 1.8 | 0.0 |
| **session 3** | LH_Superior temporal sulcus (parallel sulcus) | 0.0 | 9.1 |
| **session 3** | LH_Transverse temporal sulcus | 0.0 | 0.0 |
| **session 3** | RH_Fronto_marginal gyrus (of Wernicke) and sulcus | 5.5 | 0.0 |
| **session 3** | RH_Inferior occipital gyrus (O3) and sulcus | 5.5 | 0.0 |
| **session 3** | RH_Paracentral lobule and sulcus | 1.8 | 9.1 |
| **session 3** | RH_Subcentral gyrus (central operculum) and sulci | 0.0 | 0.0 |
| **session 3** | RH_Transverse frontopolar gyri and sulci | 1.8 | 9.1 |
| **session 3** | RH_Anterior part of the cingulate gyrus and sulcus (ACC) | 0.0 | 0.0 |
| **session 3** | RH_Middle_anterior part of the cingulate gyrus and sulcus (aMCC) | 0.0 | 0.0 |
| **session 3** | RH_Middle_posterior part of the cingulate gyrus and sulcus (pMCC) | 0.0 | 0.0 |

| | | | |
|---|---|---|---|
| **session 3** | RH_Posterior_dorsal part of the cingulate gyrus (dPCC) | 0.0 | 9.1 |
| **session 3** | RH_Posterior_ventral part of the cingulate gyrus (vPCC, isthmus of the cingulate gyrus) | 0.0 | 0.0 |
| **session 3** | RH_Cuneus (O6) | 1.8 | 0.0 |
| **session 3** | RH_Opercular part of the inferior frontal gyrus | 0.0 | 0.0 |
| **session 3** | RH_Orbital part of the inferior frontal gyrus | 1.8 | 0.0 |
| **session 3** | RH_Triangular part of the inferior frontal gyrus | 7.3 | 0.0 |
| **session 3** | RH_Middle frontal gyrus (F2) | 5.5 | 0.0 |
| **session 3** | RH_Superior frontal gyrus (F1) | 0.0 | 0.0 |
| **session 3** | RH_Long insular gyrus and central sulcus of the insula | 1.8 | 0.0 |
| **session 3** | RH_Short insular gyri | 0.0 | 9.1 |
| **session 3** | RH_Middle occipital gyrus (O2, lateral occipital gyrus) | 1.8 | 0.0 |
| **session 3** | RH_Superior occipital gyrus (O1) | 3.6 | 0.0 |
| **session 3** | RH_Lateral occipito_temporal gyrus (fusiform gyrus, O4_T4) | 3.6 | 0.0 |
| **session 3** | RH_Lingual gyrus, ligual part of the medial occipito_temporal gyrus, (O5) | 1.8 | 0.0 |
| **session 3** | RH_Parahippocampal gyrus, parahippocampal part of the medial occipito_temporal gyrus, (T5) | 3.6 | 0.0 |
| **session 3** | RH_Orbital gyri | 3.6 | 9.1 |
| **session 3** | RH_Angular gyrus | 1.8 | 0.0 |
| **session 3** | RH_Supramarginal gyrus | 3.6 | 0.0 |
| **session 3** | RH_Superior parietal lobule (lateral part of P1) | 0.0 | 0.0 |
| **session 3** | RH_Postcentral gyrus | 3.6 | 9.1 |
| **session 3** | RH_Precentral gyrus | 7.3 | 9.1 |
| **session 3** | RH_Precuneus (medial part of P1) | 1.8 | 0.0 |
| **session 3** | RH_Straight gyrus, Gyrus rectus | 0.0 | 0.0 |
| **session 3** | RH_Subcallosal area, subcallosal gyrus | 0.0 | 0.0 |
| **session 3** | RH_Anterior transverse temporal gyrus (of Heschl) | 3.6 | 0.0 |
| **session 3** | RH_Lateral aspect of the superior temporal gyrus | 0.0 | 18.2 |
| **session 3** | RH_Planum polare of the superior temporal gyrus | 3.6 | 0.0 |
| **session 3** | RH_Planum temporale or temporal plane of the superior temporal gyrus | 0.0 | 0.0 |
| **session 3** | RH_Inferior temporal gyrus (T3) | 1.8 | 0.0 |
| **session 3** | RH_Middle temporal gyrus (T2) | 1.8 | 0.0 |
| **session 3** | RH_Horizontal ramus of the anterior segment of the lateral sulcus (or fissure) | 3.6 | 0.0 |
| **session 3** | RH_Vertical ramus of the anterior segment of the lateral sulcus (or fissure) | 0.0 | 0.0 |
| **session 3** | RH_Posterior ramus (or segment) of the lateral sulcus (or fissure) | 0.0 | 0.0 |
| **session 3** | RH_Occipital pole | 0.0 | 0.0 |
| **session 3** | RH_Temporal pole | 0.0 | 0.0 |
| **session 3** | RH_Calcarine sulcus | 0.0 | 0.0 |
| **session 3** | RH_Central sulcus (Rolandos fissure) | 0.0 | 0.0 |
| **session 3** | RH_Marginal branch (or part) of the cingulate sulcus | 1.8 | 0.0 |
| **session 3** | RH_Anterior segment of the circular sulcus of the insula | 1.8 | 0.0 |
| **session 3** | RH_Inferior segment of the circular sulcus of the insula | 3.6 | 9.1 |
| **session 3** | RH_Superior segment of the circular sulcus of the insula | 3.6 | 0.0 |
| **session 3** | RH_Anterior transverse collateral sulcus | 0.0 | 0.0 |

| | | | |
|---|---|---|---|
| **session 3** | RH_Posterior transverse collateral sulcus | 3.6 | 0.0 |
| **session 3** | RH_Inferior frontal sulcus | 1.8 | 0.0 |
| **session 3** | RH_Middle frontal sulcus | 0.0 | 0.0 |
| **session 3** | RH_Superior frontal sulcus | 0.0 | 0.0 |
| **session 3** | RH_Sulcus intermedius primus (of Jensen) | 1.8 | 18.2 |
| **session 3** | RH_Intraparietal sulcus (interparietal sulcus) and transverse parietal sulci | 5.5 | 0.0 |
| **session 3** | RH_Middle occipital sulcus and lunatus sulcus | 3.6 | 9.1 |
| **session 3** | RH_Superior occipital sulcus and transverse occipital sulcus | 1.8 | 9.1 |
| **session 3** | RH_Anterior occipital sulcus and preoccipital notch (temporo_occipital incisure) | 5.5 | 0.0 |
| **session 3** | RH_Lateral occipito_temporal sulcus | 7.3 | 0.0 |
| **session 3** | RH_Medial occipito_temporal sulcus (collateral sulcus) and lingual sulcus | 0.0 | 0.0 |
| **session 3** | RH_Lateral orbital sulcus | 0.0 | 0.0 |
| **session 3** | RH_Medial orbital sulcus (olfactory sulcus) | 3.6 | 0.0 |
| **session 3** | RH_Orbital sulci (H_shaped sulci) | 3.6 | 0.0 |
| **session 3** | RH_Parieto_occipital sulcus (or fissure) | 1.8 | 0.0 |
| **session 3** | RH_Pericallosal sulcus (S of corpus callosum) | 0.0 | 0.0 |
| **session 3** | RH_Postcentral sulcus | 5.5 | 0.0 |
| **session 3** | RH_Inferior part of the precentral sulcus | 0.0 | 0.0 |
| **session 3** | RH_Superior part of the precentral sulcus | 0.0 | 9.1 |
| **session 3** | RH_Suborbital sulcus (sulcus rostrales, supraorbital sulcus) | 0.0 | 9.1 |
| **session 3** | RH_Subparietal sulcus | 0.0 | 0.0 |
| **session 3** | RH_Inferior temporal sulcus | 5.5 | 0.0 |
| **session 3** | RH_Superior temporal sulcus (parallel sulcus) | 1.8 | 0.0 |
| **session 3** | RH_Transverse temporal sulcus | 3.6 | 0.0 |

### Supplementary References

1. Destrieux C, Fischl B, Dale A, Halgren E. Automatic parcellation of human cortical gyri and sulci using standard anatomical nomenclature. *Neuroimage*. 2010;53(1):1-15. doi:10.1016/j.neuroimage.2010.06.010
2. Bethlehem RAI, Seidlitz J, White SR, et al. Brain charts for the human lifespan. *Nature*. 2022;604(7906):525-533. doi:10.1038/s41586-022-04554-y
3. Ducharme S, Albaugh MD, Nguyen TV, et al. Trajectories of cortical thickness maturation in normal brain development — The importance of quality control procedures. *Neuroimage*. 2016;125:267-279. doi:10.1016/j.neuroimage.2015.10.010
4. Frangou S, Modabbernia A, Williams SCR, et al. Cortical thickness across the lifespan: Data from 17,075 healthy individuals aged 3–90 years. *Hum Brain Mapp*. 2022;43(1):431-451. doi:10.1002/hbm.25364
5. Kia SM, Huijsdens H, Dinga R, et al. Hierarchical Bayesian Regression for Multi-Site Normative Modeling of Neuroimaging Data. Published online May 25, 2020:1-12. http://arxiv.org/abs/2005.12055
6. Nesvåg R, Jönsson EG, Bakken IJ, et al. The quality of severe mental disorder diagnoses in a national health registry as compared to research diagnoses based on structured interview. *BMC Psychiatry*. 2017;17(1):93. doi:10.1186/s12888-017-1256-8

7. Knussmann GN, Anderson JS, Prigge MBD, et al. Test-retest reliability of FreeSurfer-derived volume, area and cortical thickness from MPRAGE and MP2RAGE brain MRI images. *Neuroimage: Reports*. 2022;2(2):100086. doi:10.1016/j.ynirp.2022.100086
8. Hedges EP, Dimitrov M, Zahid U, et al. Reliability of structural MRI measurements: The effects of scan session, head tilt, inter-scan interval, acquisition sequence, FreeSurfer version and processing stream. *Neuroimage*. 2022;246(November 2021):118751. doi:10.1016/j.neuroimage.2021.118751
9. Schulz MA, Bzdok D, Haufe S, Haynes JD, Ritter K. Performance reserves in brain-imaging-based phenotype prediction. *bioRxiv*. Published online 2022:2022.02.23.481601. https://www.biorxiv.org/content/10.1101/2022.02.23.481601v1%0Ahttps://www.biorxiv.org/content/10.1101/2022.02.23.481601v1.abstract
10. Ayesa-Arriola R, de la Foz VOG, Setién-Suero E, et al. Understanding sex differences in long-term outcomes after a first episode of psychosis. *NPJ Schizophr*. 2020;6(1):1-8. doi:10.1038/s41537-020-00120-5
11. Kjelkenes R, Wolfers T, Alnæs D, et al. Deviations from normative brain white and gray matter structure are associated with psychopathology in youth. *Dev Cogn Neurosci*. 2022;58:101173. doi:10.1016/j.dcn.2022.101173
12. Kjelkenes R, Wolfers T, Alnæs D, et al. Mapping Normative Trajectories of Cognitive Function and Its Relation to Psychopathology Symptoms and Genetic Risk in Youth. *Biological Psychiatry Global Open Science*. Published online 2022:18-20. doi:10.1016/j.bpsgos.2022.01.007
13. Andreasen NC, Carpenter WT, Kane JM, Lasser RA, Marder SR, Weinberger DR. Remission in Schizophrenia: Proposed Criteria and Rationale for Consensus. *American Journal of Psychiatry*. 2005;162(3):441-449. doi:10.1176/appi.ajp.162.3.441
14. Salazar De Pablo G, Radua J, Pereira J, et al. Probability of Transition to Psychosis in Individuals at Clinical High Risk: An Updated Meta-analysis. *JAMA Psychiatry*. Published online 2021:1-9. doi:10.1001/jamapsychiatry.2021.0830
15. Leucht S, Samara M, Heres S, Davis JM. Dose Equivalents for Antipsychotic Drugs : The DDD Method. 2016;42(1):1-3. doi:10.1093/schbul/sbv167
16. Remiszewski N, Bryant JE, Rutherford SE, et al. Contrasting Case-Control and Normative Reference Approaches to Capture Clinically Relevant Structural Brain Abnormalities in Patients With First-Episode Psychosis Who Are Antipsychotic Naive. *JAMA Psychiatry*. 2022;79(11):1133. doi:10.1001/jamapsychiatry.2022.3010
